# TEMR: Trans-ethnic Mendelian Randomization Method using Large-scale GWAS Summary Datasets

**DOI:** 10.1101/2024.06.16.24308874

**Authors:** Lei Hou, Sijia Wu, Zhongshang Yuan, Hongkai Li, Fuzhong Xue

## Abstract

Available large-scale GWAS summary datasets predominantly stem from European populations, while sample sizes for other ethnicities, notably Central/South Asian, East Asian, African, Hispanic, etc. remain comparatively limited, which induces the low precision of causal effect estimation within these ethnicities using Mendelian Randomization (MR). In this paper, we propose a Trans-ethnic MR method called TEMR to improve statistical power and estimation precision of MR in the target population using trans-ethnic large-scale GWAS summary datasets. TEMR incorporates trans-ethnic genetic correlation coefficients through a conditional likelihood-based inference framework, producing calibrated p-values with substantially improved MR power. In the simulation study, TEMR exhibited superior precision and statistical power in the causal effects estimation within the target populations than other existing MR methods. Finally, we applied TEMR to infer causal relationships from 17 blood biomarkers to four diseases (hypertension, ischemic stroke, type 2 diabetes and schizophrenia) in East Asian, African and Hispanic/Latino populations leveraging the biobank-scale GWAS summary data from European. We found that causal biomarkers were mostly validated by previous MR methods, and we also discovered 13 new causal relationships that were not identified using previously published MR methods.

## Introduction

In recent years, the evolving landscape has witnessed a progressive expansion of large-scale Genome-Wide Association Studies (GWAS), leading to the widespread release and utilization of GWAS summary data among researchers. At the forefront of these developments is Mendelian Randomization (MR) ^[1-2]^, a method that hinges on the use of publicly available GWAS summary data for causal inference. MR uses genetic variants as instrumental variables (IVs) to infer causal effect of an exposure on an outcome. It requires three assumptions: Relevance, IVs are strongly associated with the exposure; Exchangeability, IVs are independent with confounders among the exposure and outcome; Exclusion restriction, IVs affect the outcome only through the exposure. However, a noteworthy challenge surfaces as the bulk of available large-scale datasets predominantly stem from European populations, such as the UK Biobank (UKB) ^[3-7]^ and FinnGen consortium ^[8]^, while sample sizes for other ethnicities, notably Central/South Asian, East Asian, African, Hispanic, etc. remain comparatively limited ^[9-12]^. Take the East Asian population as an example, despite the substantial data provided by the BioBank Japan (BBJ) ^[11-12]^, Taiwan Biobank (TWB) ^[13]^ and China Kadoorie Biobank (CKB) ^[14]^ for the East Asian population (> 100,000 individuals), it falls short of the extensive dataset available from the UKB (> 500,000 individuals) or FinnGen consortium (> 620,000 individuals). Moreover, the UKB incorporates a substantial amount of omics data, including imaging omics ^[4]^, exomes ^[5]^, proteomics ^[6]^ and metabolomics ^[7]^—BBJ, TWB and CKB have significantly smaller sample sizes and may also lack some omics data ^[12]^. Furthermore, omics databases dedicated to other ethnicities tend to exhibit relatively smaller sample sizes ^[15-21]^. The potential inadequacy of GWAS summary data from smaller samples to furnish robust causal evidence for MR becomes apparent. Additionally, causal evidence derived from a substantial European population cannot be directly extrapolated to other ethnic groups due to diversity in the genetic structure between different ethnicities ^[22-23]^. The unbalanced sample makeup across global populations may exacerbate the disparities in genetic studies of non-Europeans. Therefore, it is crucial to propose a methodology that leverages the genetic correlations ^[24-25]^ among different ethnicities, harnessing the advantages of large European datasets to enhance the accuracy and statistical power of MR in estimating causal effects within smaller populations.

A number of trans-ethnic MR analyses has been published, predominantly featured in applied research articles ^[26-28]^. The common approach in these studies involves conducting separate MR analyses within distinct ethnic groups and subsequently comparing the nuances in the MR results between these groups. This is unfair for ethnicities with small sample sizes, as its statistical power of MR is much lower than that of large sample sizes. For methodology, advancements have been made in cross-ethnic approaches within GWAS meta-analysis and Polygenic Risk Score (PRS). The published trans-ethnic meta-analysis approaches take into account the similarity in allelic effects between the most closely related populations while allowing for heterogeneity between more diverse ethnic group ^[29-32]^. While trans-ethnic GWAS meta-analysis has the potential to improve the efficiency of identifying new loci by merging populations of different ethnicities, it operates at a mixed-population level and may not necessarily contribute to the discovery of genetic loci specific to particular ethnic groups. Tans-ethnic PRS prediction methods leverage shared genetic effects across ancestries to increase the accuracy of predicting the genetic predisposition of complex phenotypes in non-European populations ^[33-35]^. However, these methods highlight that improving the power to discovery new loci or disease prediction, a noticeable gap in the current literature lies in the lack of attention to methods facilitating the transfer of causal effects in MR across different ethnicities. Despite progress in various methodological aspects of trans-ethnic analysis, there remains an unexplored avenue concerning the migration of causal effects across ethnic groups in the context of MR.

In this paper, we propose a MR method based on Trans-ethnic Population called TEMR to improve statistical power and estimation precision of MR in target population using trans-ethnic large-scale GWAS summary datasets. Under the framework of conditional likelihood-based inference framework, TEMR bridges the causal effects of different ethnics using a trans-ethnic genetic correlation coefficient, which is the correlation of Wald ratios for shared SNPs in different ethnic populations. In the simulation study, TEMR showed superior precision and power of causal effect estimation in the target population relative to other seven methods in the case of continuous and binary outcome variables. Finally, we apply TEMR to infer causal relationships from 17 blood biomarkers to four diseases (hypertension, ischemic stroke, type 2 diabetes (T2D) and schizophrenia) in East Asian, African and Hispanic/Latino populations leveraging the biobank-scale GWAS summary data from European.

## Results

### TEMR Method overview

We consider a target dataset {*G*_1_, *X*_1_,*Y*_1_} from an under-represented ancestry (e.g. East Asian, African and Hispanic/Latino population, etc) with small sample size, where *G*_1_, *X*_1_ and *Y*_1_ represent the Single Nucleotide Polymorphisms (SNPs), exposure and outcome, respectively. Now suppose we have an auxiliary dataset {*G*_2_, *X*_2_,*Y*_2_} (e.g. European population) with a biobank-scale sample size available. We assume the sample sizes of two datasets satisfy the condition *N*_2_ □ *N*_1_. We choose the *p* independent SNPs as IVs, which are associated with at least one of exposures in two ancestries (*X*_1_ and *X*_2_). The workflow of TEMR is shown in Figure 1.

**Figure 1.**
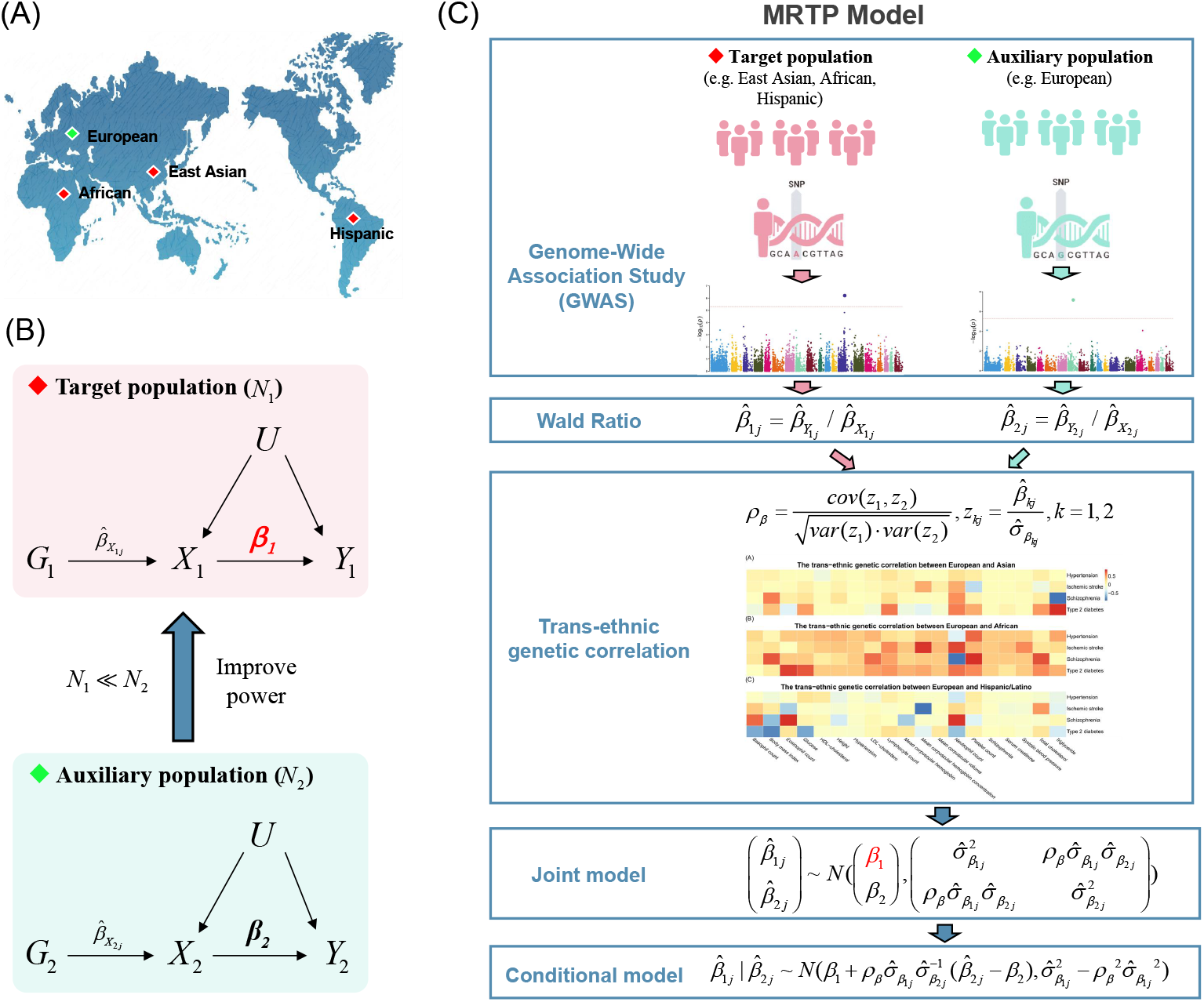
TEMR flowchart. (A) The example of multiple ethnics, which also are the ethnics that we are interested in the applied example. (B) The aim of TEMR is to improve the statistical power and estimation accuracy of MR in target population only using trans-ethnic large-scale auxiliary dataset. (C) The flowchart of TEMR model, take two ethnics as example, one target population and one auxiliary population.

When the three core assumptions of MR are all satisfied, we can obtain the Wald ratio estimation for each SNP: 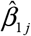 in the target population and 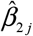 in the target population, as well as their variances 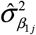 and 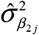, respectively, using the summary-level data of *p* SNPs, including beta-coefficients (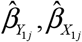 and 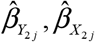) and their standard error (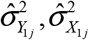 and 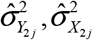). We set up the following multivariable normal distribution model for Wald ratios from two populations:

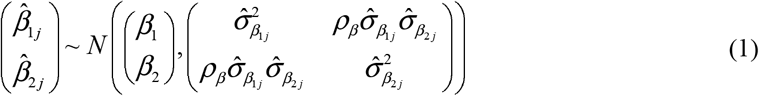

where *β*_1_ and *β*_2_ are the causal effect of exposure on outcome in the target and auxiliary populations, respectively. *ρ*_*β*_ is the trans-ethnic genetic correlation, which represents the correlation of the causal effects of one exposure on one outcome in two ancestries (e.g. East Asian and European). It bridges the causal effects of two ethnics to achieve our aim of improving the statistical power of causal effect (*β*_1_) estimation in the target population. In terms of the conditional normal distribution of 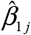 given 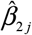,

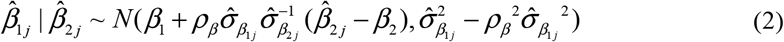

we found that the variance of *j*-th Wald ratio estimation 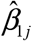 conditional on 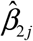 is smaller than its original variance as the trans-ethnic genetic correlation *ρ*_*β*_ increasing 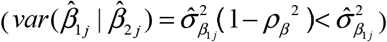. Then we obtain the causal effect (*β*_1_) estimation in the target population by maximizing the log conditional likelihood function using Nelder-Mead method [38]. We use Likelihood-ratio test to perform hypothesis testing.

When there is horizontal pleiotropy, the third assumption of MR is violated, the causal effect estimation using the traditional Wald ratio is biased and we model a new TEMR-Wald ratio by removing the impact of horizontal pleiotropy (*α*_1 *j*_) from 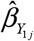. We propose a two-step process to estimate TEMR-Wald ratios for each SNP leveraging MR-Egger regression. Next, we use the new TEMR-Wald ratio to set up model (1), then infer the causal effect (*β*_1_) in the target population, and the remaining steps are the same as in the case of no pleiotropy.

If there are multiple ancestries (*E* ancestries), the target dataset is {*G*_*T*_, *X*_*T*_,*Y*_*T*_ } and the auxiliary datasets are {*G*_*a*_, *X*_*a*_,*Y*_*a*_ }(*a* = 2,…, *E*), we can also set up the multivariable normal distribution model using Wald ratios from *E* ancestries

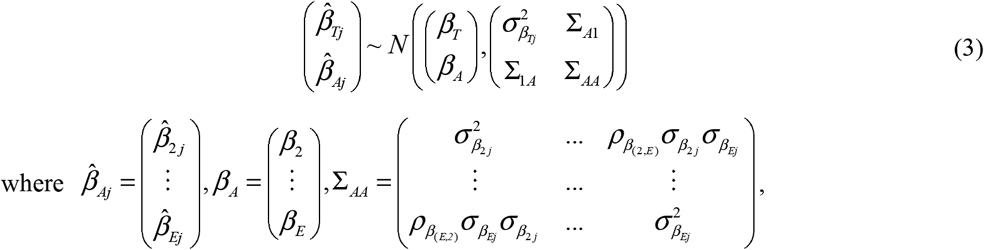

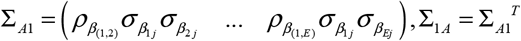 and 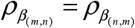. Then we derive the conditional model and obtain the estimation of *β*_*T*_ by Maximum Likelihood Estimation.

### Simulation

We conducted a series of simulation studies to evaluate the performance of TEMR, comprising with seven published MR methods. We vary with magnitudes of parameters: causal effect, trans-ethnic genetic correlation, sample size and the number of SNPs, in the scenarios of no pleiotropy and horizontal pleiotropy. We utilized boxplots to demonstrate the results of estimation bias and standard error, Q-Q plots to showcase the results of Type I error, and bar charts to depict the results of statistical power.

When there is no pleiotropy, Figure 2 shows the simulation results of causal effect estimation in the target population when there is one auxiliary population in the case of continuous outcome. Simulation results demonstrated that TEMR showed nearly unbiased estimates of causal effects regardless of the alignment between causal effects in the auxiliary and target populations. TEMR also showed superior precision and power across a broad spectrum of scenarios relative to other seven methods, which was consistently observed for both continuous and binary outcome variables. The precision of TEMR incrementally improved as the *ρ*_*β*_ increasing. When *ρ*_*β*_ < 0.4, the precision of TEMR was similar with the Inverse-variance weighted method (IVW) and the Weighted Median Estimation (WME) method. However, when *ρ*_*β*_ ≥ 0.4, the precision of TEMR surpassed that of other seven methods (Figure 2A, Figure S1). Additionally, TEMR exhibited stable Type I errors, unaffected by variations in *ρ*_*β*_ or causal effects in the auxiliary population (Figure 2B-C, Figure S2). Moreover, the statistical power of TEMR significantly increased with *ρ*_*β*_ rising, especially outperforming other seven methods when *ρ*_*β*_ ≥ 0.4 (Figure 2D-E, Figure S3-S6). Specifically, when *ρ*_*β*_ = 0.6, there was a notable decrease in the standard error by approximately 15-20%, and an increase in power by about 20%. At a higher *ρ*_*β*_, the standard error could decrease by up to 50%, while the power could improve by 40% (Table S1).

**Figure 2.**
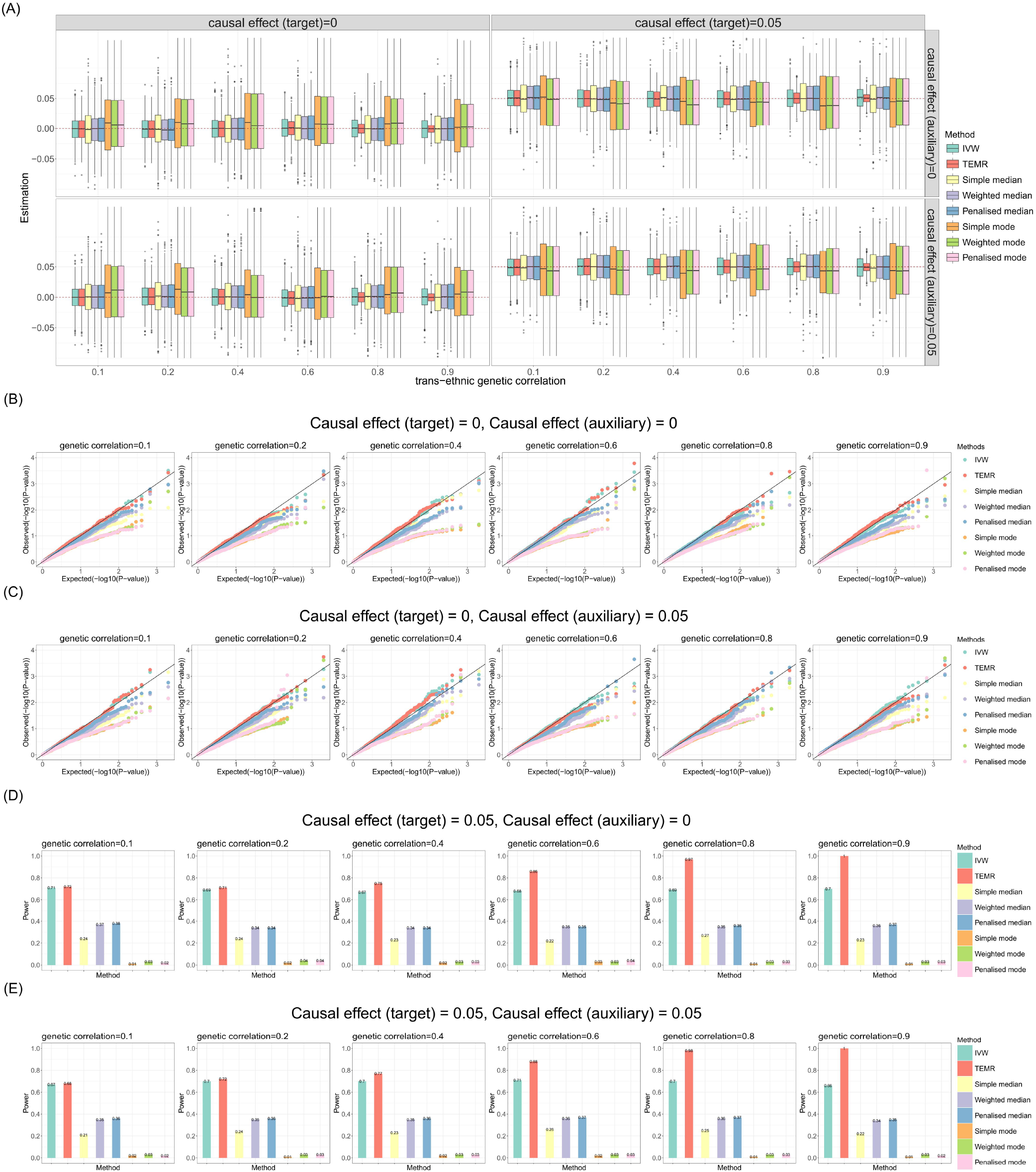
Simulation results for causal effect estimation in the target population when there is one auxiliary population (no pleiotropy). Sample size of target population is 3,000 and the sample size of auxiliary population is 300,000. IVs include 100 common SNPs. **A)** Boxplots show the performances of causal effect estimation in target population; **B-C)** Q-Q plots show the performances of Type I error rates of zero causal effect estimation in target population when the causal effect of auxiliary population is 0 and 0.05, respectively; **D-E)** Bar chart plots show the performances of statistical power of non-zero causal effect estimation in target population when the causal effect of auxiliary population is 0 and 0.05, respectively. IVW, Inverse-variance weighted method.

Then we extended our simulation to scenario where there is horizontal pleiotropy, both balance and directional, are present. In addition to achieving unbiased estimates of causal effects and stable Type I errors, TEMR also maintained the precision and power advantages as described above (Figure 3, Figure S7-18). In cases involving categorical outcome, we obtained results consistent with those for continuous variables (Figure S19-36, Table S2). Additionally, we also observed that the target population can also enhance the precision and test power of causal effect estimates in the auxiliary population, exemplified by scenarios directional horizontal pleiotropy (Figure S37, Table S3). These observations underscore the robustness and effectiveness of TEMR in various genetic correlation contexts.

**Figure 3.**
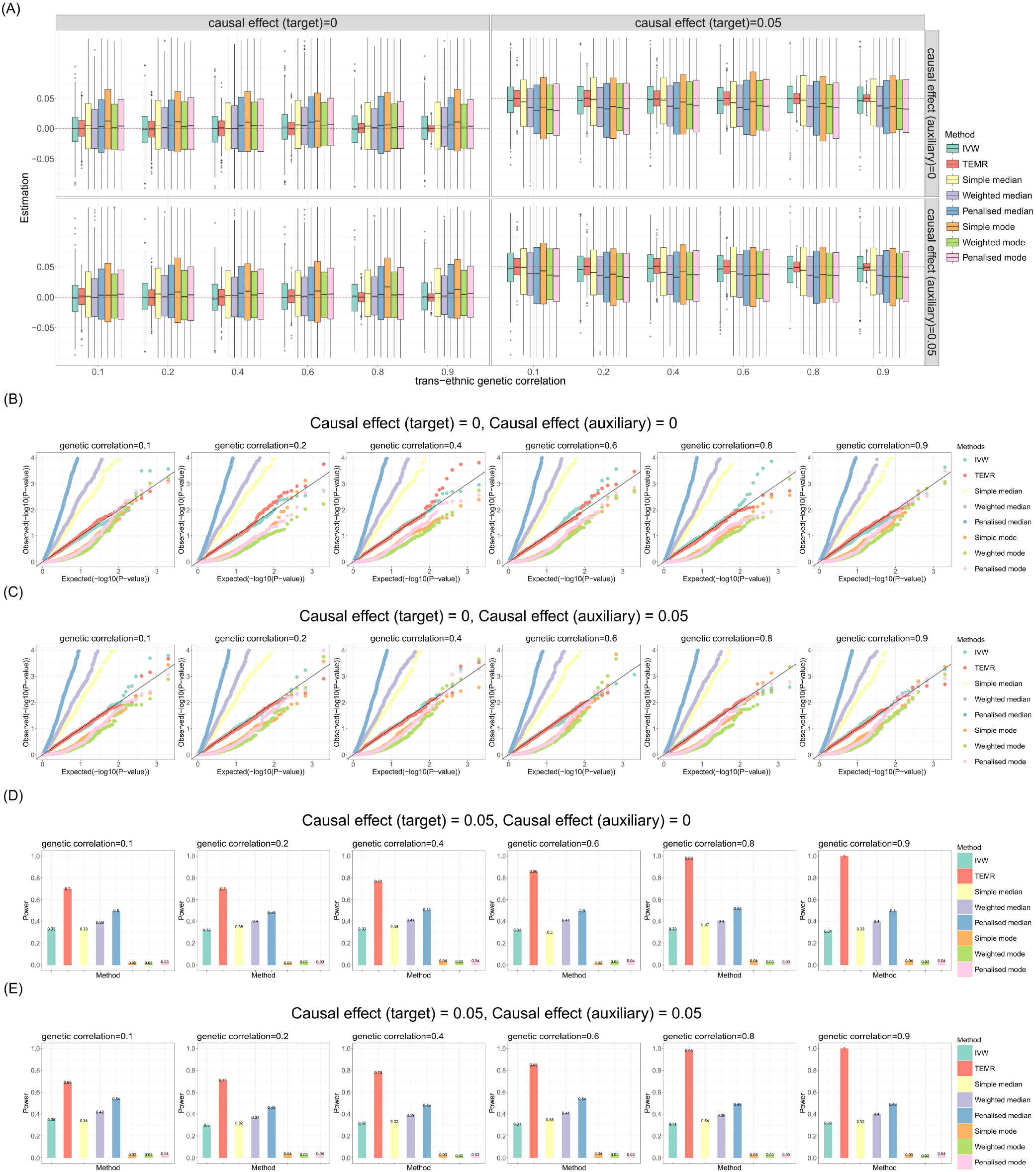
Simulation results for causal effect estimation in the target population when there is one auxiliary population (directional horizontal pleiotropy). Sample size of target population is 3,000, and the sample size of auxiliary population is 300,000. IVs include 100 common SNPs. **A)** Boxplots show the performances of causal effect estimation in target population; **B-C)** Q-Q plots show the performances of Type I error rates of zero causal effect estimation in target population when the causal effect of auxiliary population is 0 and 0.05, respectively; **D-E)** Bar chart plots show the performances of statistical power of non-zero causal effect estimation in target population when the causal effect of auxiliary population is 0 and 0.05, respectively. IVW, Inverse-variance weighted method.

In order to investigate the impact of the number of SNPs (Figure 4, Table S4) and sample size (Figure S38-43, Table S5) on the causal effect estimates, firstly, we conducted a thorough exploration by varying the number of SNPs while maintaining other parameters at their initial settings. Simulation results indicated that an increase in the number of SNPs leads to higher precision and greater test power in the causal effect estimates derived from the TEMR. While other methods also demonstrated improvements with more SNPs, the enhancement was not as pronounced as that observed with TEMR.

**Figure 4.**
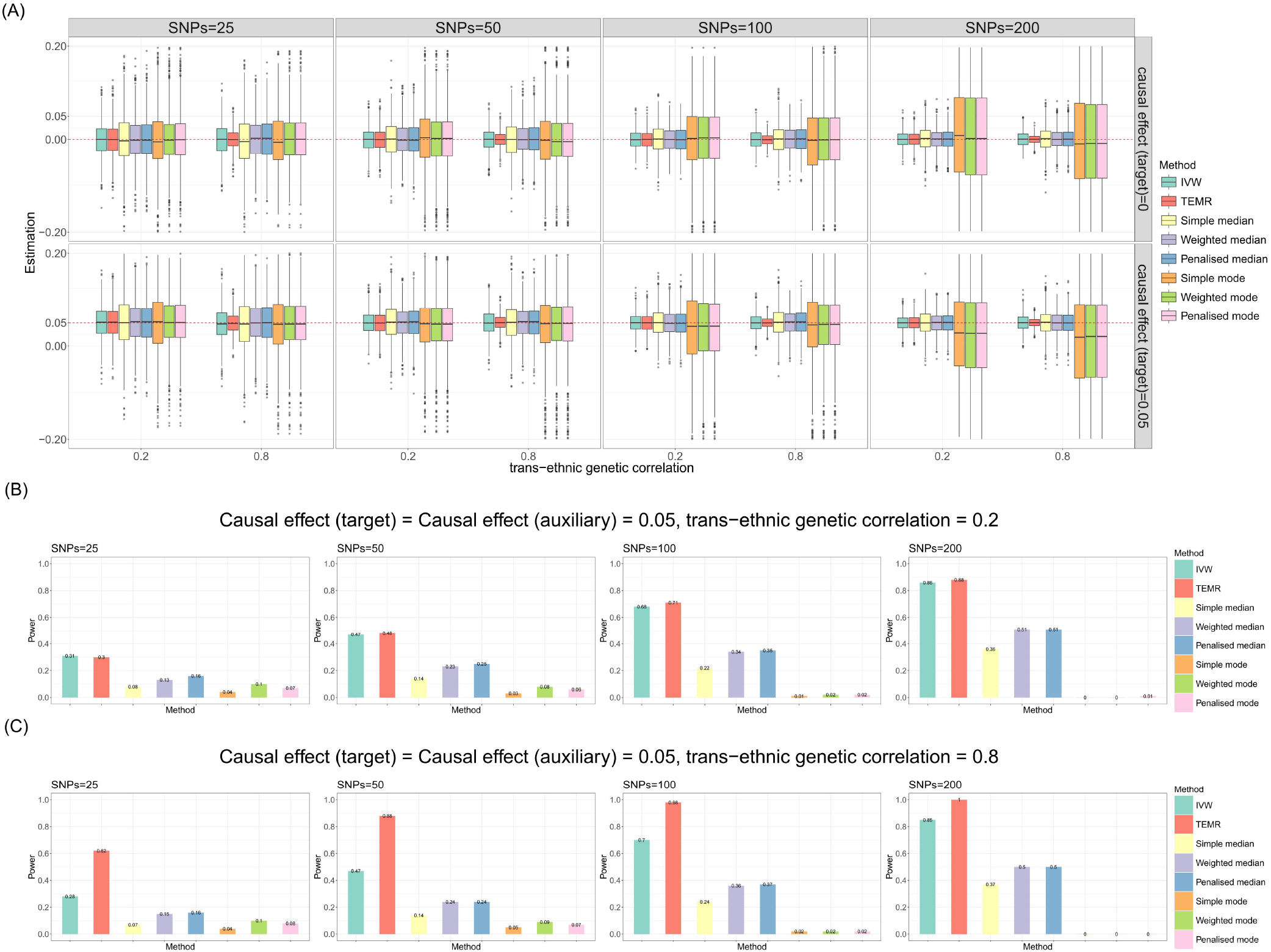
Simulation results for causal effect estimation in the target population with different number of SNPs. Continuous outcome, no horizontal pleiotropy. Sample size of target population is 3,000, and the sample size of auxiliary population is 300,000. **A)** Boxplots show the performances of causal effect estimation in target population; **B-C)** Bar chart plots show the performances of statistical power of non-zero causal effect estimation in target population. IVW, Inverse-variance weighted method.

Additionally, we explored the causal effect estimates using the TEMR when there is a negative genetic correlation between ethnic groups. The results indicated that the precision and the statistical power significantly improved as the absolute value of the genetic correlation increased, which was consistent with the aforementioned findings (Figure S44-45, Table S6).

Subsequently, we consider the case of multiple auxiliary populations, taking three auxiliary populations and one target population as an example, assuming uniform causal effects across different populations. In the absence of horizontal pleiotropy, TEMR produced nearly unbiased estimates of causal effects. And the precision and statistical power of TEMR also incrementally improved as the *ρ*_*β*_ increased, when *ρ*_*β*_ ≥ 0.4, the precision and power of TEMR surpassed that of other methods (Figure 5A). Specifically, when *ρ*_*β*_ = 0.6, there was a notable decrease in the standard error by approximately 15-20%, and an increase in power by about 20%. At a higher *ρ*_*β*_, the standard error could decrease by up to 50%, while the power could improve by 40% (Figure 5C). Furthermore, TEMR exhibited stable Type I errors, unaffected by variations in *ρ*_*β*_ (Figure 5B). In cases involving horizontal pleiotropy, we obtained results consistent with those (Figure S46-54, Table S7). And we also obtained consistent results when the genetic correlations were negative (Figure S55-56, Table S8). Moreover, compared to having only one auxiliary population, the precision (standard error) of the causal effect estimate obtained from three auxiliary populations also improved with the increase in genetic correlations, with an approximate 15% improvement when *ρ*_*β*_ = 0.9.

**Figure 5.**
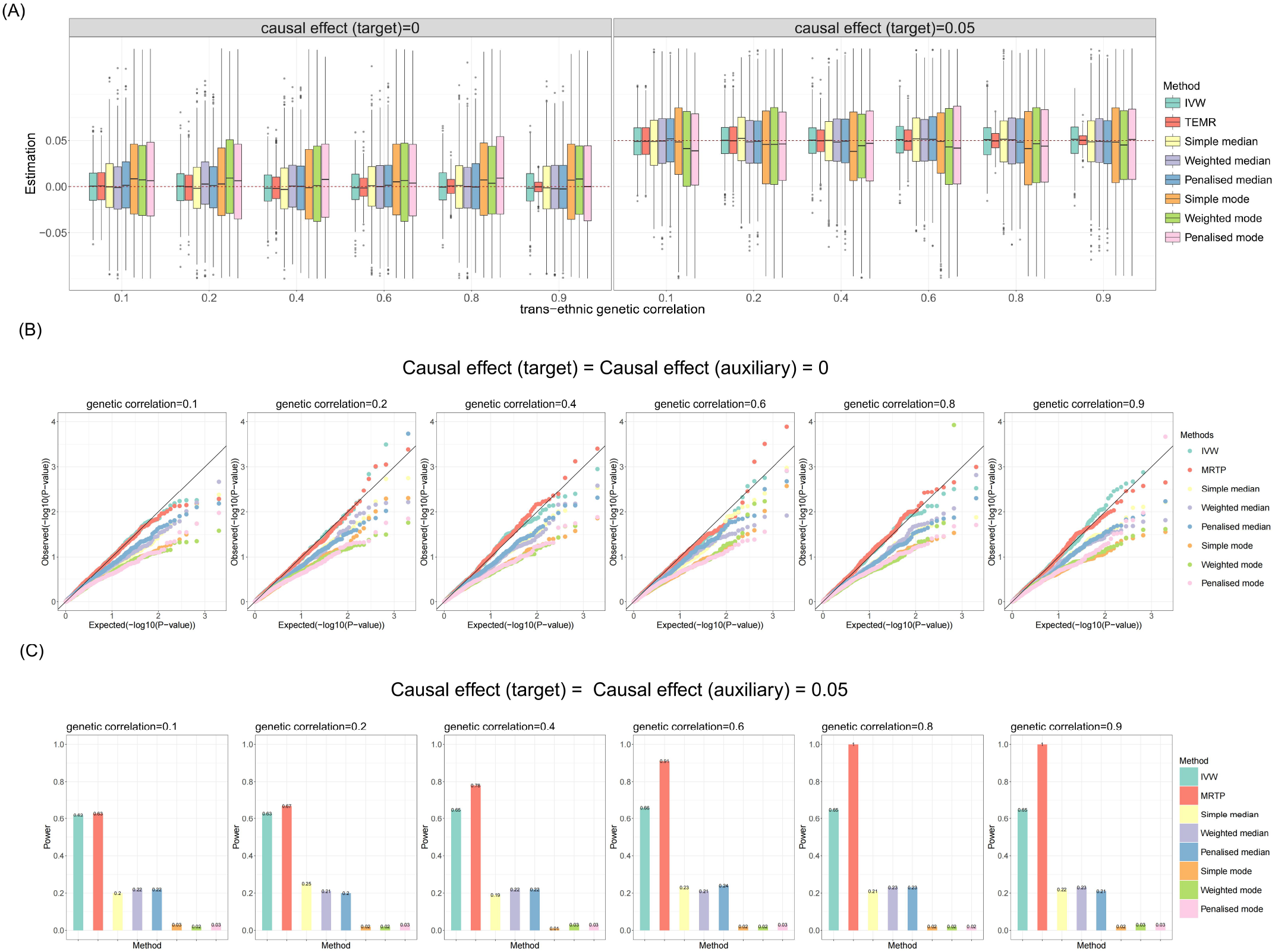
Simulation results for causal effect estimation in the target population when there are multiple auxiliary populations. Continuous outcome. No pleiotropy. Sample size of target population is 3,000, and the sample size of auxiliary populations are 3,000, 3,000, 300,000. IVs include 100 common SNPs. A) Boxplots show the performances of causal effect estimation in target population; B) Q-Q plots show the performances of Type I error rates of zero causal effect estimation in target population; C) Bar chart plots show the performances of statistical power of non-zero causal effect estimation in target population. IVW, Inverse-variance weighted method.

### Application

In this section, we applied TEMR to infer the causal relationships between different biomarkers and four diseases (hypertension, ischemic stroke, T2D and schizophrenia) in the East Asian, African, Hispanic/Latino population, leveraging GWAS summary data from large European cohorts (Table S9). Initially, we identified 17 specific biomarkers that were significantly associated with at least 2 SNPs from a multitude of biomarkers. Then we calculated the trans-ethnic genetic correlation for all pairs of biomarkers, results are shown in Figure 6 (Table S10). The results showed that there were trans-ethnic genetic correlations between the causal effects of all biomarkers and diseases in four populations, which could be analyzed by TEMR. Among these, the absolute value of correlation of 14 pairs exhibited 0.5 (total 17×4=68 pairs), including basophil count to hypertension (between East Asian and Hispanic/Latino), neutrophil count to hypertension (between African, Hispanic/Latino and East Asian), mean corpuscular hemoglobin concentration (MCHC) to ischemic stroke (between East Asian, European and Hispanic/Latino, African and East Asian), eosinophil count to schizophrenia (between European and Hispanic/Latino), neutrophil count to schizophrenia (except between East Asian and Hispanic/Latino), platelet count to schizophrenia (between East Asian, Hispanic/Latino and African), body mass index (BMI) to schizophrenia (between European and East Asian), triglyceride (TG) to schizophrenia (between European and East Asian), lymphocyte count to T2D (between European, Hispanic/Latino and East Asian), BMI to T2D (between European and Hispanic/Latino), glucose to T2D (between European Hispanic/Latino and East Asian), neutrophil count to T2D (between European, African and East Asian), Total cholesterol (TC) to T2D (between European and East Asian), TG to T2D (between European and East Asian).

**Figure 6.**
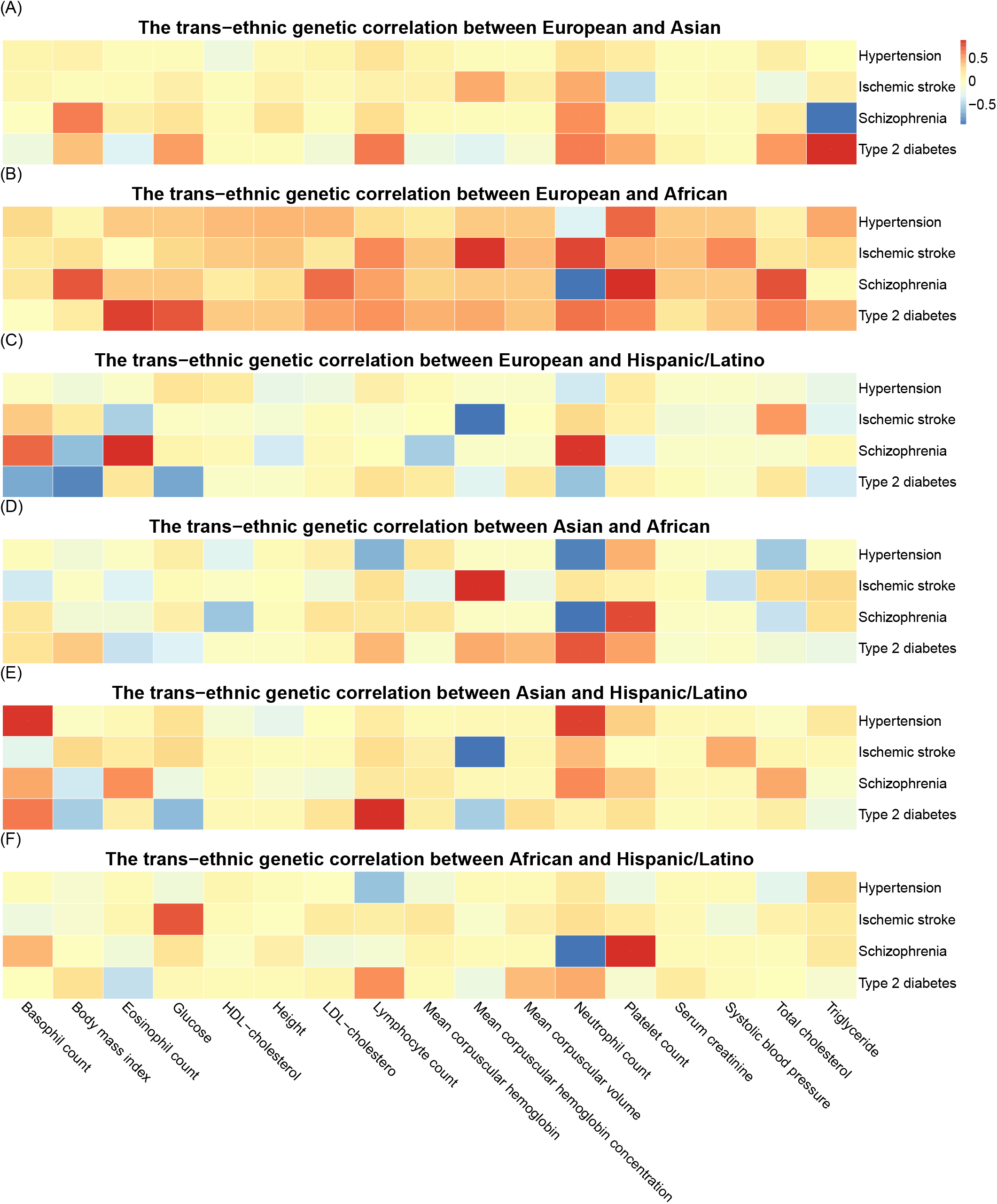
Heatmap of trans-ethnic genetic correlation for 22 biomarkers and four diseases. The color intensity indicates the strength of the correlation. Warmer colors, tending towards red, signify a correlation coefficient approaching 1, indicating a strong positive correlation. Conversely, cooler colors, leaning towards blue, denote a correlation coefficient nearing -1, suggesting a strong negative correlation.

Then we perform trans-ethnic MR analysis using TEMR and other seven methods. The results indicated that TEMR identified a greater number of biomarker pairs with significant causal associations compared to the other seven methods (Figure 7). Among these, TEMR emerged as the method identifying the most significant biomarker pairs in each target population, with IVW following closely behind (Table S11), and most of the significant biomarker pairs identified by the other methods were also detected by TEMR with smaller *P*-values. Notably, there were several significant relationships across different ethnic groups that only TEMR identified as significant (*P*<0.0007(0.05/68)): three new causal relationships in East Asian, four new causal relationships in African and six in Hispanic/Latino population.

**Figure 7.**
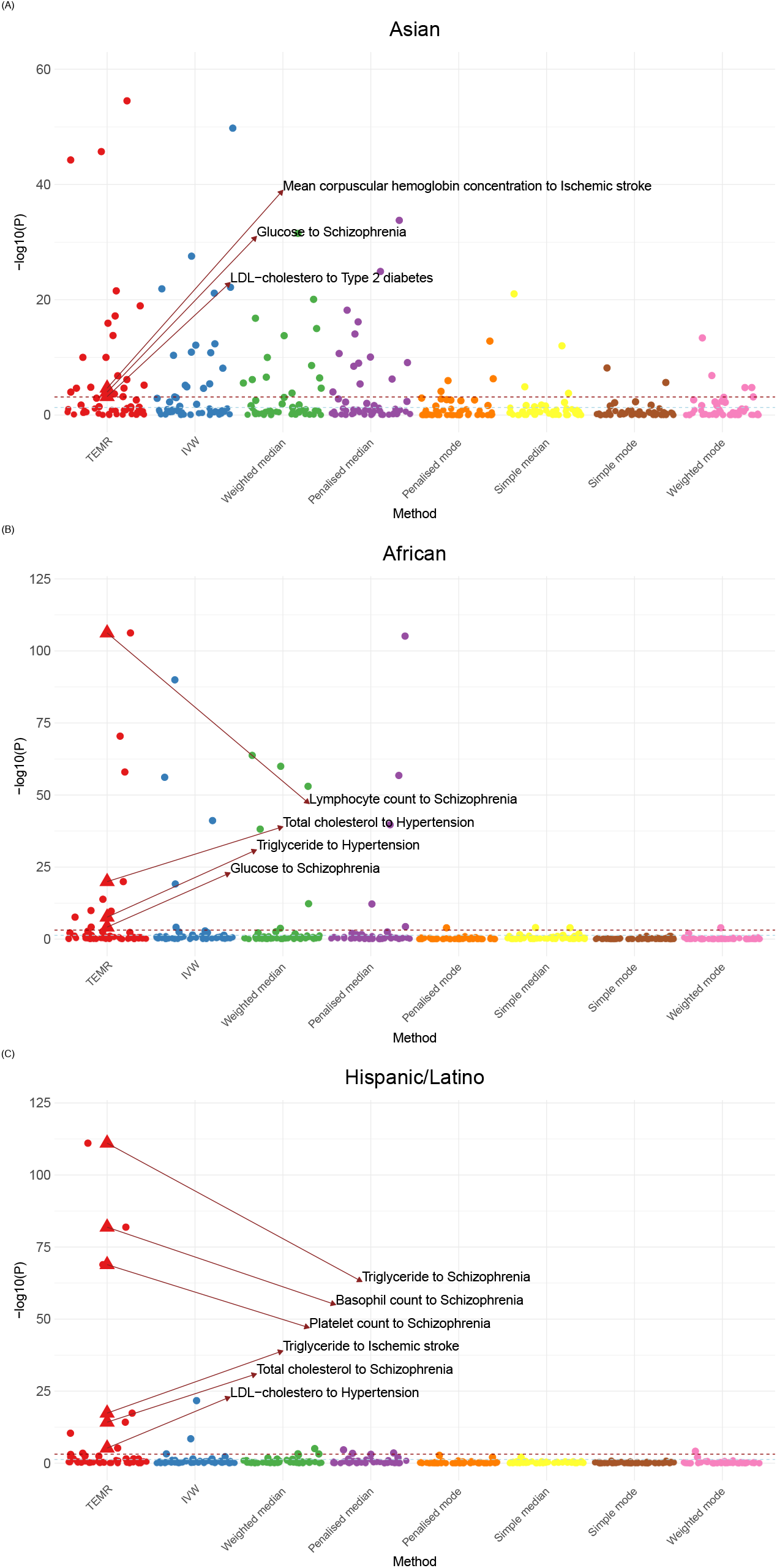
Results of trans-ethnic MR analysis for causal relationships from 22 biomarkers to four diseases. Different colors represent the -log_10_(P) calculated by different methods. The triangle points represent the significant relationships in TEMR results but not significant in other methods. The solid or dashed points indicate whether the causal effects are significant (P<0.05). In cases where the MR-Egger test suggests the presence of horizontal pleiotropy between biomarker pairs, the P-values presented are those adjusted for such pleiotropy.

In the East Asian population, significant causal associations including TC to schizophrenia (*OR*=2.30, *P*=0.019), HDL-cholesterol (HDL-C) to schizophrenia (*OR*=0.50, *P*=0.044) and neutrophil count to T2D (*OR*=0.89, *P*=0.038) were detected (Table S12). The association between TC and schizophrenia suggested that higher TC levels might influence the risk of developing schizophrenia. This connection could be through the alteration of cell membrane fluidity, which in turn may impact neurotransmitter signaling. Many studies supported that elevated serum TC levels could be linked to enhanced cognitive function in individuals with schizophrenia ^[39-41]^. For the relationship between HDL-C and schizophrenia, the inverse association could indicate that higher levels of HDL-C, often considered good cholesterol, might have a protective effect against the development of schizophrenia. Studies corroborated these findings, indicating that patients with schizophrenia often have lower levels of HDL-C compared to those without the condition ^[40]^. Finally, while the relationship between neutrophil count and the risk of T2D is complex and not fully understood, some studies have suggested that increased neutrophil activity may be associated with a reduced risk of the disease, potentially due to their role in modulating inflammatory responses ^[42-43]^.

Similarly, in the African population, the TEMR analysis revealed notable associations such as lymphocyte count to schizophrenia (*OR*=0.01, *P<*0.001) which might suggest a substantial protective role of higher lymphocyte counts against schizophrenia, potentially through immune regulation mechanisms ^[44]^. The link between glucose levels and schizophrenia (*OR*=0.84, *P*<0.001) could reflect the metabolic disturbances that are often observed in patients with schizophrenia and might indicate a broader metabolic syndrome component of the disorder ^[45]^. The causal relationship between TC and schizophrenia (*OR*=0.78, *P*=0.007) in this demographic implies a protective effect of lower cholesterol levels, which contrasts with findings in the Asian population, suggesting the influence of genetic and environmental factors in different populations. Cholesterol is involved in the production of steroid hormones and neurosteroids, which neurosteroids have been found to modulate the central nervous system’s activity and may influence symptoms of schizophrenia. They can also affect the immune system, which has been implicated in the pathophysiology of schizophrenia ^[46-47]^. Higher TC levels could reflect more robust synthesis of such compounds, potentially contributing to more stable cellular functions and better disease outcomes ^[48]^. Additionally, the significant association between TC and hypertension (*OR*=0.74, *P*<0.001) could hint at the complex interplay between lipid metabolism and blood pressure regulation. There have been suggestions that certain lipid components might have a role in immune defense systems and that some aspects of the inflammatory response may be influenced by lipid levels ^[49]^. Cholesterol may also modulate cell membrane properties, affecting the reactivity of blood vessels and contributing to the regulation of blood pressure (BP) ^[50]^ (Table S13).

In the Hispanic/Latino population, the TEMR method unveiled notable significant causal relationships, indicative of unique pathophysiological pathways. The association of TG to schizophrenia (*OR*=0.80, *P*<0.001) and TC to schizophrenia (*OR*=0.61, *P*<0.001) also suggested a link between metabolic dysregulation and the development of schizophrenia. The significant causal associations between basophil count (*OR*<0.01, *P*<0.001) and platelet count (*OR*=0.06, *P*<0.001) emphasized the role of the immune system in schizophrenia ^[51]^. The association of LDL-cholesterol (LDL-C) with hypertension (*OR*=0.97, *P*<0.001) parallelled the findings of TC in the African population, adding to the evidence that lipid metabolism plays a complex role in BP regulation across different ethnicities. Finally, the associations between TG (*OR*=0.96, *P*<0.001) and ischemic stroke suggested a pathogenic role for blood components and lipid metabolism in vascular health. For TG, while high levels are typically considered a risk factor for atherosclerosis and thus ischemic strokes, However, very low TG levels might also not be ideal, as they can indicate an insufficient energy reserve for normal cellular functions, which could potentially affect overall health and cellular processes, including those in vascular cells ^[52]^ (Table S14).

## Discussion

In this paper, we propose a trans-ethnic MR method called TEMR to improve statistical power and estimation accuracy of MR in the target population only using trans-ethnic large-scale GWAS summary datasets. TEMR showed superior precision and power of causal effect estimation in the target population relative to other published MR methods in the simulation study. Leveraging the biobank-scale GWAS summary data from European, application of inferring causal relationships from 17 blood biomarkers to four diseases in East Asian, African population and Hispanic populations discover 13 new causal relationships that not found using published MR methods.

TEMR bridges the causal effects of multiple ethnics using a trans-ethnic genetic correlation coefficient. With the increase of trans-ethnic genetic association, the statistical power of causal effect in the non-European population is significantly improved. Trans-ethnic genetic correlation measures the extent to which genetic variants influence phenotypes similarly across different populations. With the advent of genomic technologies, researchers were able to conduct genome-wide studies of large cohorts from different ethnicities. These studies revealed that while there is substantial genetic variation between different populations, certain variants have similar frequencies and effects across groups. Numerous studies showed that the genetic variants for many traits were highly correlated across different populations. Trans-ethnic genetic correlation is assessed using various methods, such as multi-ancestry GWAS, TWAS and PRS prediction, etc. It can be estimated by abundant methods including LD score regression ^[53]^, HDL ^[54]^, GCTA-GREML ^[55]^, BOLT-REML ^[56]^ and PAINTOR ^[57]^, etc. They can achieve much higher accuracy than z-score based method. In this paper, TEMR uses a simple Z-score method to get results quickly, and using these methods will make TEMR perform better. TEMR is suitable for traits with high genetic association between different ethnics. When the genetic association between traits is nearly zero, TEMR method behaves similar to traditional MR Method.

There are several limitations in our study. The impact of pleiotropy is an important topic in MR study. Here we consider the case of no pleiotropy and horizontal pleiotropy. For the latter, we propose a two-step process to remove the pleiotropy effect from traditional Wald ratio using MR-Egger regression, and obtain the TEMR-Wald ratio estimation. The limitation of this process is that it also requires the InSIDE assumption and cannot remove the influence of correlated pleiotropy. Available solution is that detect outliers using published methods such as MR Radial and MR-PRESSO, etc, and then remove them before conducting TEMR. In addition, when there are multiple ethnics, TEMR can improve the statistical power of causal effect estimation only in one target population leveraging other target populations and European population. In the future, we will extend TEMR to implement that improving the statistical power of causal effect estimation in multiple target population leveraging only European population. The degree of improvement in statistical power is closely related to the number of IVs and the magnitude of trans-ethnic genetic correlations.

In conclusion, we proposed a new method TEMR to improve statistical power and estimation accuracy of MR in the target population only using trans-ethnic large-scale GWAS summary dataset. It has important guiding significance for the discovery of new disease-related factors.

## Methods

### TEMR model based on two ancestries

Consider a target dataset {*G*_1_, *X*_1_,*Y*_1_} from an under-represented ancestry (e.g. East Asian ancestry) with small sample size, where *G*_1_ is an *N*_1_×*p* genotype matrix, *X*_1_ and *Y*_1_ are *N*_1_×1 phenotype/disease vectors, represent the exposure and outcome, respectively. Now we suppose a biobank-scale dataset {*G*_2_, *X*_2_,*Y*_2_} (e.g. European ancestry) is also available, where *G*_2_ is an *N*_2_× *p* genotype matrix, *X*_2_ and *Y*_2_ are *N*_2_×1 phenotype/disease vectors, represent exposure and outcome, respectively. We assume *N*_2_ □ *N*_1_. Since we are mainly interested in improving the statistical power of causal effect estimation in the target population leveraging biobank scale datasets from another ancestry. We choose the *p* independent IVs (SNPs) which are associated with at least one of exposures in two ancestries (*X*_1_ and *X*_2_). We can obtain the summary-level data of *p* SNPs from published GWAS studies, including the beta-coefficients (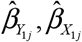 and 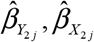) and their standard error (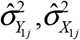 and 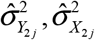).

When the three core assumptions of MR are all satisfied, we can obtain the causal effect estimation using the Wald ratio for each SNP

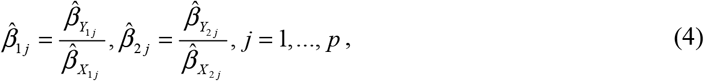

with their variances

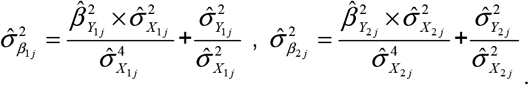

The 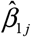 and 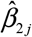 are the causal effect estimation of exposure on outcome using *j* -th SNPs in the target and auxiliary populations, respectively. We set up the following multivariable normal distribution model for Wald ratios from two populations

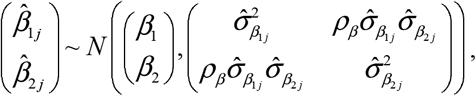

where *β*_1_ and *β*_2_ are the causal effect of exposure on outcome in the target and auxiliary populations, respectively. They can be the same or different. *ρ*_*β*_ is the trans-ethnic genetic correlation, which represents the correlation of the causal effects of one exposure on one outcome in two ancestries (e.g. Chinese and European), and it can be calculated by the Pearson correlation

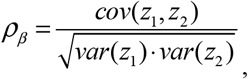

where *z*_1_ and *z*_2_ are the *z*-scores of *p*-dimensional Wald ratio vectors in two ancestries, respectively

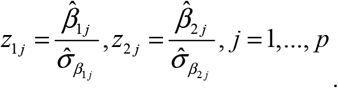

We aim to improve the statistical power of causal effect (*β*_1_) estimation in the target population using the trans-ethnic genetic correlation *ρ*_*β*_, which connect the causal effects of two ethnics. Based on the model (1) and the conditional normal distribution formula ^[58]^, we have

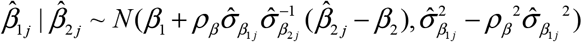

with its variance

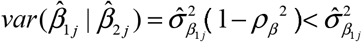

Therefore, the variance of *j*-th Wald ratio estimation 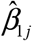 conditional on 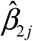 is smaller than its original variance as the trans-ethnic genetic correlation *ρ*_*β*_ increasing. Then we obtain the conditional log-likelihood function of model (2)

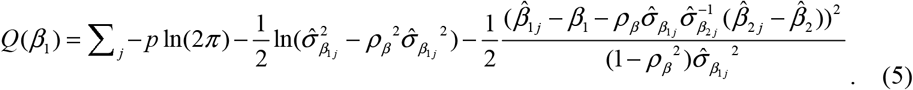

where 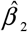 is obtained by IVW or other effective MR methods using large-scale dataset in the auxiliary population. We aim to maximize the log conditional likelihood function using Nelder-Mead method ^[38]^ to obtain the estimation of *β*_1_. Then we use Likelihood-ratio test to perform hypothesis testing,

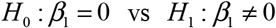

the testing statistics is

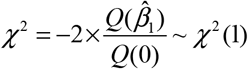

When there is horizontal pleiotropy, the third assumption of MR is violated, the causal effect estimation using the traditional Wald ratio is biased and we model the TEMR-Wald ratio as following

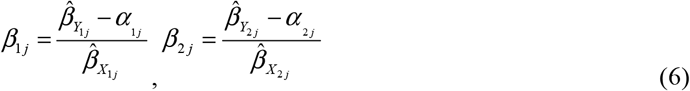

where *α*_1 *j*_ represent the horizontal pleiotropy and it is unknown. Therefore, in the first step, we need to estimate *α*_1 *j*_ and *α*_2 *j*_ using MR-Egger regression

1. seperately estimate causal effect 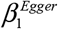 and 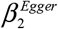 in each ancestry using MR-Egger regression

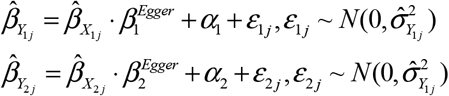
2. seperately estimate horizontal pleiotropy *α*_1*j*_ and *α*_2*j*_ in each ancestry using

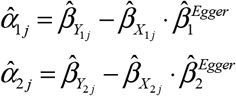

Then we can obtain the estimations of new Wald ratio 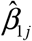 and 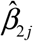 by substituting 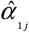 and 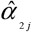 into equation (6). Following we use models (1,2,4,5) to obtain the estimation of *β*_1_. The difference is that the 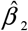 in model (5) is obtained by horizontal pleiotropy-robust MR methods using large-scale dataset in the auxiliary population.

### TEMR model based on multiple ancestries

If there are *E>* 2 ancestries, the target dataset is {*G*_*T*_, *X*_*T*_,*Y*_*T*_ } and the auxiliary datasets are {*G*_*a*_, *X*_*a*_,*Y*_*a*_ }(*a* = 2,…, *E*), we set up the following multivariable normal distribution model using Wald ratios from *E* ancestries

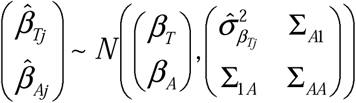

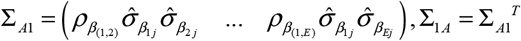 and 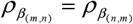. The conditional distribution of 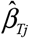 given 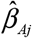 is

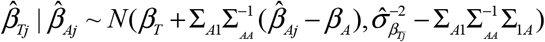

Then we obtain the estimation of *β*_*T*_ by Maximum Likelihood Estimation using Nelder-Mead method.

Due to the predominant representation of European individuals in public GWAS summary dataset, with smaller sample sizes for other ethnicities, our aim is to utilize the information from the European population to improve the causal effect estimation precision and testing efficacy for smaller sample populations. Furthermore, if our focus is exclusively on the Asian population, the inclusion of other small-sample ethnicities could still contribute to enhancing the estimation performance of causal effects on the Asian population, although the contribution may not be as substantial as that from the European population.

### Simulation settings

In our simulation study, we systematically evaluated the performance of TEMR through several key steps. Initially, we generated individual data for exposure (*X*_*e*_), outcome (*Y*_*e*_) and genotypes (*G*_*je*_, *j* =1,…, *p*) in multiple ethnicities (*e* =1,…,*E*):

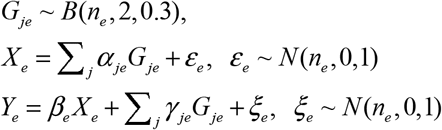

Subsequently, we obtain GWAS summary data (including the regression coefficients (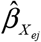 and 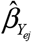) and their standard errors 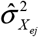 and 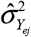)) by linear regressions of continuous variables on each SNP and logistic regressions of binary variables on each SNP, enabling the calculation of the Wald ratios’ standard errors for each SNP:

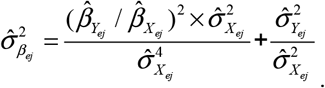

The reason for initially generating individual-level data was to simulate the variation in the estimates and precision of the Wald ratio obtained with different sample sizes in real-world applications. While it is possible to directly simulate based on the observed Wald ratio from practical data, the limitation lies in the finite range of sample sizes in public GWAS summary datasets. This constraint prevents a comprehensive simulation of the performance of TEMR under various sample size scenarios. Next, we generate the Wald ratios (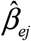) for different ethnicities using trans-ethnic genetic correlation 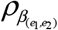:

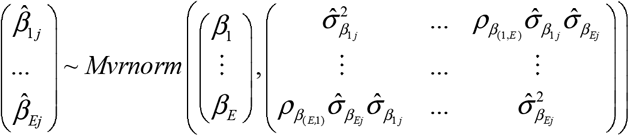

We considered scenarios where the causal effects are the same or different across different ethnicities, as well as situations where the causal effects are either zero (*β*_*e*_ = 0) or non-zero (*β*_*e*_ = 0.05, 0.1, 0.15, 0.2). We also explore various scenarios, including different trans-ethnic genetic correlation coefficients between ethnicities. We considered the number of ethnicities is *E*=2 or *E*=4, 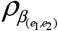 vary 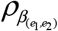 with 0.1-0.9 as well as consider the trans-ethnic genetic correlations are the same or different across different race-pairs. Acknowledging the potential influence of genetic factors across diverse racial backgrounds, this exploration aimed to account for variations in genetic correlation. Furthermore, in an effort to optimize precision and statistical power, we systematically varied the number of SNPs (*p*=25, 50, 100 and 200) while keeping other parameters constant. This process allows us to determine how much the precision and statistical power of causal effect estimates can be significantly improved under different numbers of IVs. Finally, our simulation study was designed to encompass three distinct scenarios: one where pleiotropy was absent (*γ*_*je*_ = 0), another where balanced horizontal pleiotropy was present(*γ*_*je*_ ∼ *U* (−0.2, 0.2)), and a third scenario where directional horizontal pleiotropy was present (*γ*_*je*_ ∼ *U* (0, 0.2)). We then applied our method TEMR to estimate causal effects in the target populations. To benchmark the performance of our approach, we conducted a comparative analysis with previously published MR methods ^[2]^ based on the Wald ratio, including IVW method ^[59]^, MR-Egger ^[60]^, Simple Median ^[61]^, Weighted Median ^[62]^, Simple Mode ^[63]^, Weighted Mode ^[64]^. By thoroughly examining these scenarios, we aimed to provide a comprehensive assessment of TEMR’s performance and robustness under diverse genetic and phenotypic conditions.

The evaluation metrics include estimation bias, standard error, type I error for testing null causal effect and statistical power for testing non-null causal effect. We utilized boxplots to demonstrate the results of bias and standard error, Q-Q plots to showcase the results of Type I error, and bar charts to depict the results of statistical power.

### Application

We applied TEMR to estimate the causal effects between different biomarkers and four diseases (hypertension, ischemic stroke, T2D and schizophrenia) in the East Aisan, African, Hispanic/Latino population, leveraging data from large European cohorts. These diseases were chosen for their significant public health impact, high prevalence, and representative nature of the complex interplay between genetics and environment. The GWAS summary data for the Asian population was mainly sourced from the BBJ with a sample size of 170,000. For the African population, the data was mainly obtained from the Pan-UKB with a sample size of 6,000, and for the Hispanic population from the GWAS Catalog with a sample size of. The GWAS summary data of European population was derived from the UKB with a sample size of 500,000. Details of datasets information are shown in Supplementary Table S7. Firstly, for each trait, we chose SNPs based on the criterion of *P*-value less than 5 × 10^−8^ in at least one ethnicity and no linkage disequilibrium (*r*^2^<0.001). The SNP satisfying above criterion in at least one of the ethnics are selected as IVs. Then, we applied TEMR and other six MR methods for trans-ethnic MR analysis using the 17 biomarkers and four diseases. For each target population, we use the other three datasets as auxiliary datasets.

## Data Availability

The GWAS summary data in UK Biobank are publicly available at http://www.nealelab.is/uk-biobank. The GWAS summary data in Biobank Japan are publicly available at https://pheweb.jp/. The GWAS summary data in Pan-UKB are publicly available at https://pan.ukbb.broadinstitute.org/. Other GWAS summary data are publicly available at IEU OpenGWAS project (https://gwas.mrcieu.ac.uk/) and GWAS Catalog (https://www.ebi.ac.uk/gwas/). All the analysis in our article were implemented by R software (version 4.3.2). R packages used in our analysis include TwoSampleMR, MendelianRandomization, ggplot2, plinkbinr and ieugwasr. TEMR package can be implemented by https://github.com/hhoulei/TEMR. All the codes for simulation are uploaded in https://github.com/hhoulei/TEMR_Simul.

## Acknowledgements

We thank Haoran Xue for his constructive suggestions.

## Author Contributions

HL and FX conceived the study. LH contributed to theoretical derivation with assistance from ZY. SW contributed to the data simulation. LH and SW contributed to the application. LH and SW wrote the manuscript with input from all other authors. All authors reviewed and approved the final manuscript.

## Data and code availability

The GWAS summary data in UK Biobank are publicly available at http://www.nealelab.is/uk-biobank. The GWAS summary data in Biobank Japan are publicly available at https://pheweb.jp/. The GWAS summary data in Pan-UKB are publicly available at https://pan.ukbb.broadinstitute.org/. Other GWAS summary data are publicly available at IEU OpenGWAS project (https://gwas.mrcieu.ac.uk/) and GWAS Catalog (https://www.ebi.ac.uk/gwas/). All the analysis in our article were implemented by R software (version 4.3.2). R packages used in our analysis include *TwoSampleMR, MendelianRandomization, ggplot*2, *plinkbinr* and *ieugwasr*. TEMR package can be implemented by https://github.com/hhoulei/TEMR. All the codes for simulation are uploaded in https://github.com/hhoulei/TEMR_Simul.

## Ethics approval and consent to participate

The data used in our study was all publicly available and obtained written informed consent from all participants.

## Supplementary File

### Supplementary Materials and Methods

Details of methods and results of simulation.

### Supplementary Table

GWAS summary datasets information and results of application.

