## Supplementary Materials and Methods for "TEMR: Trans-ethnic Mendelian Randomization Method using Large-scale GWAS Summary Datasets"

### Estimation of causal effect in target population

When there is no pleiotropy, the conditional distribution of causal effect in target population given that in auxiliary dataset is

$$\hat{\beta}_{1j} \mid \hat{\beta}_{2j} \sim N(\beta_1 + \rho_\beta \sigma_{\beta_{1j}} \sigma_{\beta_{2j}}^{-1} (\hat{\beta}_{2j} - \beta_2), \sigma_{\beta_{1j}}^2 - \rho_\beta{}^2 \sigma_{\beta_{1j}}{}^2)$$

The conditional likelihood function is

$$p = \prod_j \frac{1}{(2\pi)\sqrt{\sigma_{\beta_{1j}}^2 - \rho_\beta{}^2 \sigma_{\beta_{1j}}{}^2}} \exp\{-\frac{1}{2} \frac{(\hat{\beta}_{1j} - \beta_1 - \rho_\beta \sigma_{\beta_{1j}} \sigma_{\beta_{2j}}^{-1} (\hat{\beta}_{2j} - \beta_2))^2}{\sigma_{\beta_{1j}}^2 - \rho_\beta{}^2 \sigma_{\beta_{1j}}{}^2}\}$$

The log-likelihood function is

$$Q = \ln p$$

$$= \sum_j -p\ln(2\pi) - \frac{1}{2}\ln(\sigma_{\beta_{1j}}^2 - \rho_\beta{}^2 \sigma_{\beta_{1j}}{}^2) - \frac{1}{2} \frac{(\hat{\beta}_{1j} - \beta_1 - \rho_\beta \sigma_{\beta_{1j}} \sigma_{\beta_{2j}}^{-1} (\hat{\beta}_{2j} - \beta_2))^2}{(1 - \rho_\beta{}^2) \sigma_{\beta_{1j}}{}^2}$$

Then we obtain the derivation of $Q$:

$$\frac{\partial Q}{\partial \beta_1} = -\frac{1}{2} \sum_j \frac{-2(\hat{\beta}_{1j} - \beta_1 - \rho_\beta \sigma_{\beta_{1j}} \sigma_{\beta_{2j}}^{-1} (\hat{\beta}_{2j} - \beta_2))}{(1 - \rho_\beta{}^2) \sigma_{\beta_{1j}}{}^2} = 0$$

Finally, we obtain the *i*-th iteration of $\beta_1$

$$\sum_j \sigma_{\beta_{1j}}{}^{-2} (\hat{\beta}_{1j} - \rho_\beta \sigma_{\beta_{1j}} \sigma_{\beta_{2j}}^{-1} (\hat{\beta}_{2j} - \beta_2)) - \beta_1 \sum_j \sigma_{\beta_{1j}}{}^{-2} = 0$$

$$\beta_1 = \frac{\sum_j \sigma_{\beta_{1j}}{}^{-2} (\hat{\beta}_{1j} - \rho_\beta \sigma_{\beta_{1j}} \sigma_{\beta_{2j}}^{-1} (\hat{\beta}_{2j} - \beta_2))}{\sum_j \sigma_{\beta_{1j}}{}^{-2}}$$

#### *Multiple ancestries*

The conditional distribution of $\hat{\beta}_{Tj}$ given $\hat{\beta}_{Aj}$ is

$$\hat{\beta}_{Tj} \mid \hat{\beta}_{Aj} \sim N(\beta_T + \Sigma_{A1} \Sigma_{AA}^{-1} (\hat{\beta}_{Aj} - \beta_A), \sigma_{\beta_{Tj}}^{-2} - \Sigma_{A1} \Sigma_{AA}^{-1} \Sigma_{1A})$$

where $\hat{\beta}_{Aj} = \begin{pmatrix} \hat{\beta}_{2j} \\ \ldots \\ \hat{\beta}_{Ej} \end{pmatrix}, \beta_A = \begin{pmatrix} \beta_2 \\ \ldots \\ \beta_E \end{pmatrix}, \Sigma_{AA} = \begin{pmatrix} \sigma_{\beta_{2j}}^2 & \ldots & \rho_{\beta_{(2,E)}} \sigma_{\beta_{2j}} \sigma_{\beta_{Ej}} \\ \ldots & \ldots & \ldots \\ \rho_{\beta_{(E,2)}} \sigma_{\beta_{Ej}} \sigma_{\beta_{2j}} & \ldots & \sigma_{\beta_{Ej}}^2 \end{pmatrix}$

and $\Sigma_{A1} = \begin{pmatrix} \rho_{\beta_{(1,2)}} \sigma_{\beta_{1j}} \sigma_{\beta_{2j}} & \ldots & \rho_{\beta_{(1,E)}} \sigma_{\beta_{1j}} \sigma_{\beta_{Ej}} \end{pmatrix}, \Sigma_{1A} = \Sigma_{A1}{}^T$, $\rho_{\beta_{(m,n)}} = \rho_{\beta_{(n,m)}}$.

The conditional likelihood function is

$$p = \prod_j (2\pi)^{-1/2} \,|\, \sigma_{\beta_{Tj}}^{-2} - \Sigma_{A1}\Sigma_{AA}^{-1}\Sigma_{1A} \,|^{-1/2}$$
$$\exp\{-\frac{1}{2}(\hat{\beta}_{Tj} - \beta_T - \Sigma_{A1}\Sigma_{AA}^{-1}(\hat{\beta}_{Aj} - \beta_A))^T$$
$$(\sigma_{\beta_{Tj}}^{-2} - \Sigma_{A1}\Sigma_{AA}^{-1}\Sigma_{1A})^{-1}(\hat{\beta}_{Tj} - \beta_T - \Sigma_{A1}\Sigma_{AA}^{-1}(\hat{\beta}_{Aj} - \beta_A))\}$$

The log-likelihood function is

$$Q = \ln p$$
$$= \sum_j -p\ln(2\pi) - \frac{1}{2}\ln |\, \sigma_{\beta_{Tj}}^{-2} - \Sigma_{A1}\Sigma_{AA}^{-1}\Sigma_{1A} \,|$$
$$-\frac{1}{2}\{(\hat{\beta}_{Tj} - \beta_T - \Sigma_{A1}\Sigma_{AA}^{-1}(\hat{\beta}_{Aj} - \beta_A))^T(\sigma_{\beta_{Tj}}^{-2} - \Sigma_{A1}\Sigma_{AA}^{-1}\Sigma_{1A})^{-1}$$
$$(\hat{\beta}_{Tj} - \beta_T - \Sigma_{A1}\Sigma_{AA}^{-1}(\hat{\beta}_{Aj} - \beta_A))\}$$
$$= \sum_j -p\ln(2\pi) - \frac{1}{2}\ln |\, \sigma_{\beta_{Tj}}^{-2} - \Sigma_{A1}\Sigma_{AA}^{-1}\Sigma_{1A} \,|$$
$$-\frac{1}{2}\{(\hat{\beta}_{Tj} - \beta_T - \begin{pmatrix} \rho_{\beta_{(1,2)}}\sigma_{\beta_{1j}}\sigma_{\beta_{2j}} & \cdots & \rho_{\beta_{(1,E)}}\sigma_{\beta_{1j}}\sigma_{\beta_{Ej}} \end{pmatrix} \begin{pmatrix} \sigma_{\beta_{2j}}^2 & \cdots & \rho_{\beta_{(2,E)}}\sigma_{\beta_{2j}}\sigma_{\beta_{Ej}} \\ \cdots & \cdots & \cdots \\ \rho_{\beta_{(E,2)}}\sigma_{\beta_{Ej}}\sigma_{\beta_{2j}} & \cdots & \sigma_{\beta_{Ej}}^2 \end{pmatrix}^{-1} (\hat{\beta}_{Aj} - \beta_A))^T$$
$$(\sigma_{\beta_{Tj}}^{-2} - \begin{pmatrix} \rho_{\beta_{(1,2)}}\sigma_{\beta_{1j}}\sigma_{\beta_{2j}} & \cdots & \rho_{\beta_{(1,E)}}\sigma_{\beta_{1j}}\sigma_{\beta_{Ej}} \end{pmatrix} \begin{pmatrix} \sigma_{\beta_{2j}}^2 & \cdots & \rho_{\beta_{(2,E)}}\sigma_{\beta_{2j}}\sigma_{\beta_{Ej}} \\ \cdots & \cdots & \cdots \\ \rho_{\beta_{(E,2)}}\sigma_{\beta_{Ej}}\sigma_{\beta_{2j}} & \cdots & \sigma_{\beta_{Ej}}^2 \end{pmatrix} \begin{pmatrix} \rho_{\beta_{(1,2)}}\sigma_{\beta_{1j}}\sigma_{\beta_{2j}} \\ \cdots \\ \rho_{\beta_{(1,E)}}\sigma_{\beta_{1j}}\sigma_{\beta_{Ej}} \end{pmatrix})^{-1}$$
$$(\hat{\beta}_{Tj} - \beta_T - \begin{pmatrix} \rho_{\beta_{(1,2)}}\sigma_{\beta_{1j}}\sigma_{\beta_{2j}} & \cdots & \rho_{\beta_{(1,E)}}\sigma_{\beta_{1j}}\sigma_{\beta_{Ej}} \end{pmatrix} \begin{pmatrix} \sigma_{\beta_{2j}}^2 & \cdots & \rho_{\beta_{(2,E)}}\sigma_{\beta_{2j}}\sigma_{\beta_{Ej}} \\ \cdots & \cdots & \cdots \\ \rho_{\beta_{(E,2)}}\sigma_{\beta_{Ej}}\sigma_{\beta_{2j}} & \cdots & \sigma_{\beta_{Ej}}^2 \end{pmatrix}^{-1} (\hat{\beta}_{Aj} - \beta_A))^T\}$$

Then we obtain the derivation of $Q$:

$$\frac{\partial Q}{\partial \beta_T} = -\frac{1}{2}\sum_j \frac{-2(\hat{\beta}_{Tj} - \beta_T - \Sigma_{A1}\Sigma_{AA}^{-1}(\hat{\beta}_{Aj} - \beta_A))}{\sigma_{\beta_{Tj}}^{-2} - \Sigma_{A1}\Sigma_{AA}^{-1}\Sigma_{1A}} = 0$$

Finally, we obtain the *i*-th iteration of $\beta_1$

$$\sum_j (\sigma_{\beta_{Tj}}^{-2} - \Sigma_{A1}\Sigma_{AA}^{-1}\Sigma_{1A})(\hat{\beta}_{Tj} - \Sigma_{A1}\Sigma_{AA}^{-1}(\hat{\beta}_{Aj} - \beta_A))$$
$$-\beta_T \sum_j \sigma_{\beta_{Tj}}^{-2} - \Sigma_{A1}\Sigma_{AA}^{-1}\Sigma_{1A} = 0$$
$$\beta_T = \frac{\sum_j (\sigma_{\beta_{Tj}}^{-2} - \Sigma_{A1}\Sigma_{AA}^{-1}\Sigma_{1A})(\hat{\beta}_{Tj} - \Sigma_{A1}\Sigma_{AA}^{-1}(\hat{\beta}_{Aj} - \beta_A))}{\sum_j \sigma_{\beta_{Tj}}^{-2} - \Sigma_{A1}\Sigma_{AA}^{-1}\Sigma_{1A}}$$

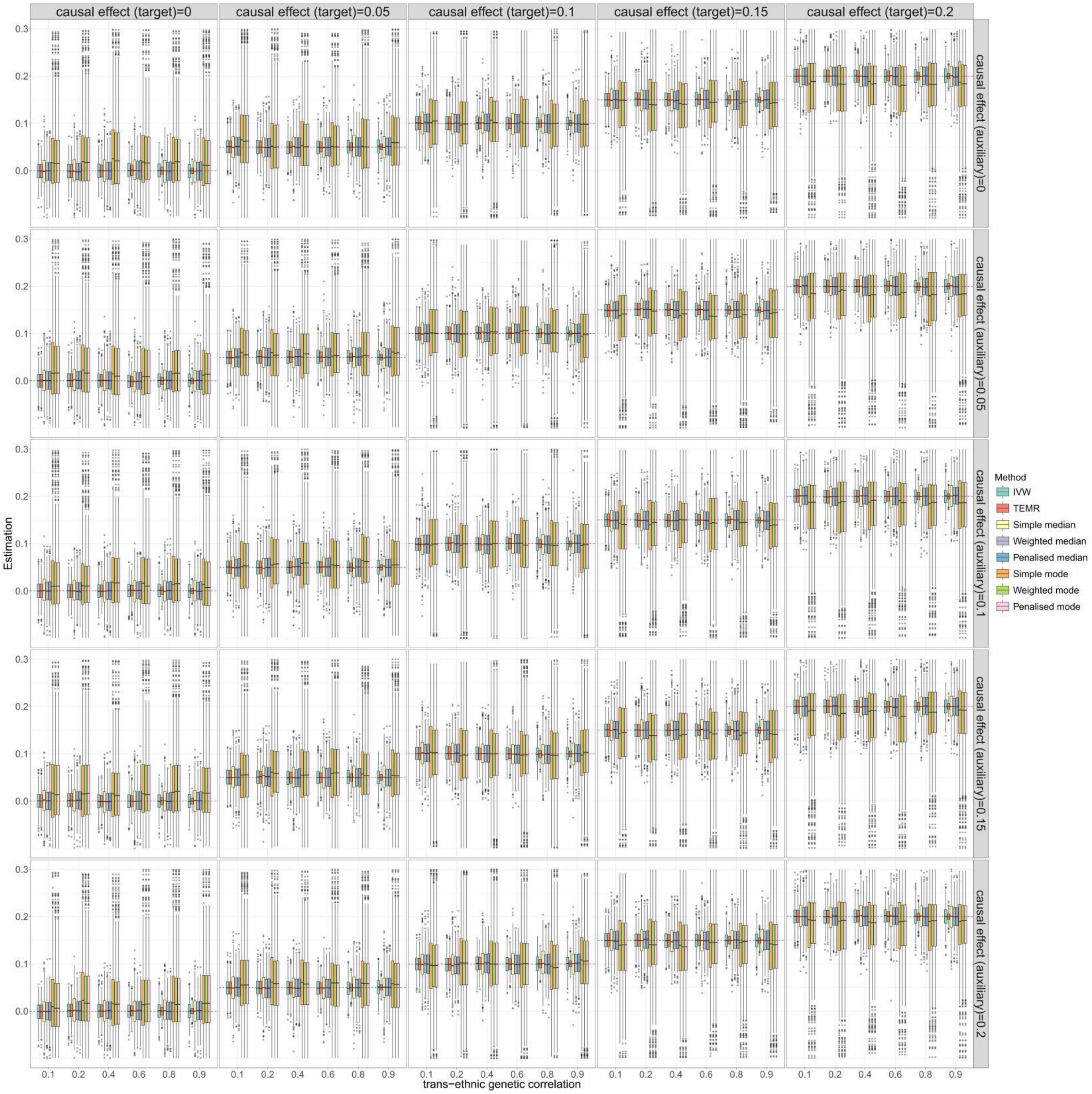


**Figure S1. Boxplots of simulation results with continuous outcome for causal effect estimation in the target population when there is one auxiliary population (no horizontal pleiotropy).** Sample size of target population is 3,000 and the sample size of auxiliary population is 300,000. IVs include 100 common SNPs. Boxplots show the performances of causal effect estimation in target population when the causal effect of target/auxiliary population is 0 to 0.2, respectively. IVW, Inverse-variance weighted method.

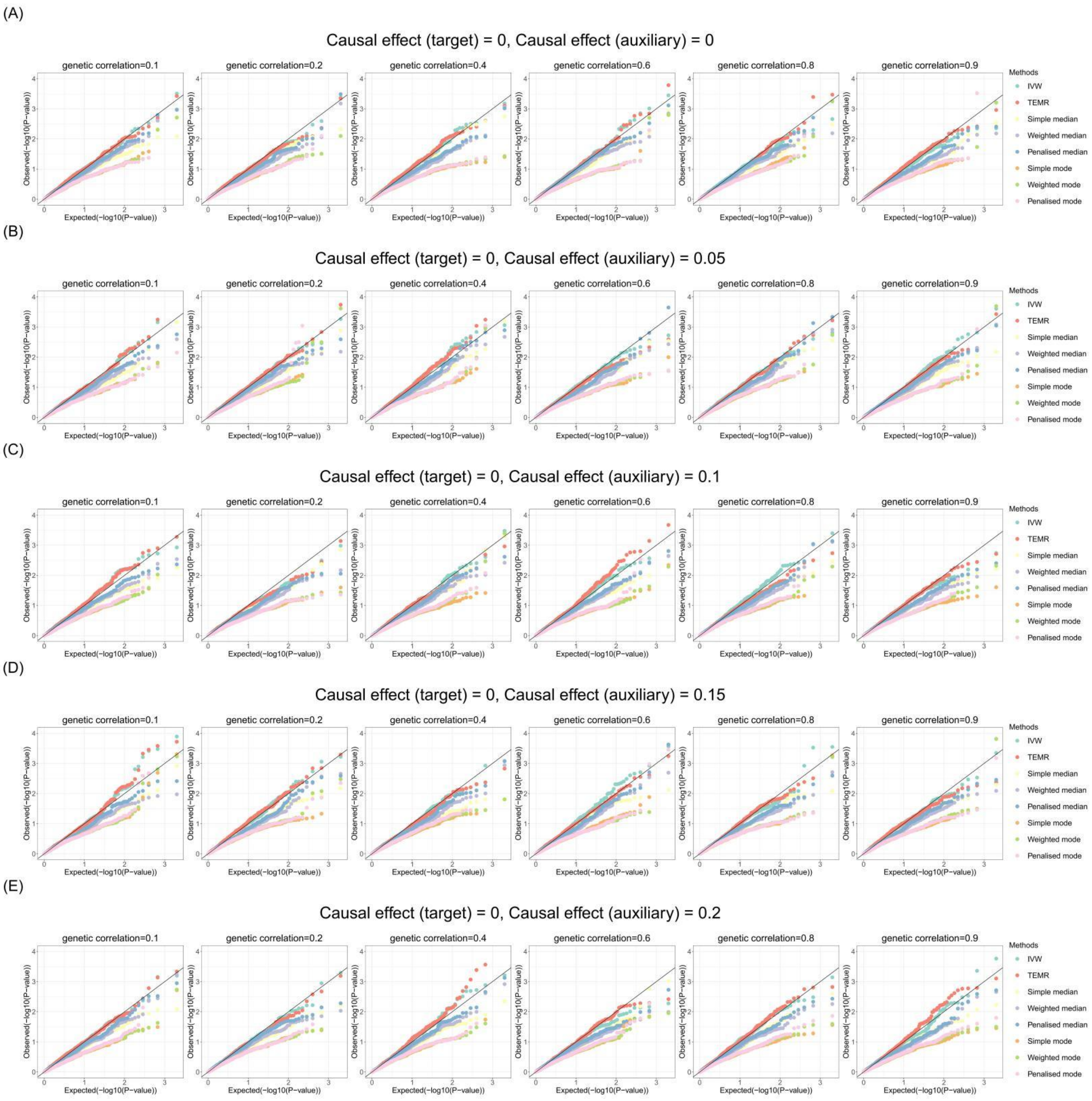


**Figure S2. Q-Q plots of simulation results with continuous outcome for causal effect estimation in the target population when there is one auxiliary population (no horizontal pleiotropy).** Sample size of target population is 3,000 and the sample size of auxiliary population is 300,000. IVs include 100 common SNPs. Q-Q plots show the performances of Type I error rates of zero causal effect estimation in target population when the causal effect of auxiliary population is 0 to 0.2, respectively. IVW, Inverse-variance weighted method.

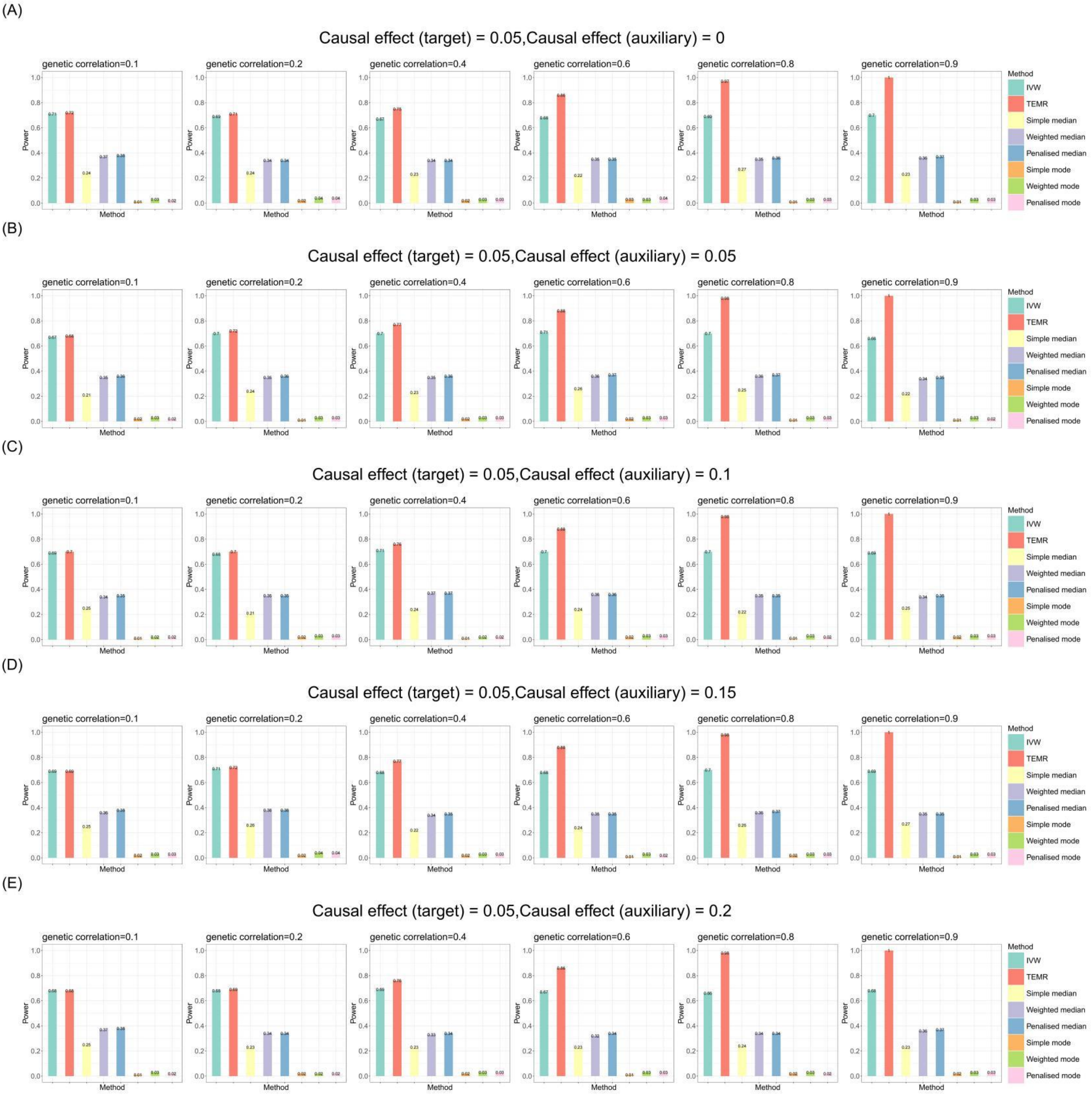


**Figure S3. Bar chart plots of simulation results with continuous outcome for causal effect estimation in the target population when there is one auxiliary population (no horizontal pleiotropy).**

Sample size of target population is 3,000 and the sample size of auxiliary population is 300,000. IVs include 100 common SNPs. Bar chart plots illustrate the statistical power performance when the causal effect estimation is 0.05 in the target population, and from 0 to 0.2 in the auxiliary population, respectively. IVW, Inverse-variance weighted method.

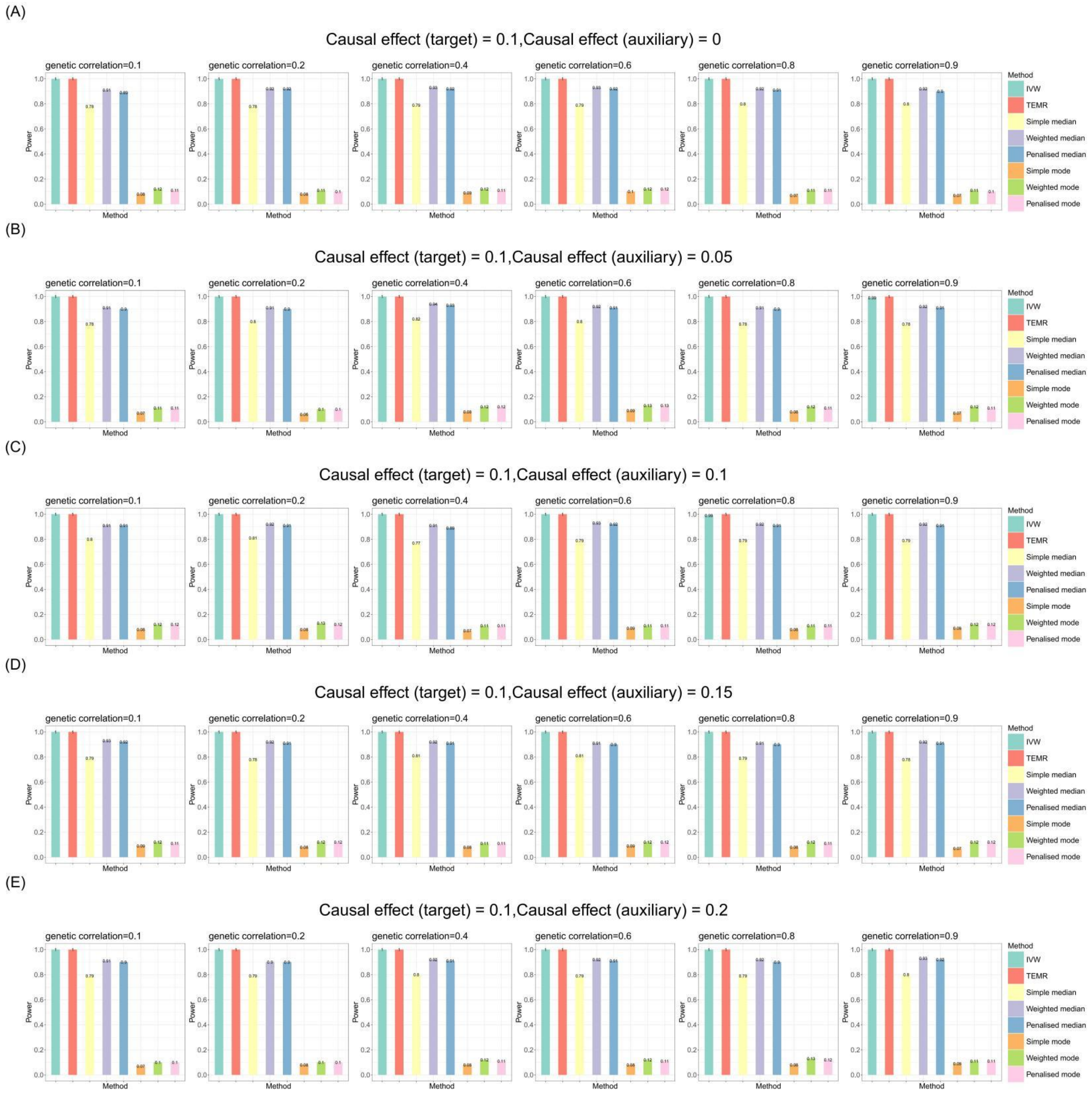


**Figure S4. Bar chart plots of simulation results with continuous outcome for causal effect estimation in the target population when there is one auxiliary population (no horizontal pleiotropy).**

Sample size of target population is 3,000 and the sample size of auxiliary population is 300,000. IVs include 100 common SNPs. Bar chart plots illustrate the statistical power performance when the causal effect estimation is 0.1 in the target population, and from 0 to 0.2 in the auxiliary population, respectively. IVW, Inverse-variance weighted method.

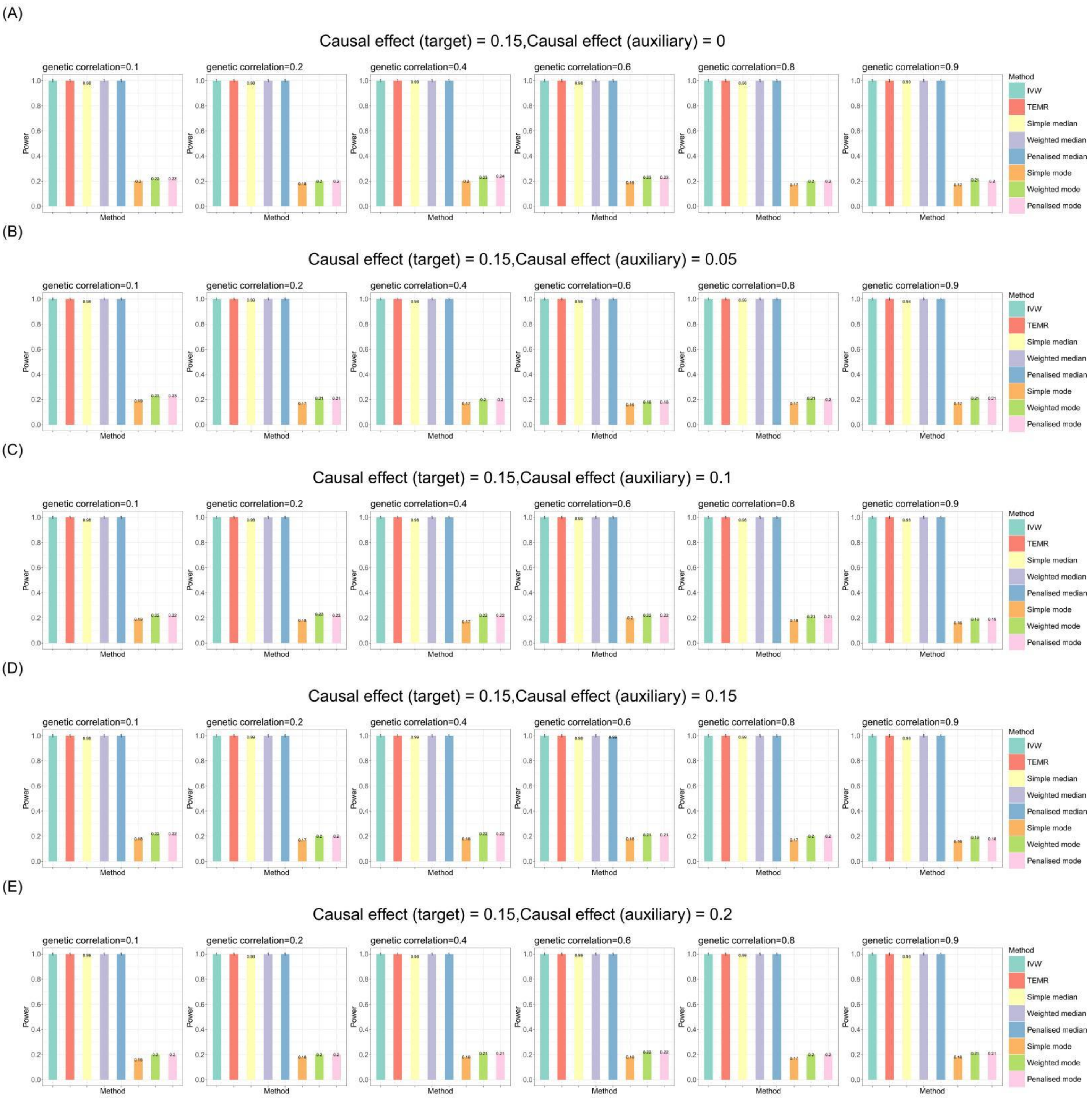


**Figure S5. Bar chart plots of simulation results with continuous outcome for causal effect estimation in the target population when there is one auxiliary population (no horizontal pleiotropy).**

Sample size of target population is 3,000 and the sample size of auxiliary population is 300,000. IVs include 100 common SNPs. Bar chart plots illustrate the statistical power performance when the causal effect estimation is 0.15 in the target population, and from 0 to 0.2 in the auxiliary population, respectively. IVW, Inverse-variance weighted method.

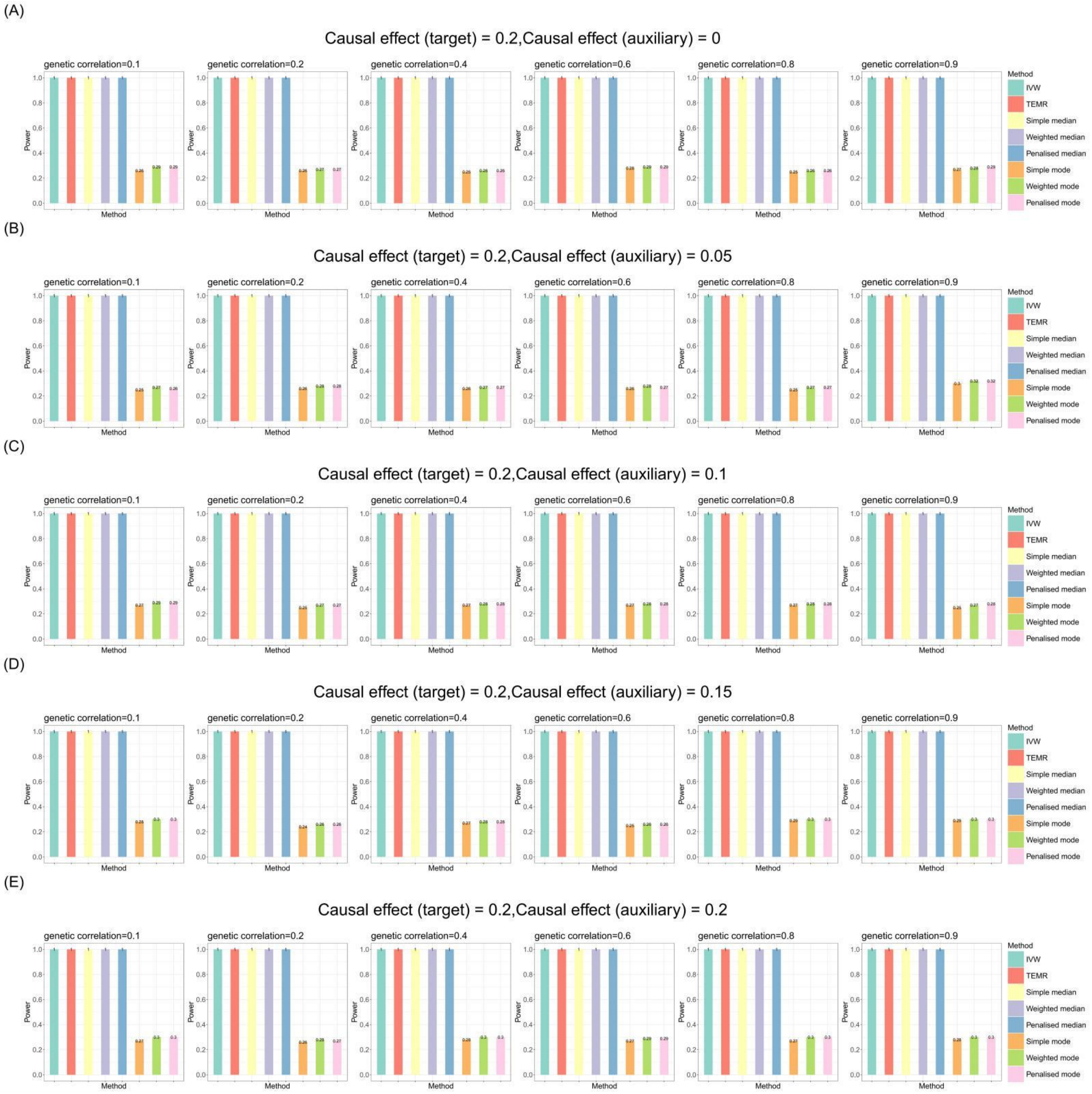


**Figure S6. Bar chart plots of simulation results with continuous outcome for causal effect estimation in the target population when there is one auxiliary population (no horizontal pleiotropy).**

Sample size of target population is 3,000 and the sample size of auxiliary population is 300,000. IVs include 100 common SNPs. Bar chart plots illustrate the statistical power performance when the causal effect estimation is 0.2 in the target population, and from 0 to 0.2 in the auxiliary population, respectively. IVW, Inverse-variance weighted method.

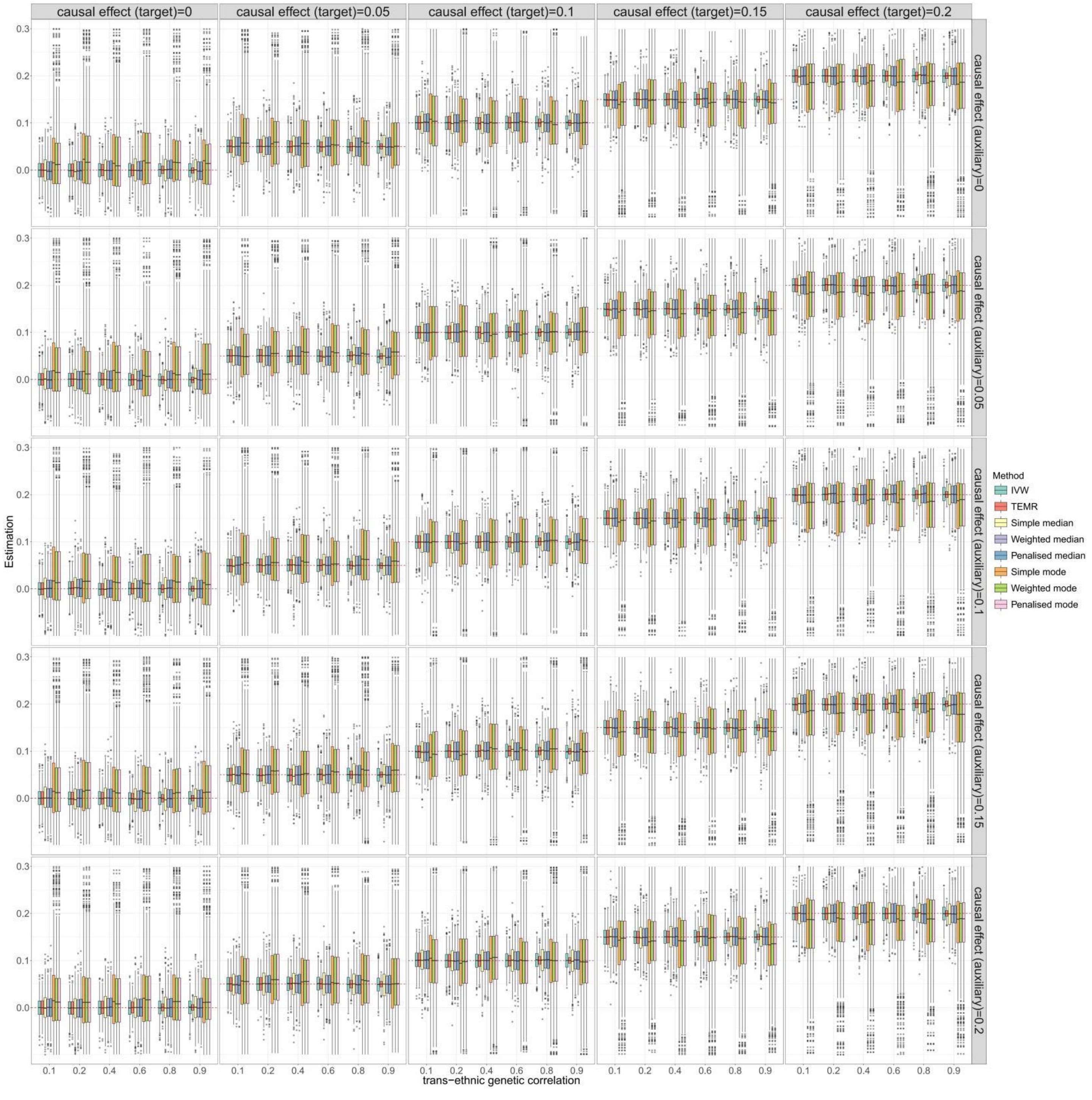


**Figure S7. Boxplots of simulation results with continuous outcome for causal effect estimation in the target population when there is one auxiliary population (balance horizontal pleiotropy).** Sample size of target population is 3,000 and the sample size of auxiliary population is 300,000. IVs include 100 common SNPs. Boxplots show the performances of causal effect estimation in target population when the causal effect of target/auxiliary population is 0 to 0.2, respectively. IVW, Inverse-variance weighted method.

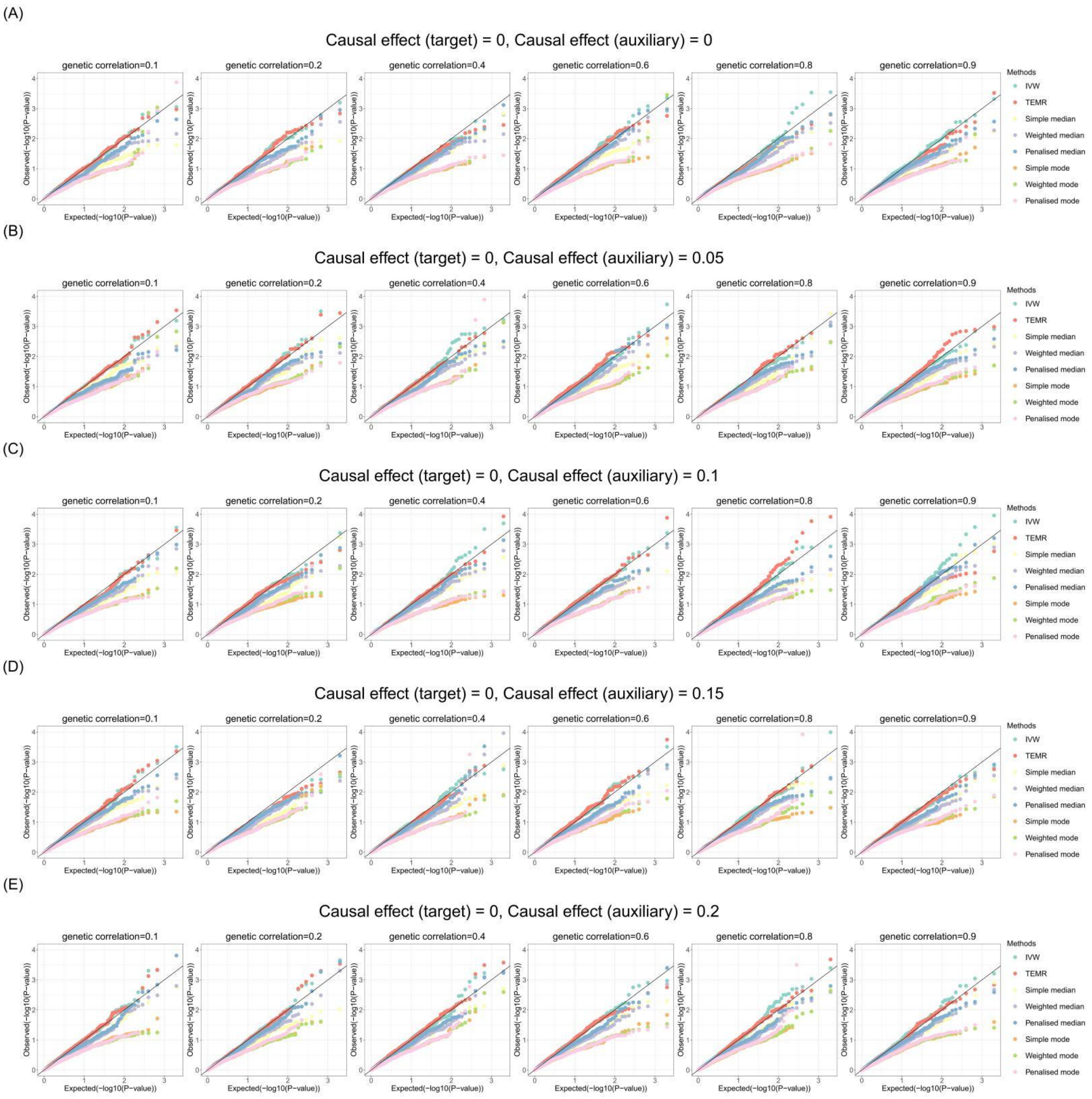


**Figure S8. Q-Q plots of simulation results with continuous outcome for causal effect estimation in the target population when there is one auxiliary population (balance horizontal pleiotropy).** Sample size of target population is 3,000 and the sample size of auxiliary population is 300,000. IVs include 100 common SNPs. Q-Q plots show the performances of Type I error rates of zero causal effect estimation in target population when the causal effect of auxiliary population is 0 to 0.2, respectively. IVW, Inverse-variance weighted method.

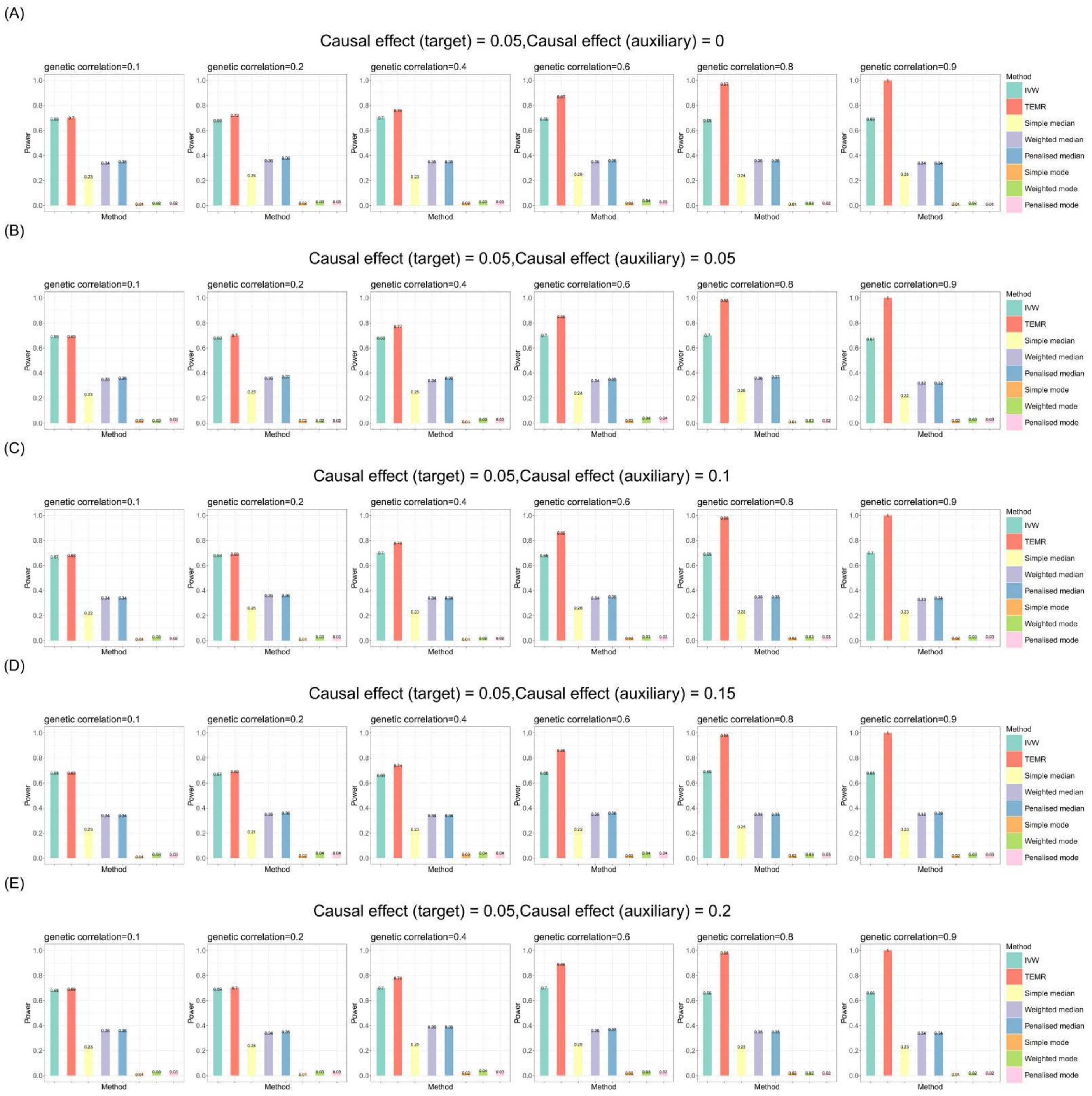


**Figure S9. Bar chart plots of simulation results with continuous outcome for causal effect estimation in the target population when there is one auxiliary population (balance horizontal pleiotropy).**

Sample size of target population is 3,000 and the sample size of auxiliary population is 300,000. IVs include 100 common SNPs. Bar chart plots illustrate the statistical power performance when the causal effect estimation is 0.05 in the target population, and from 0 to 0.2 in the auxiliary population, respectively. IVW, Inverse-variance weighted method.

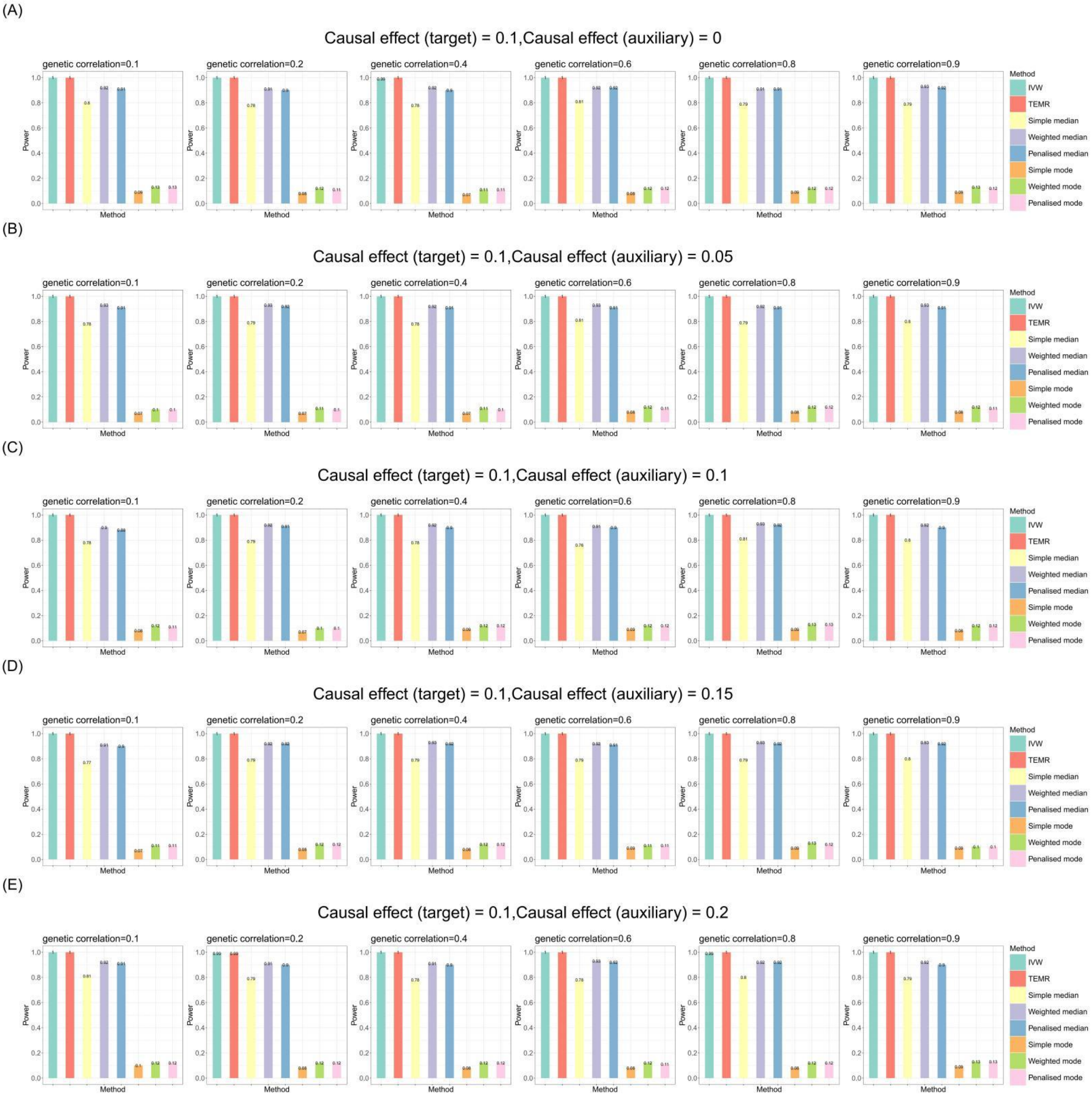


**Figure S10. Bar chart plots of simulation results with continuous outcome for causal effect estimation in the target population when there is one auxiliary population (balance horizontal pleiotropy).**

Sample size of target population is 3,000 and the sample size of auxiliary population is 300,000. IVs include 100 common SNPs. Bar chart plots illustrate the statistical power performance when the causal effect estimation is 0.1 in the target population, and from 0 to 0.2 in the auxiliary population, respectively. IVW, Inverse-variance weighted method.

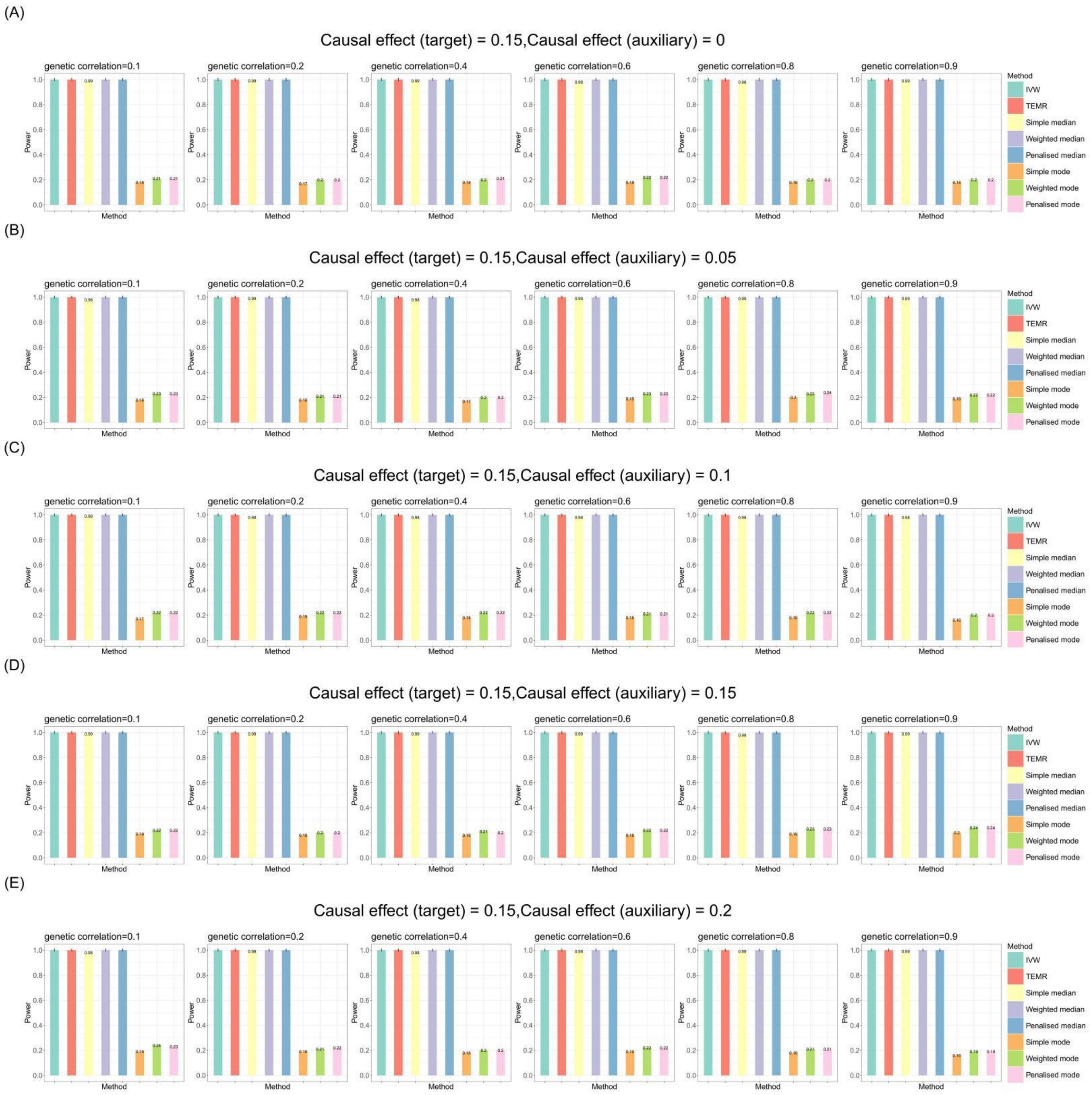


**Figure S11. Bar chart plots of simulation results with continuous outcome for causal effect estimation in the target population when there is one auxiliary population (balance horizontal pleiotropy).**

Sample size of target population is 3,000 and the sample size of auxiliary population is 300,000. IVs include 100 common SNPs. Bar chart plots illustrate the statistical power performance when the causal effect estimation is 0.15 in the target population, and from 0 to 0.2 in the auxiliary population, respectively. IVW, Inverse-variance weighted method.

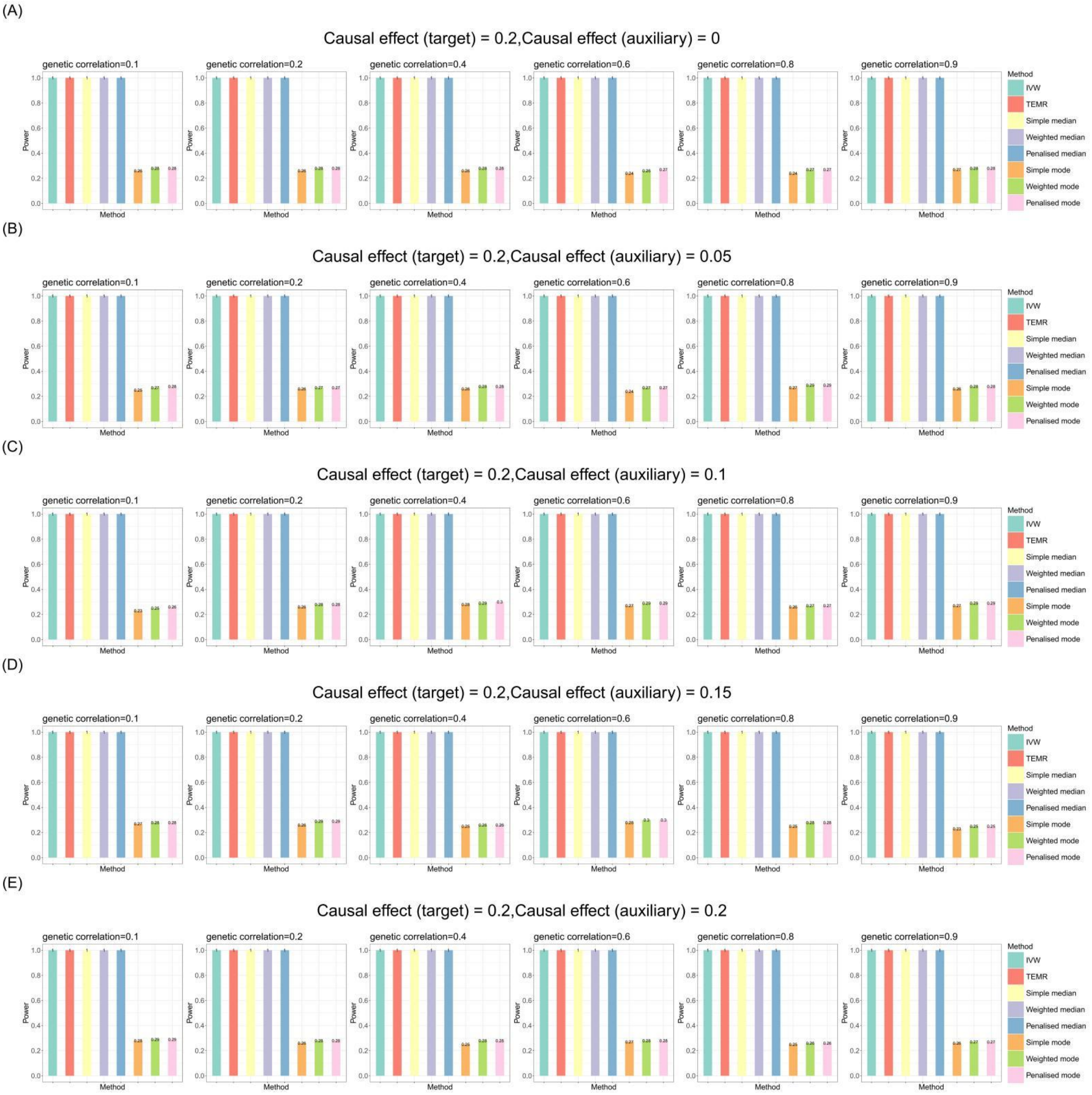


**Figure S12. Bar chart plots of simulation results with continuous outcome for causal effect estimation in the target population when there is one auxiliary population (balance horizontal pleiotropy).**

Sample size of target population is 3,000 and the sample size of auxiliary population is 300,000. IVs include 100 common SNPs. Bar chart plots illustrate the statistical power performance when the causal effect estimation is 0.2 in the target population, and from 0 to 0.2 in the auxiliary population, respectively. IVW, Inverse-variance weighted method.

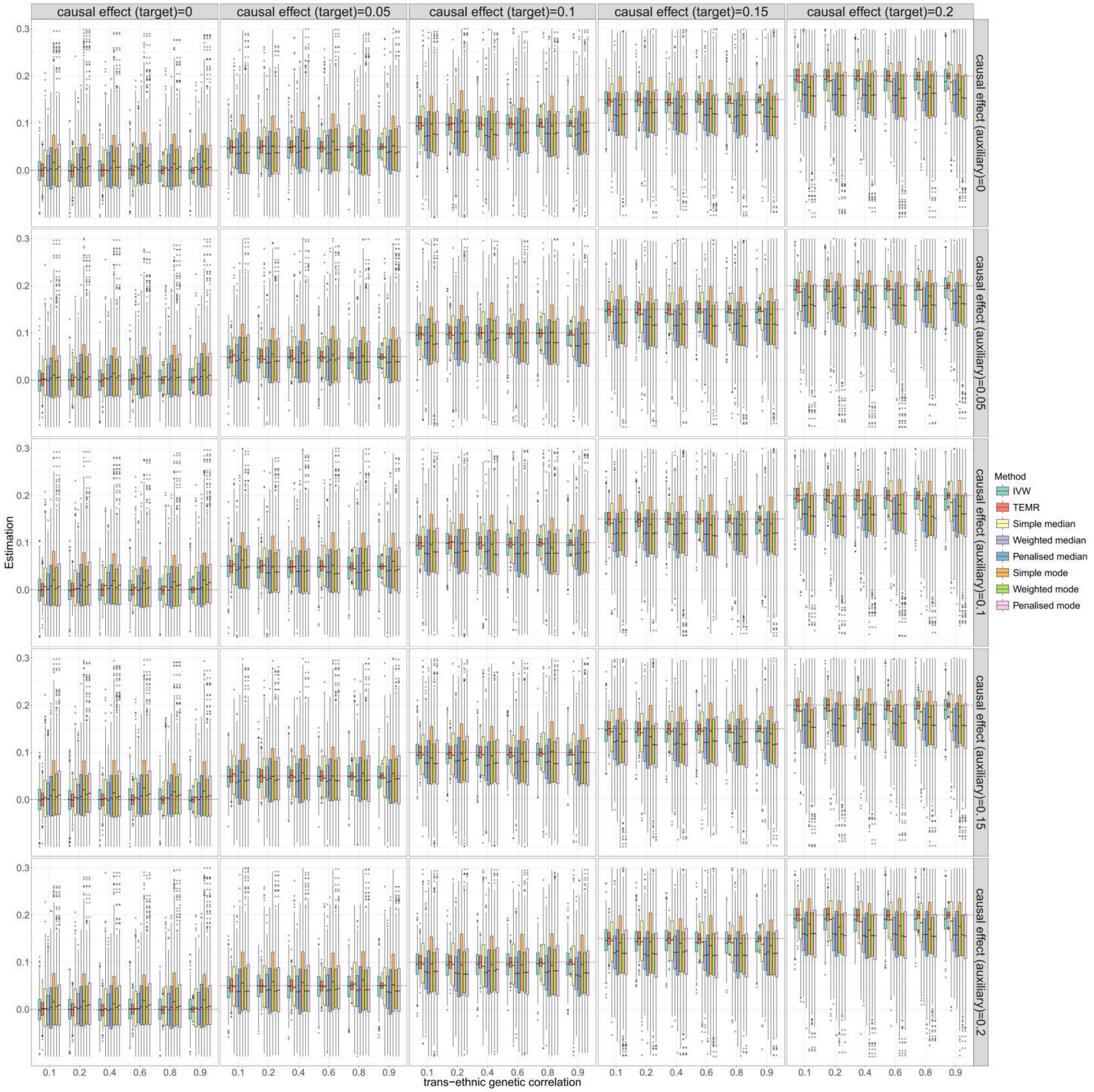


**Figure S13. Boxplots of simulation results with continuous outcome for causal effect estimation in the target population when there is one auxiliary population (directional horizontal pleiotropy).**

Sample size of target population is 3,000 and the sample size of auxiliary population is 300,000. IVs include 100 common SNPs. Boxplots show the performances of causal effect estimation in target population when the causal effect of target/auxiliary population is 0 to 0.2, respectively. IVW, Inverse-variance weighted method.

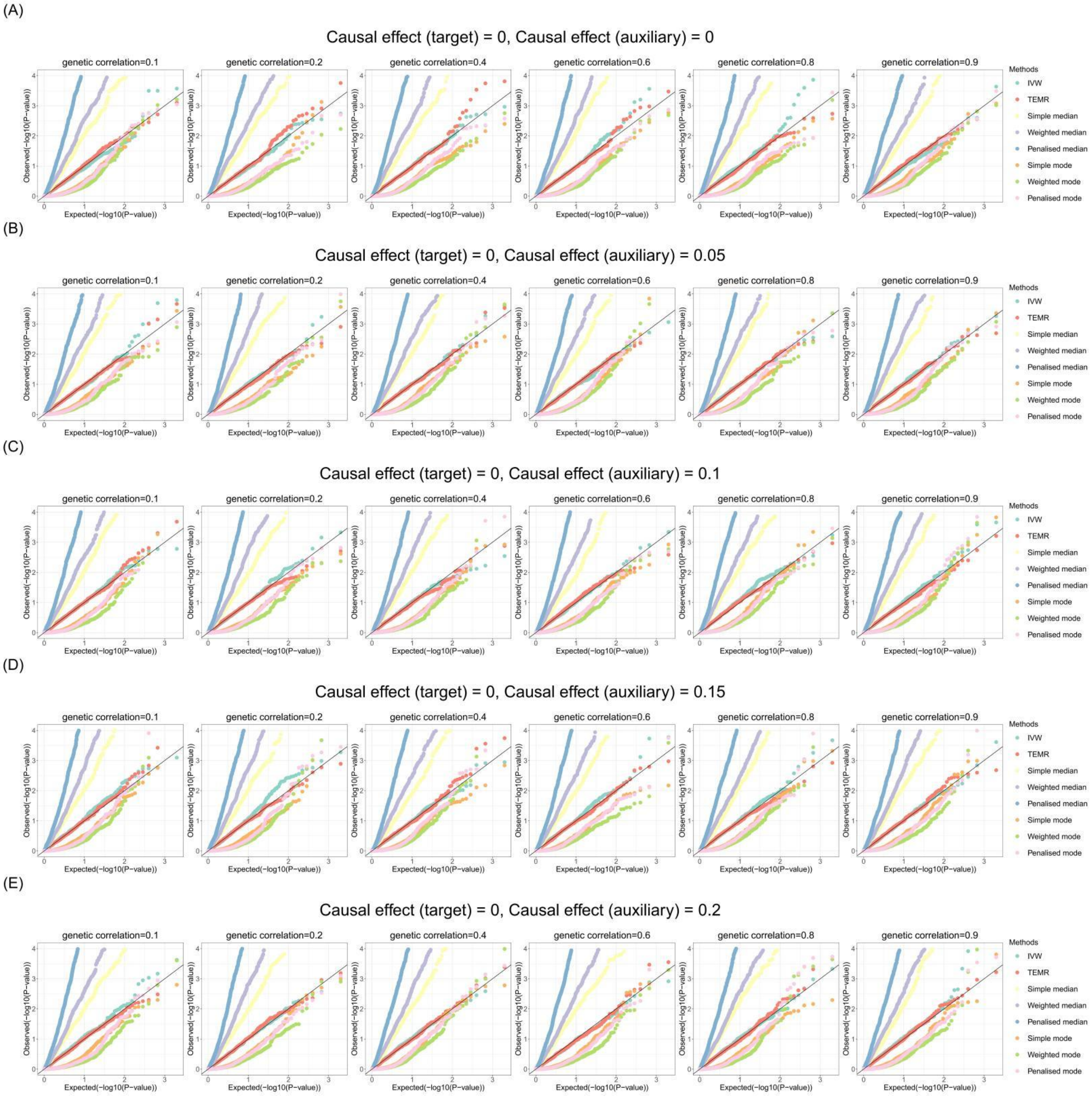


**Figure S14. Q-Q plots of simulation results with continuous outcome for causal effect estimation in the target population when there is one auxiliary population (directional horizontal pleiotropy).**

Sample size of target population is 3,000 and the sample size of auxiliary population is 300,000. IVs include 100 common SNPs. Q-Q plots show the performances of Type I error rates of zero causal effect estimation in target population when the causal effect of auxiliary population is 0 to 0.2, respectively. IVW, Inverse-variance weighted method.

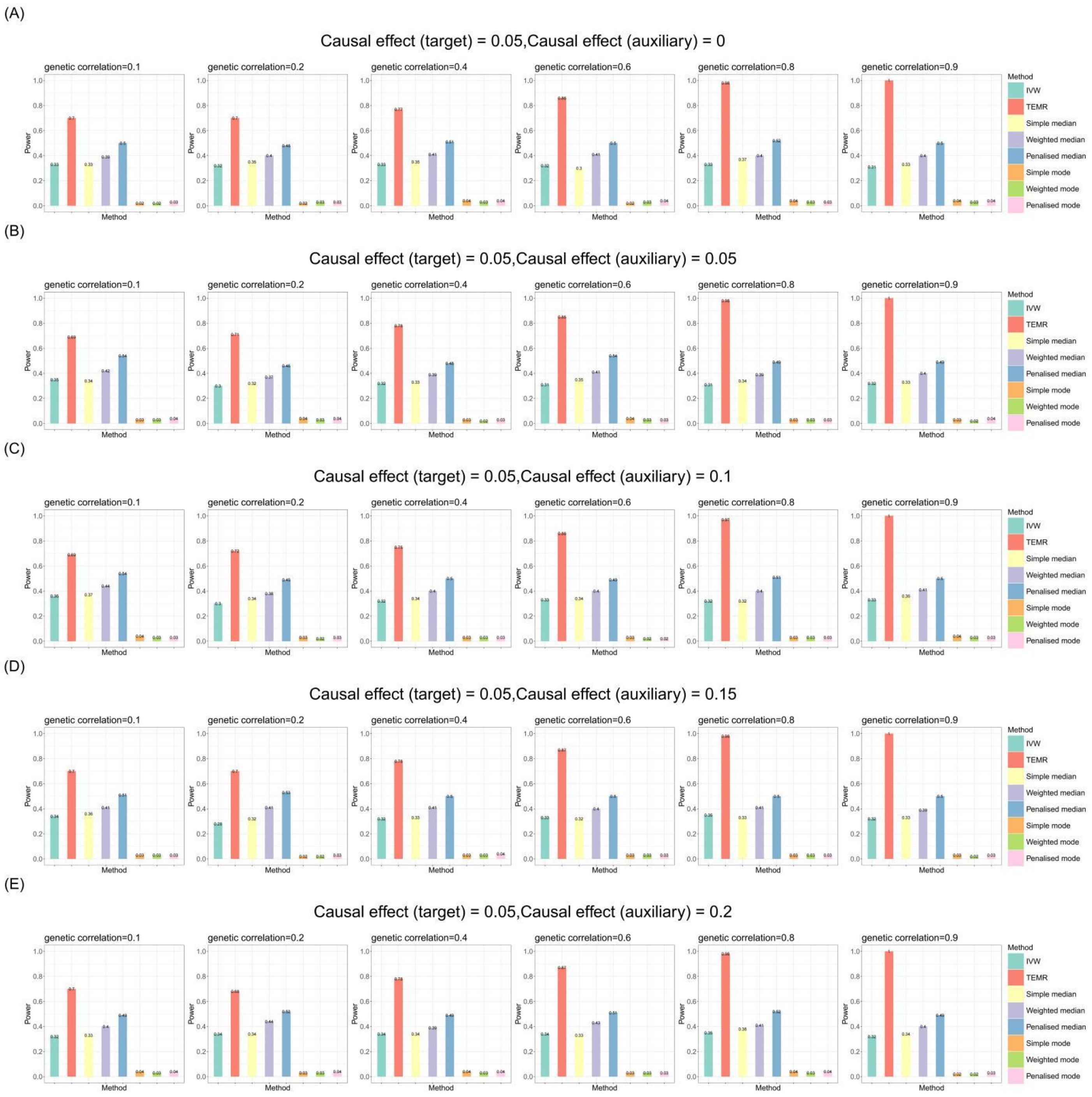


**Figure S15. Bar chart plots of simulation results with continuous outcome for causal effect estimation in the target population when there is one auxiliary population (directional horizontal pleiotropy).**

Sample size of target population is 3,000 and the sample size of auxiliary population is 300,000. IVs include 100 common SNPs. Bar chart plots illustrate the statistical power performance when the causal effect estimation is 0.05 in the target population, and from 0 to 0.2 in the auxiliary population, respectively. IVW, Inverse-variance weighted method.

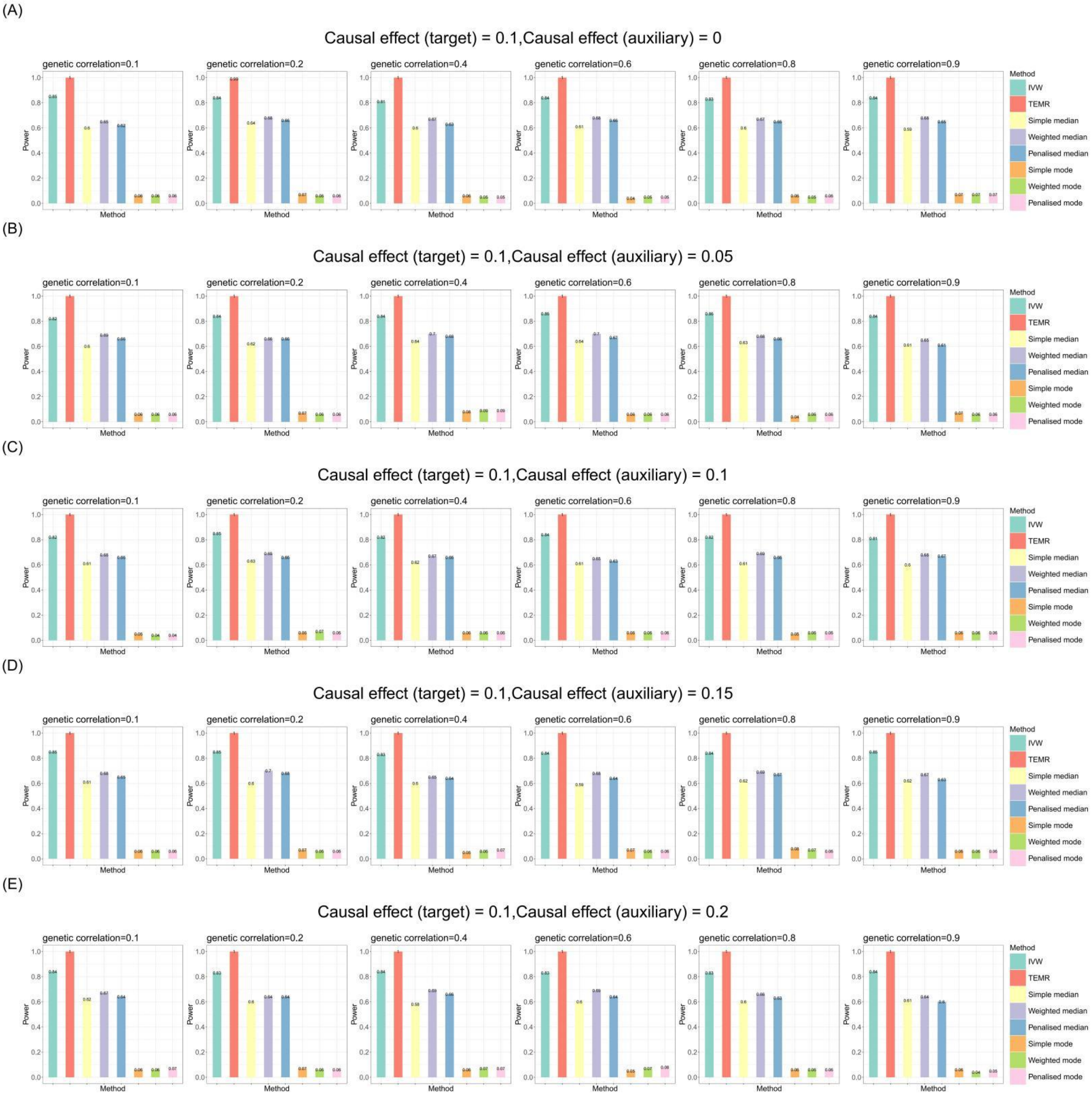


**Figure S16. Bar chart plots of simulation results with continuous outcome for causal effect estimation in the target population when there is one auxiliary population (directional horizontal pleiotropy).**

Sample size of target population is 3,000 and the sample size of auxiliary population is 300,000. IVs include 100 common SNPs. Bar chart plots illustrate the statistical power performance when the causal effect estimation is 0.1 in the target population, and from 0 to 0.2 in the auxiliary population, respectively. IVW, Inverse-variance weighted method.

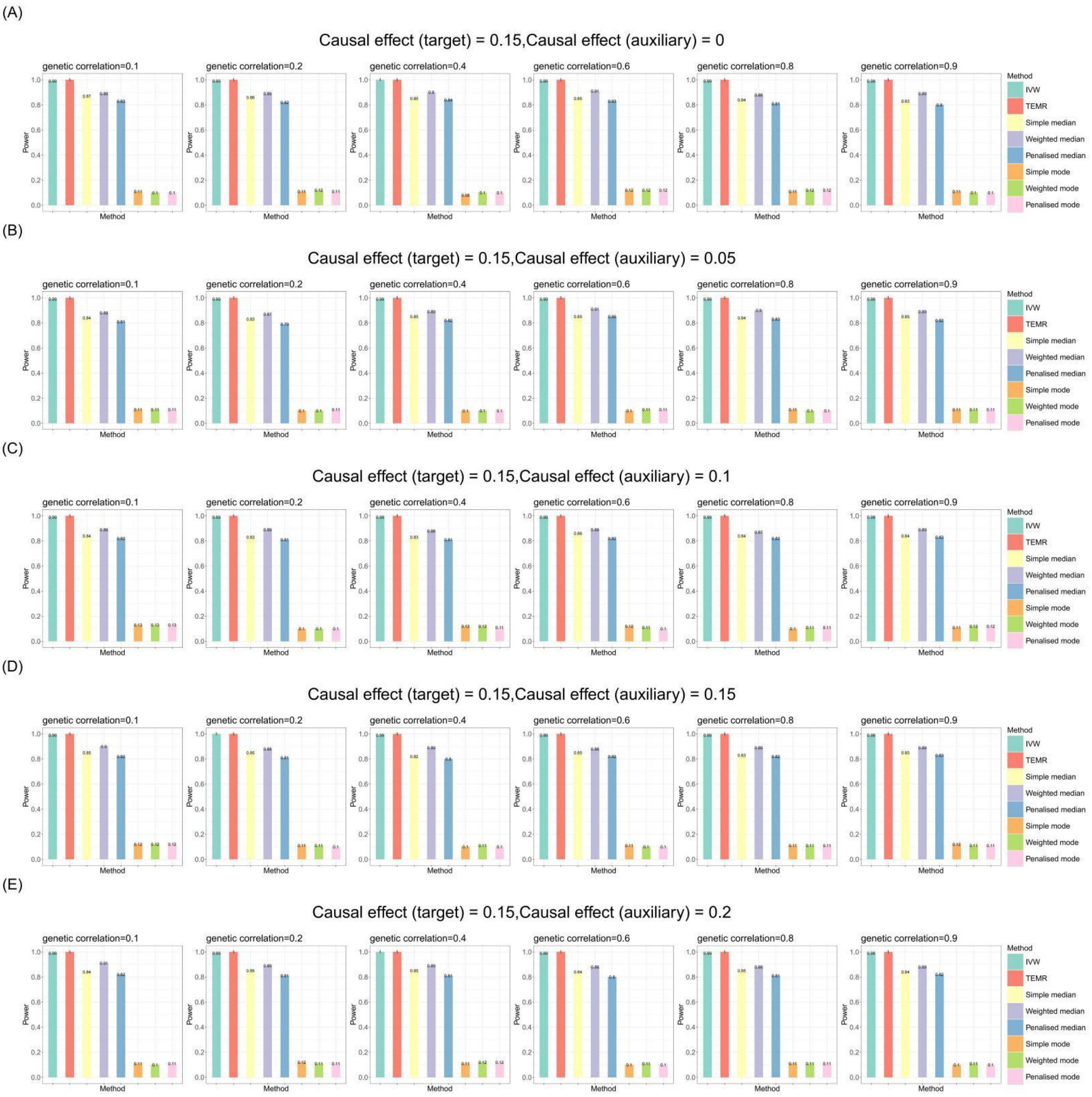


**Figure S17. Bar chart plots of simulation results with continuous outcome for causal effect estimation in the target population when there is one auxiliary population (directional horizontal pleiotropy).**

Sample size of target population is 3,000 and the sample size of auxiliary population is 300,000. IVs include 100 common SNPs. Bar chart plots illustrate the statistical power performance when the causal effect estimation is 0.15 in the target population, and from 0 to 0.2 in the auxiliary population, respectively. IVW, Inverse-variance weighted method.

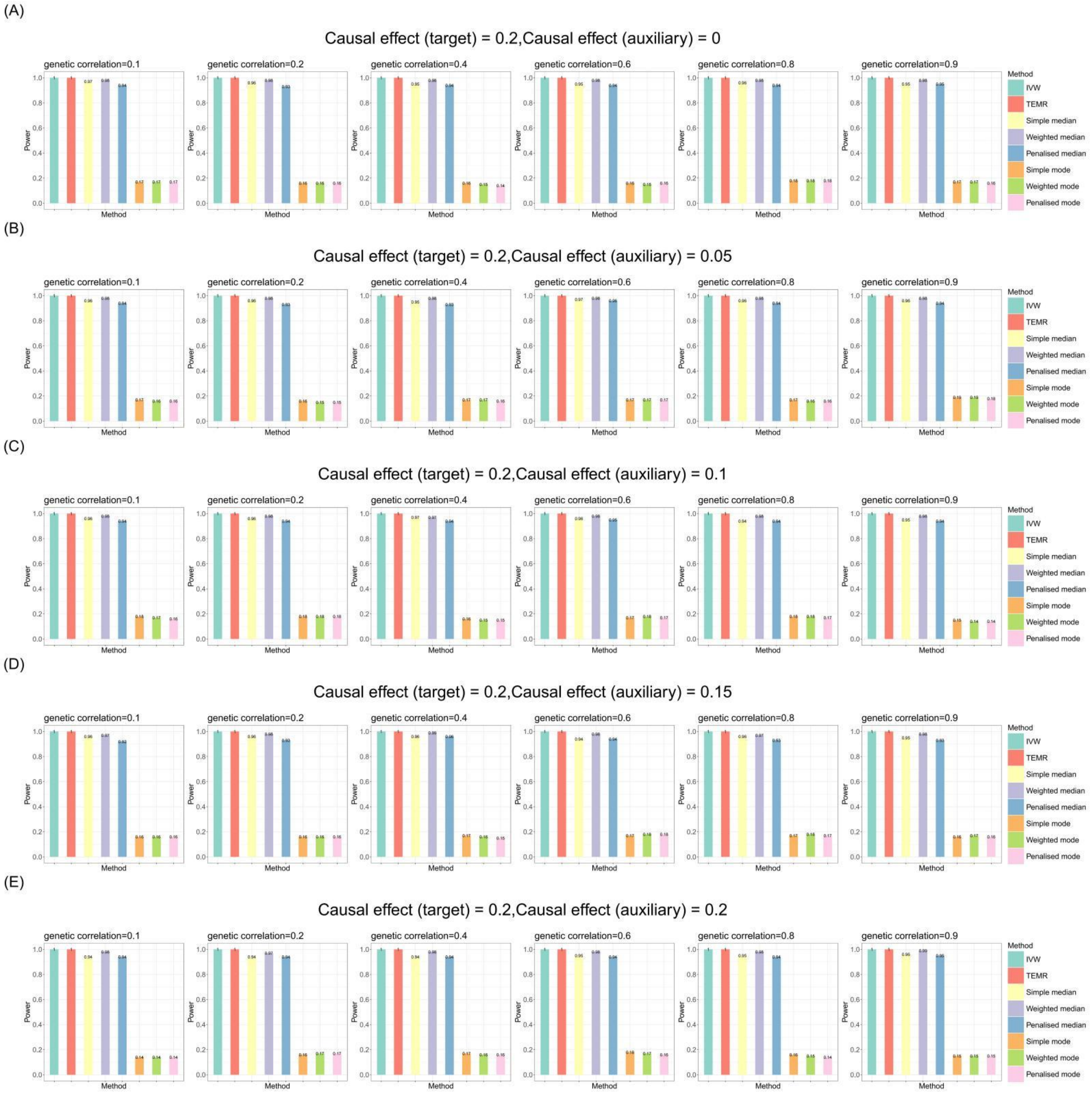


**Figure S18. Bar chart plots of simulation results with continuous outcome for causal effect estimation in the target population when there is one auxiliary population (directional horizontal pleiotropy).**

Sample size of target population is 3,000 and the sample size of auxiliary population is 300,000. IVs include 100 common SNPs. Bar chart plots illustrate the statistical power performance when the causal effect estimation is 0.2 in the target population, and from 0 to 0.2 in the auxiliary population, respectively. IVW, Inverse-variance weighted method.

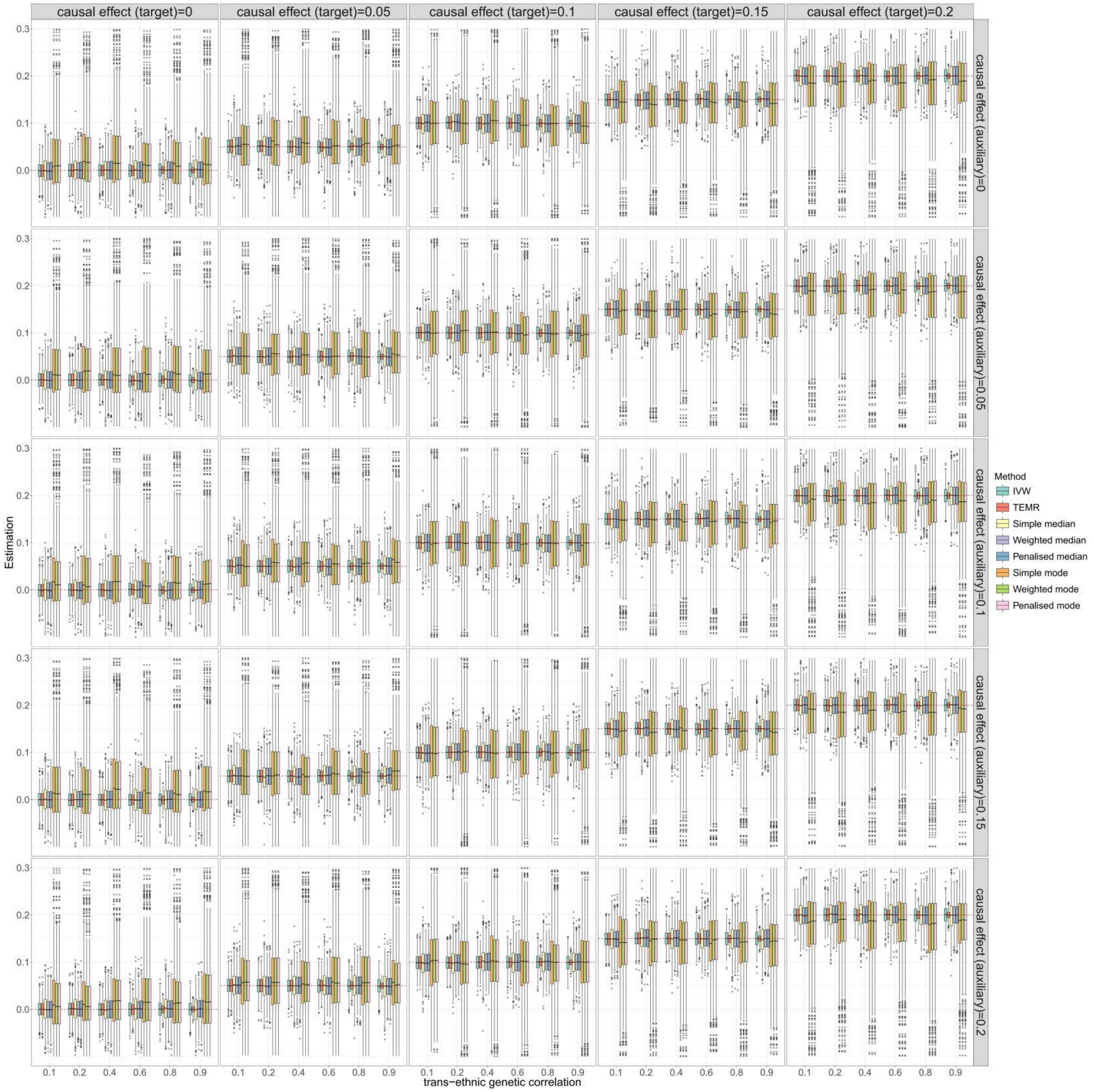


**Figure S19. Boxplots of simulation results with categorical outcome for causal effect estimation in the target population when there is one auxiliary population (no horizontal pleiotropy).** Sample size of target population is 3,000 and the sample size of auxiliary population is 300,000. IVs include 100 common SNPs. Boxplots show the performances of causal effect estimation in target population when the causal effect of target/auxiliary population is 0 to 0.2, respectively. IVW, Inverse-variance weighted method.

(A)

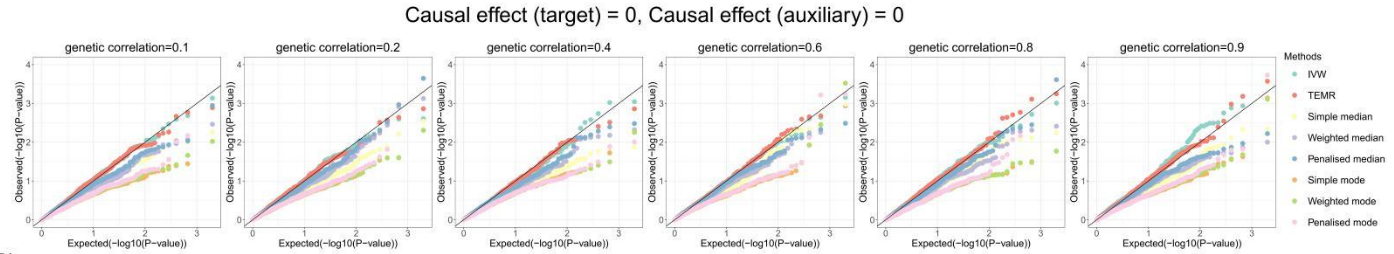


(B)

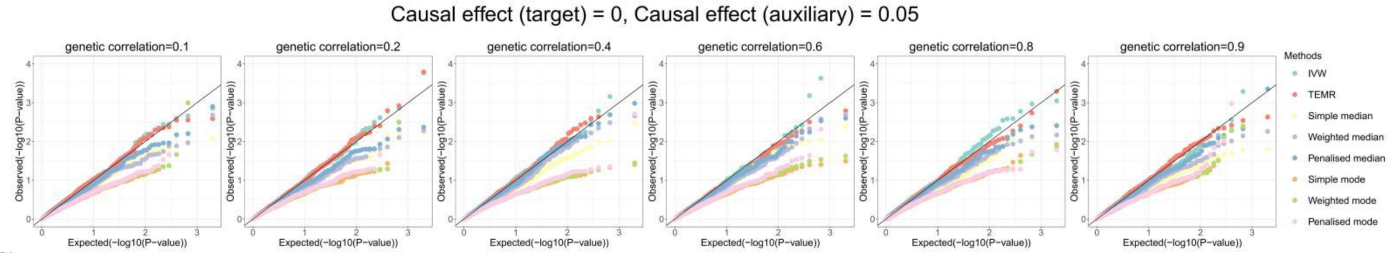


(C)

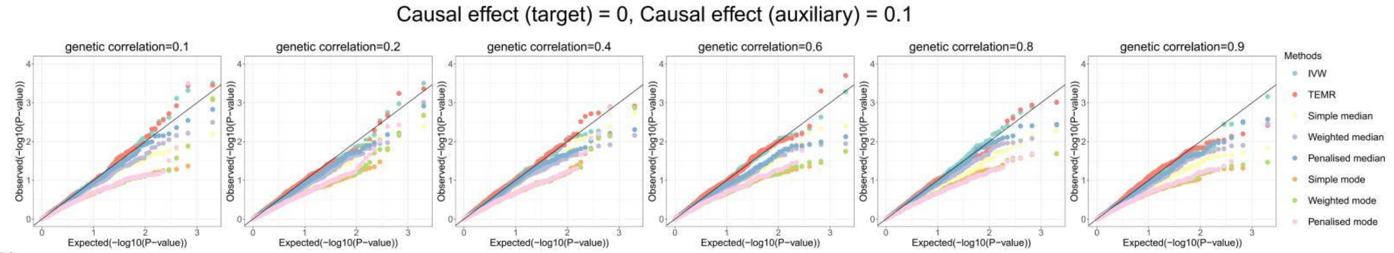


(D)

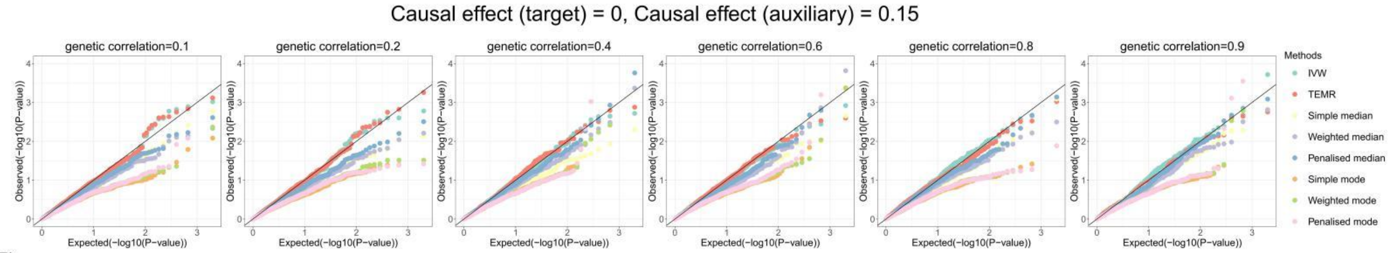


(E)

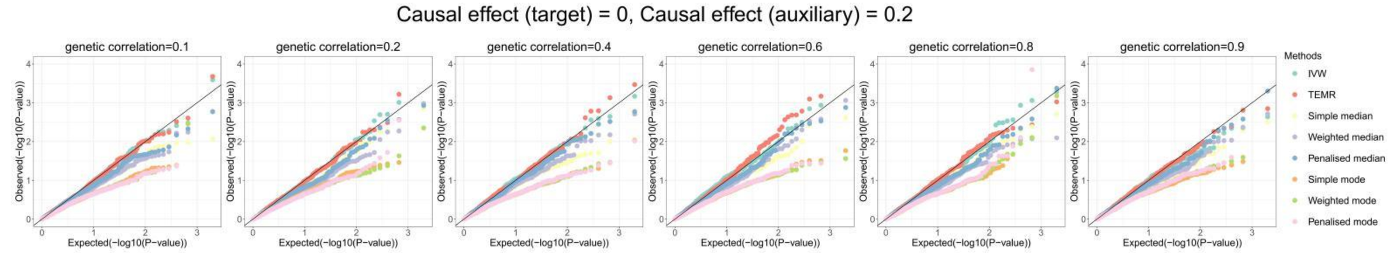


**Figure S20. Q-Q plots of simulation results with categorical outcome for causal effect estimation in the target population when there is one auxiliary population (no horizontal pleiotropy).**

Sample size of target population is 3,000 and the sample size of auxiliary population is 300,000. IVs include 100 common SNPs. Q-Q plots show the performances of Type I error rates of zero causal effect estimation in target population when the causal effect of auxiliary population is 0 to 0.2, respectively. IVW, Inverse-variance weighted method.

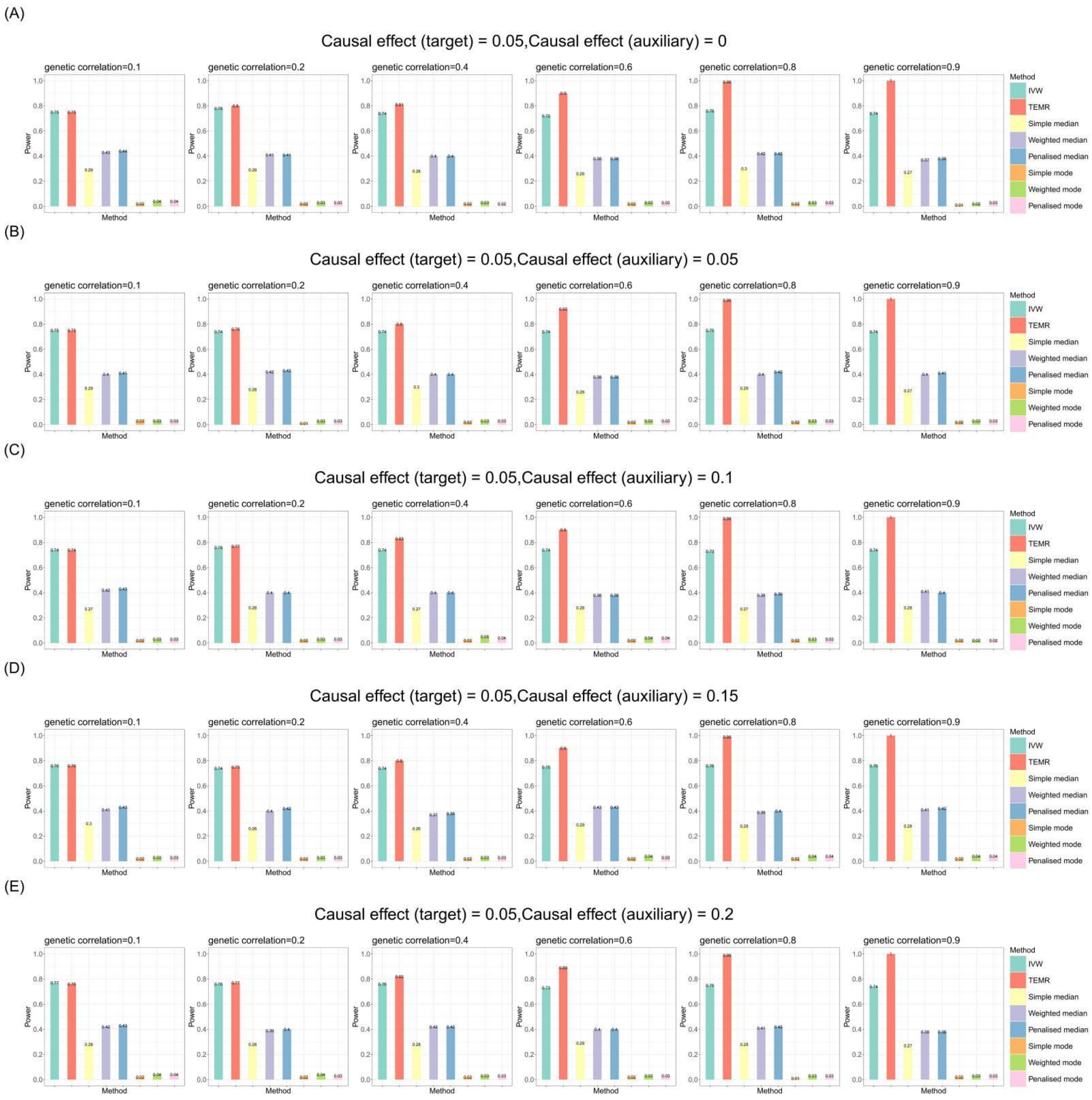


**Figure S21. Bar chart plots of simulation results with categorical outcome for causal effect estimation in the target population when there is one auxiliary population (no horizontal pleiotropy).**

Sample size of target population is 3,000 and the sample size of auxiliary population is 300,000. IVs include 100 common SNPs. Bar chart plots illustrate the statistical power performance when the causal effect estimation is 0.05 in the target population, and from 0 to 0.2 in the auxiliary population, respectively. IVW, Inverse-variance weighted method.

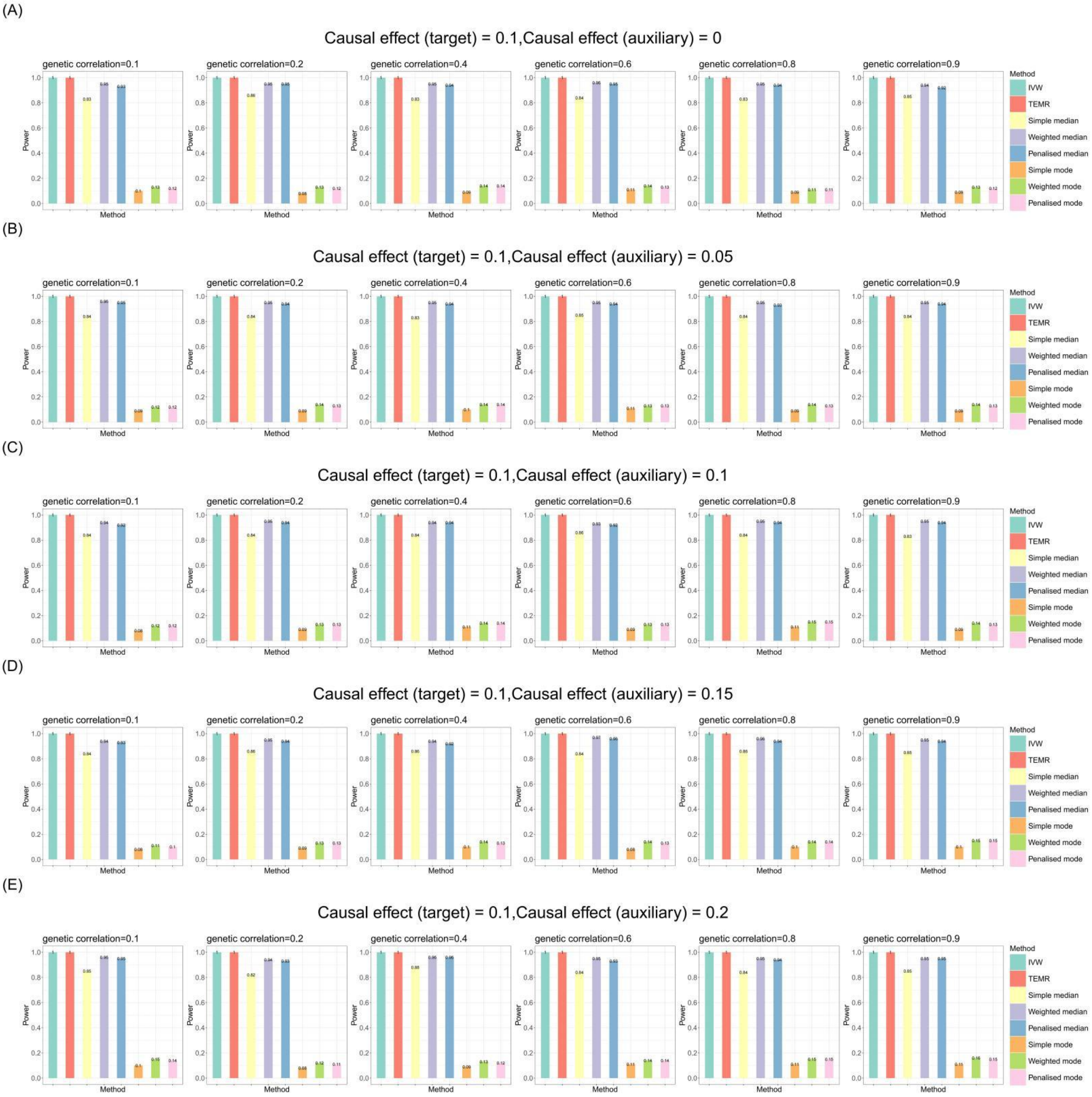


**Figure S22. Bar chart plots of simulation results with categorical outcome for causal effect estimation in the target population when there is one auxiliary population (no horizontal pleiotropy).**

Sample size of target population is 3,000 and the sample size of auxiliary population is 300,000. IVs include 100 common SNPs. Bar chart plots illustrate the statistical power performance when the causal effect estimation is 0.1 in the target population, and from 0 to 0.2 in the auxiliary population, respectively. IVW, Inverse-variance weighted method.

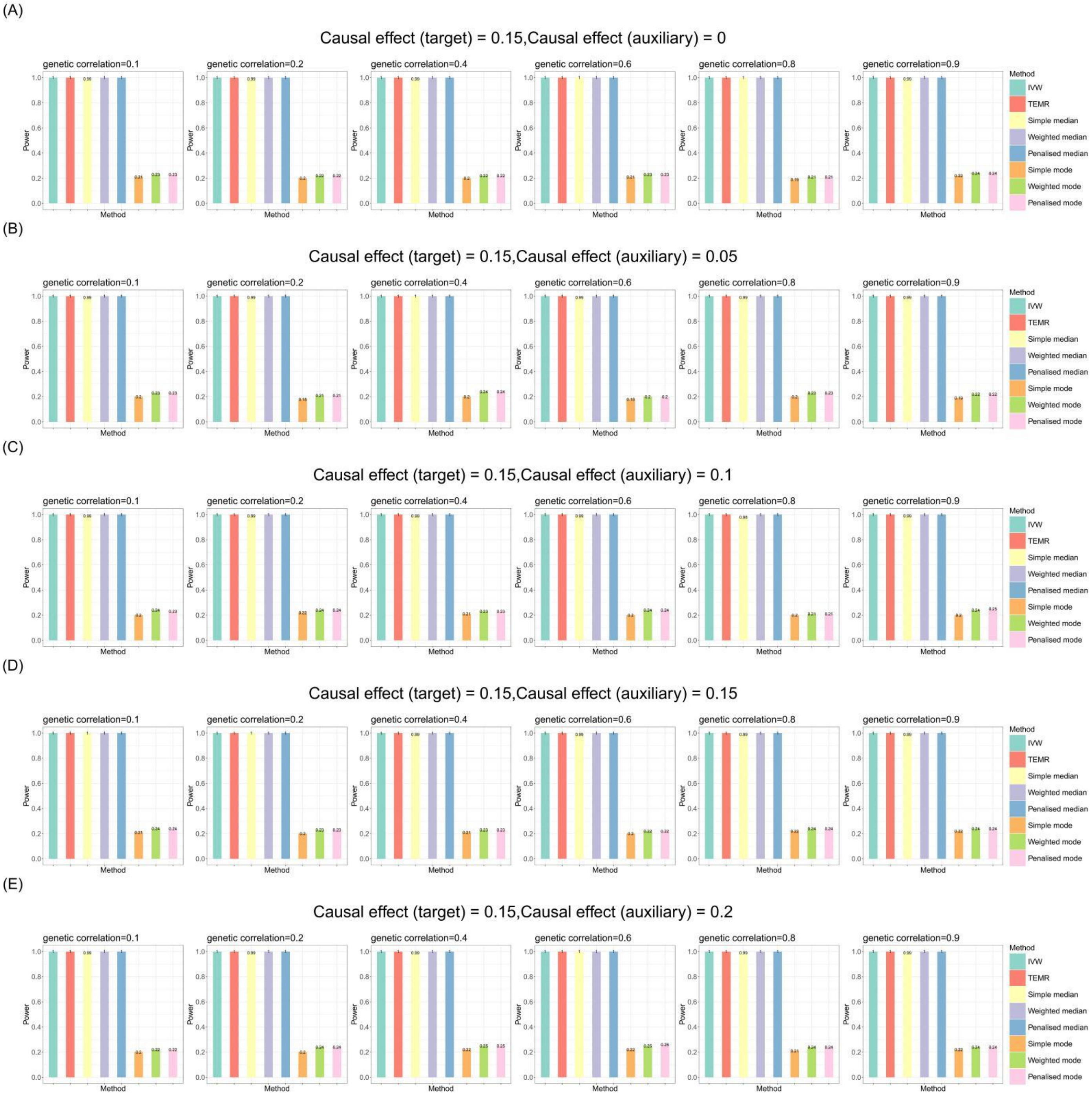


**Figure S23. Bar chart plots of simulation results with categorical outcome for causal effect estimation in the target population when there is one auxiliary population (no horizontal pleiotropy).**

Sample size of target population is 3,000 and the sample size of auxiliary population is 300,000. IVs include 100 common SNPs. Bar chart plots illustrate the statistical power performance when the causal effect estimation is 0.15 in the target population, and from 0 to 0.2 in the auxiliary population, respectively. IVW, Inverse-variance weighted method.

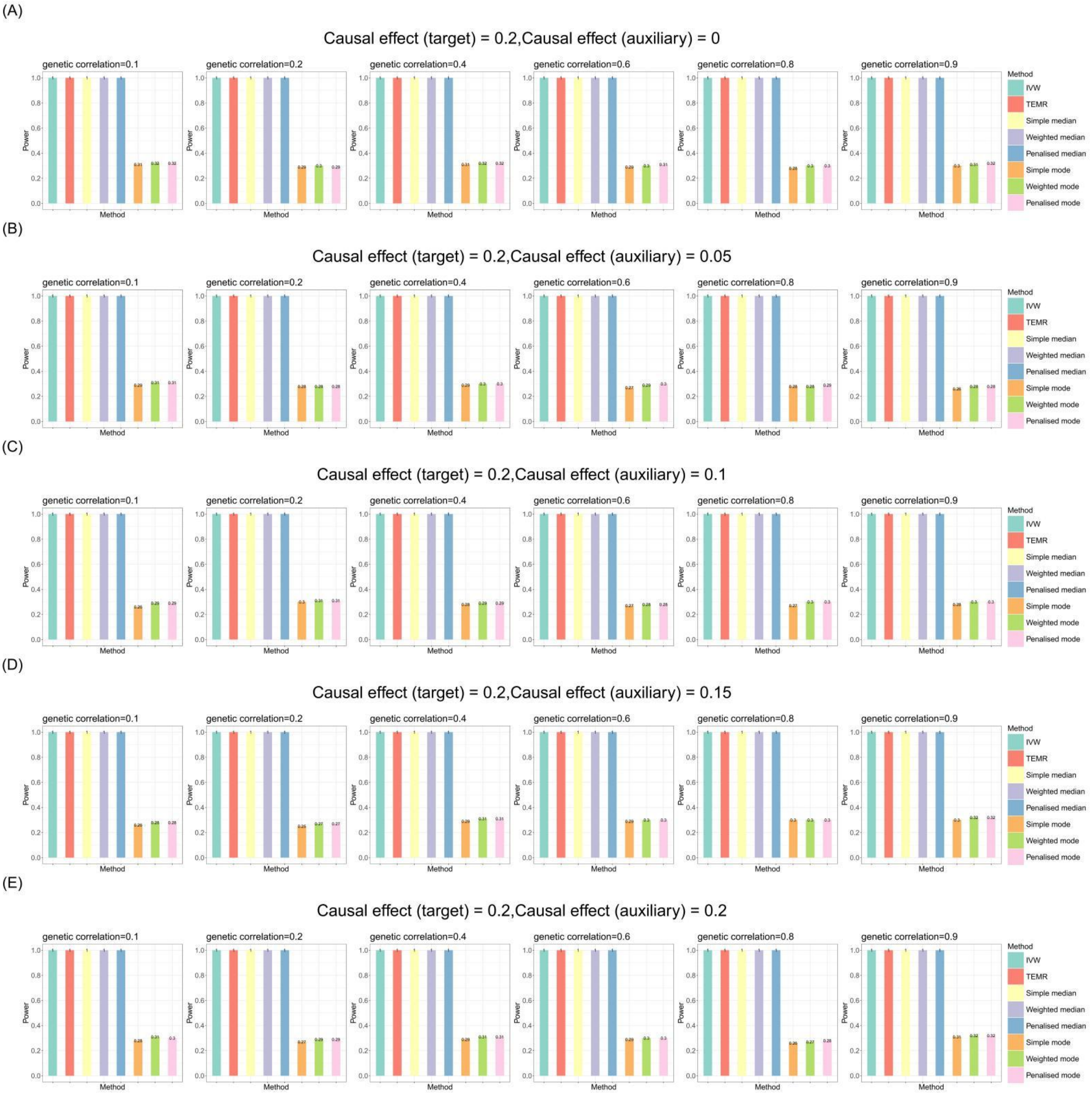


**Figure S24. Bar chart plots of simulation results with categorical outcome for causal effect estimation in the target population when there is one auxiliary population (no horizontal pleiotropy).**

Sample size of target population is 3,000 and the sample size of auxiliary population is 300,000. IVs include 100 common SNPs. Bar chart plots illustrate the statistical power performance when the causal effect estimation is 0.2 in the target population, and from 0 to 0.2 in the auxiliary population, respectively. IVW, Inverse-variance weighted method.

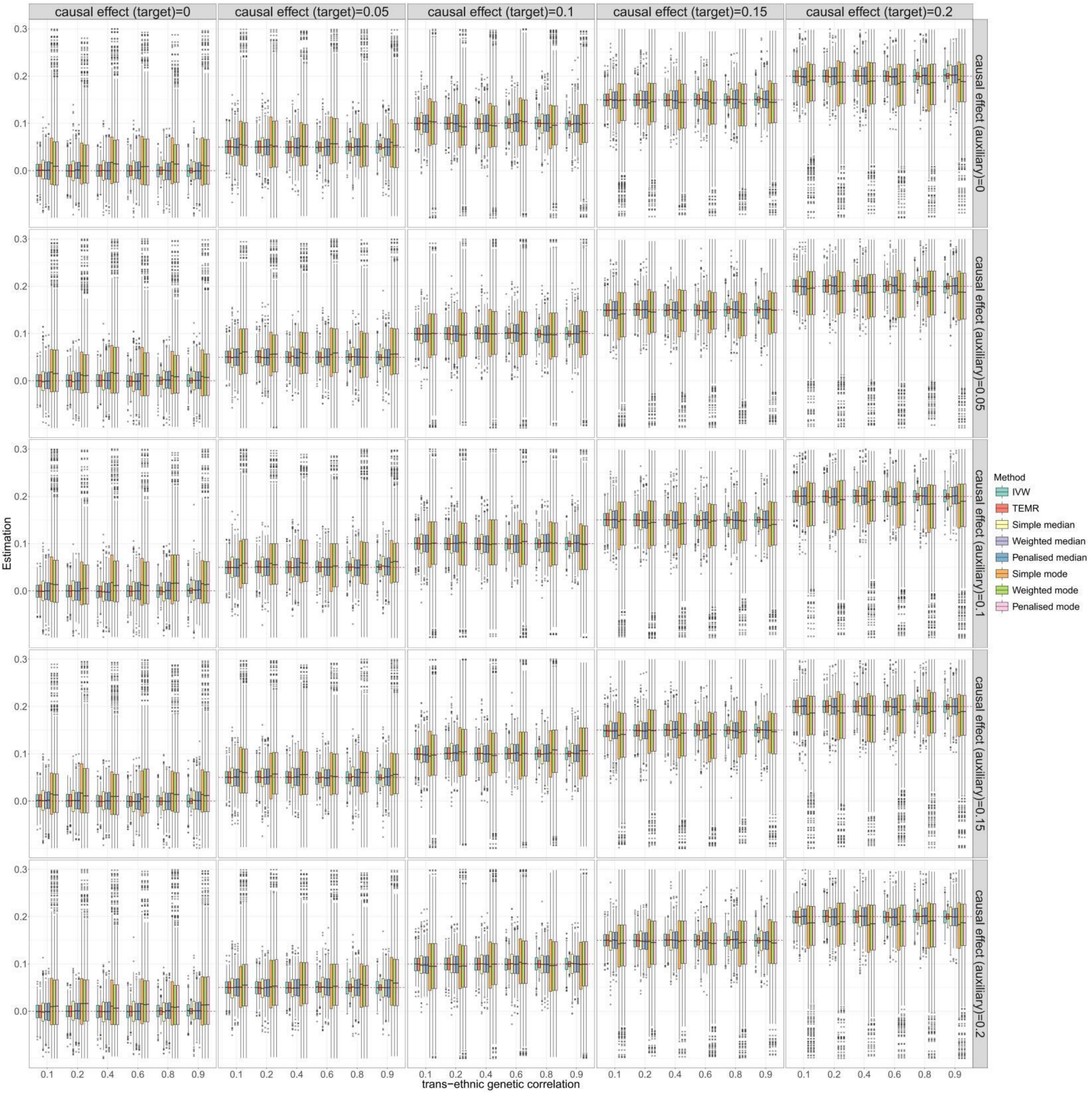


**Figure S25. Boxplots of simulation results with categorical outcome for causal effect estimation in the target population when there is one auxiliary population (balance horizontal pleiotropy).** Sample size of target population is 3,000 and the sample size of auxiliary population is 300,000. IVs include 100 common SNPs. Boxplots show the performances of causal effect estimation in target population when the causal effect of target/auxiliary population is 0 to 0.2, respectively. IVW, Inverse-variance weighted method.

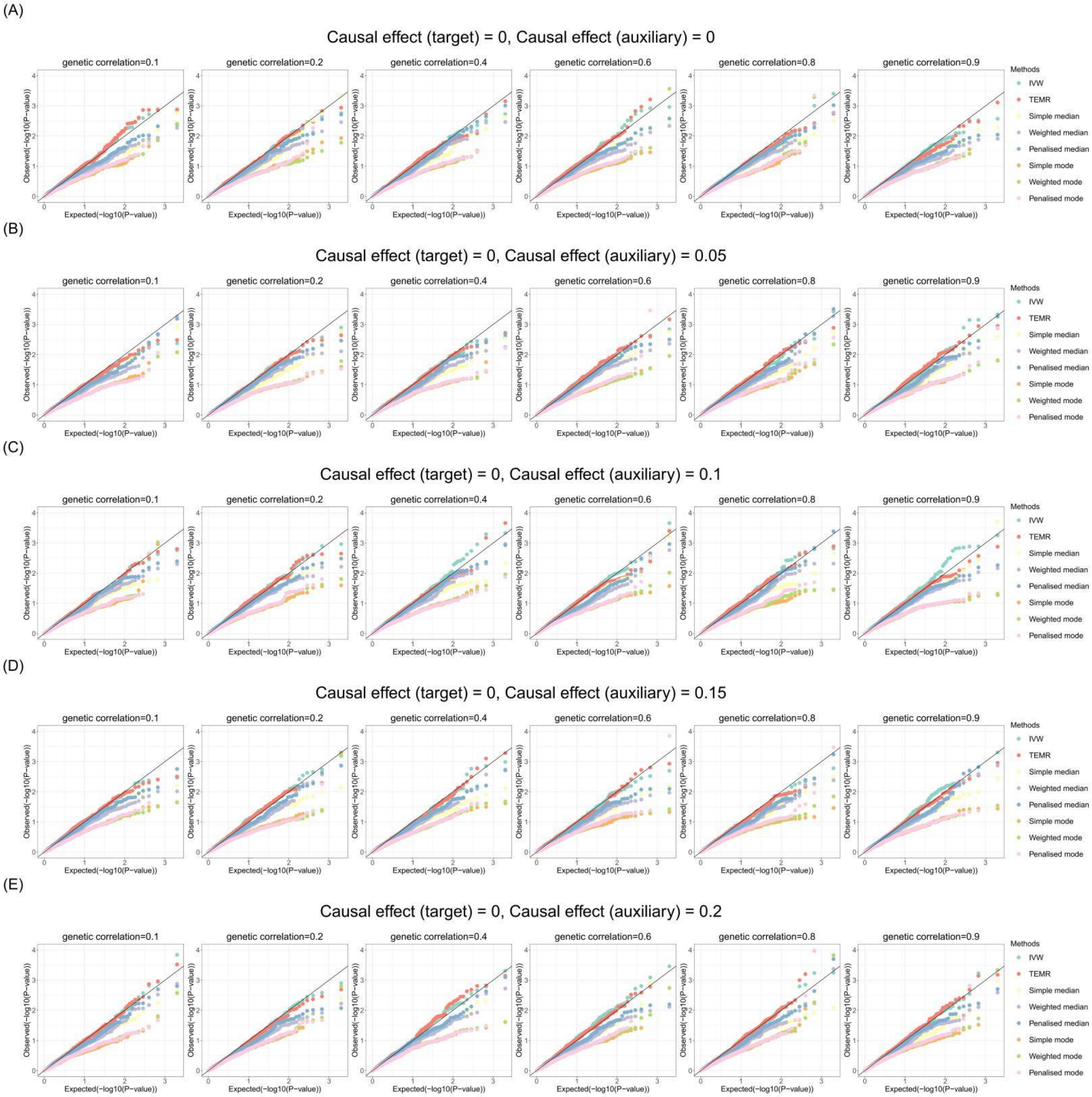


**Figure S26. Q-Q plots of simulation results with categorical outcome for causal effect estimation in the target population when there is one auxiliary population (balance horizontal pleiotropy).**

Sample size of target population is 3,000 and the sample size of auxiliary population is 300,000. IVs include 100 common SNPs. Q-Q plots show the performances of Type I error rates of zero causal effect estimation in target population when the causal effect of auxiliary population is 0 to 0.2, respectively. IVW, Inverse-variance weighted method.

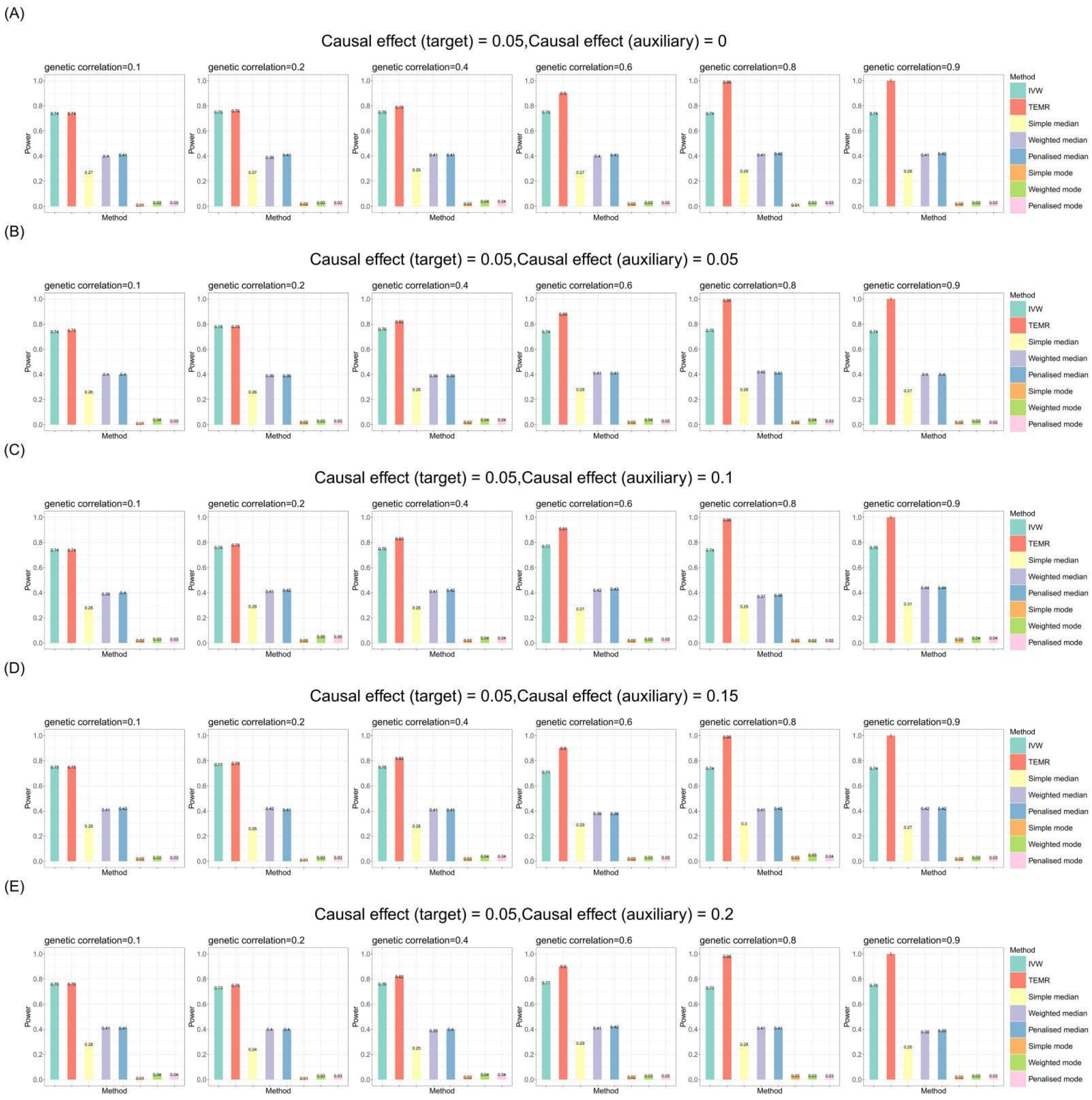

**Figure S27. Bar chart plots of simulation results with categorical outcome for causal effect estimation in the target population when there is one auxiliary population (balance horizontal pleiotropy).**

Sample size of target population is 3,000 and the sample size of auxiliary population is 300,000. IVs include 100 common SNPs. Bar chart plots illustrate the statistical power performance when the causal effect estimation is 0.05 in the target population, and from 0 to 0.2 in the auxiliary population, respectively. IVW, Inverse-variance weighted method.

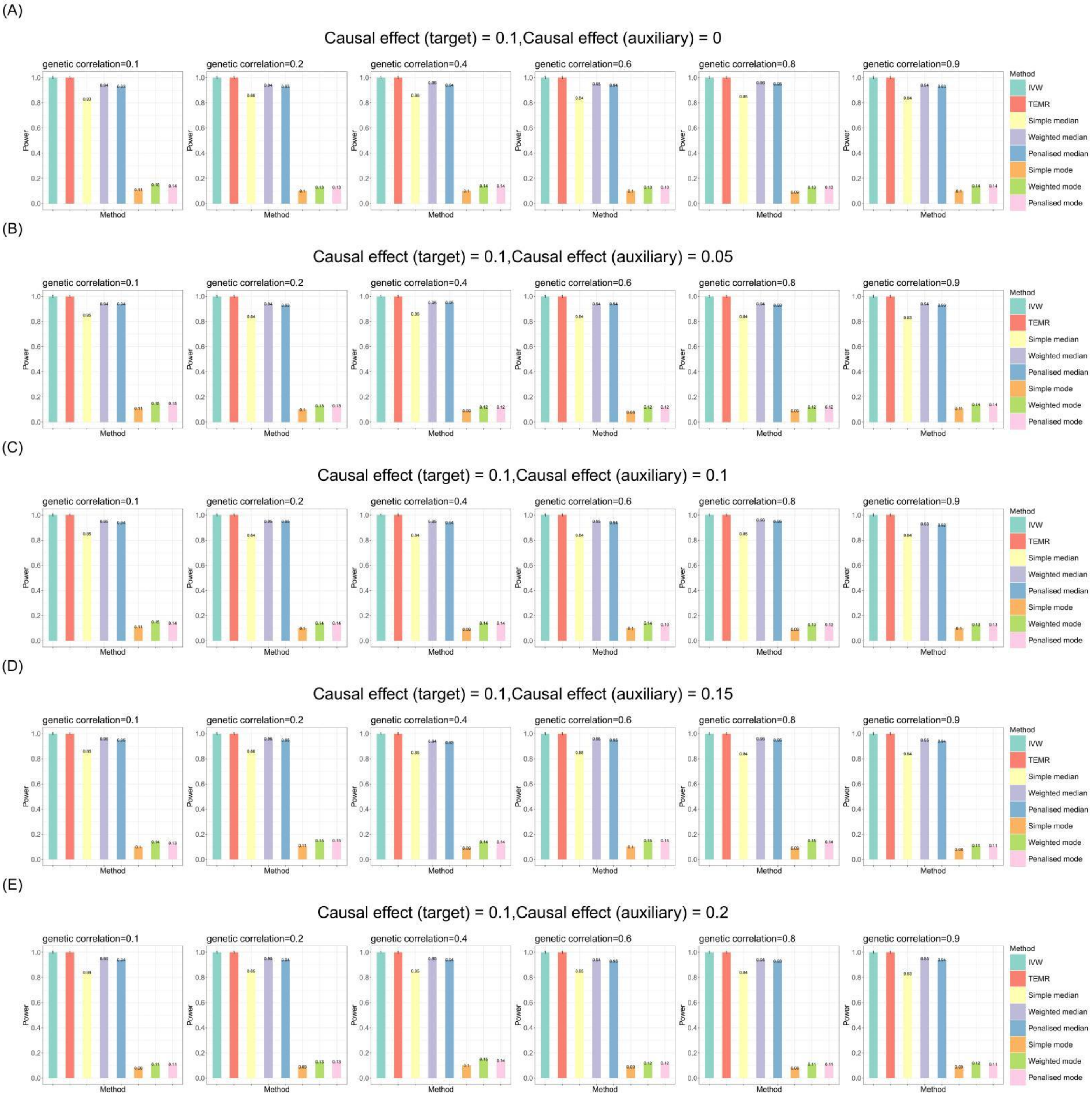


**Figure S28. Bar chart plots of simulation results with categorical outcome for causal effect estimation in the target population when there is one auxiliary population (balance horizontal pleiotropy).**

Sample size of target population is 3,000 and the sample size of auxiliary population is 300,000. IVs include 100 common SNPs. Bar chart plots illustrate the statistical power performance when the causal effect estimation is 0.1 in the target population, and from 0 to 0.2 in the auxiliary population, respectively. IVW, Inverse-variance weighted method.

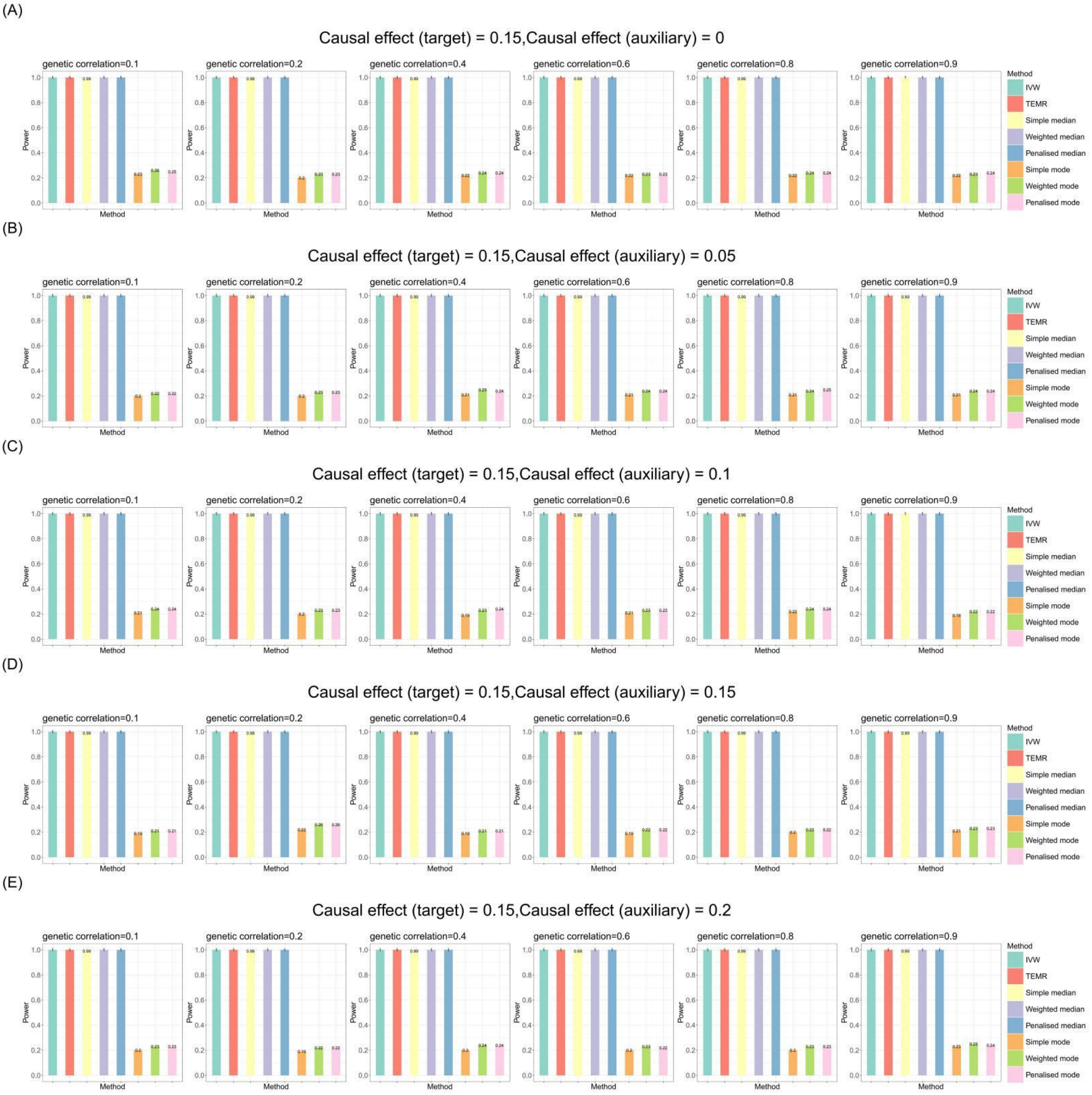


**Figure S29. Bar chart plots of simulation results with categorical outcome for causal effect estimation in the target population when there is one auxiliary population (balance horizontal pleiotropy).**

Sample size of target population is 3,000 and the sample size of auxiliary population is 300,000. IVs include 100 common SNPs. Bar chart plots illustrate the statistical power performance when the causal effect estimation is 0.15 in the target population, and from 0 to 0.2 in the auxiliary population, respectively. IVW, Inverse-variance weighted method.

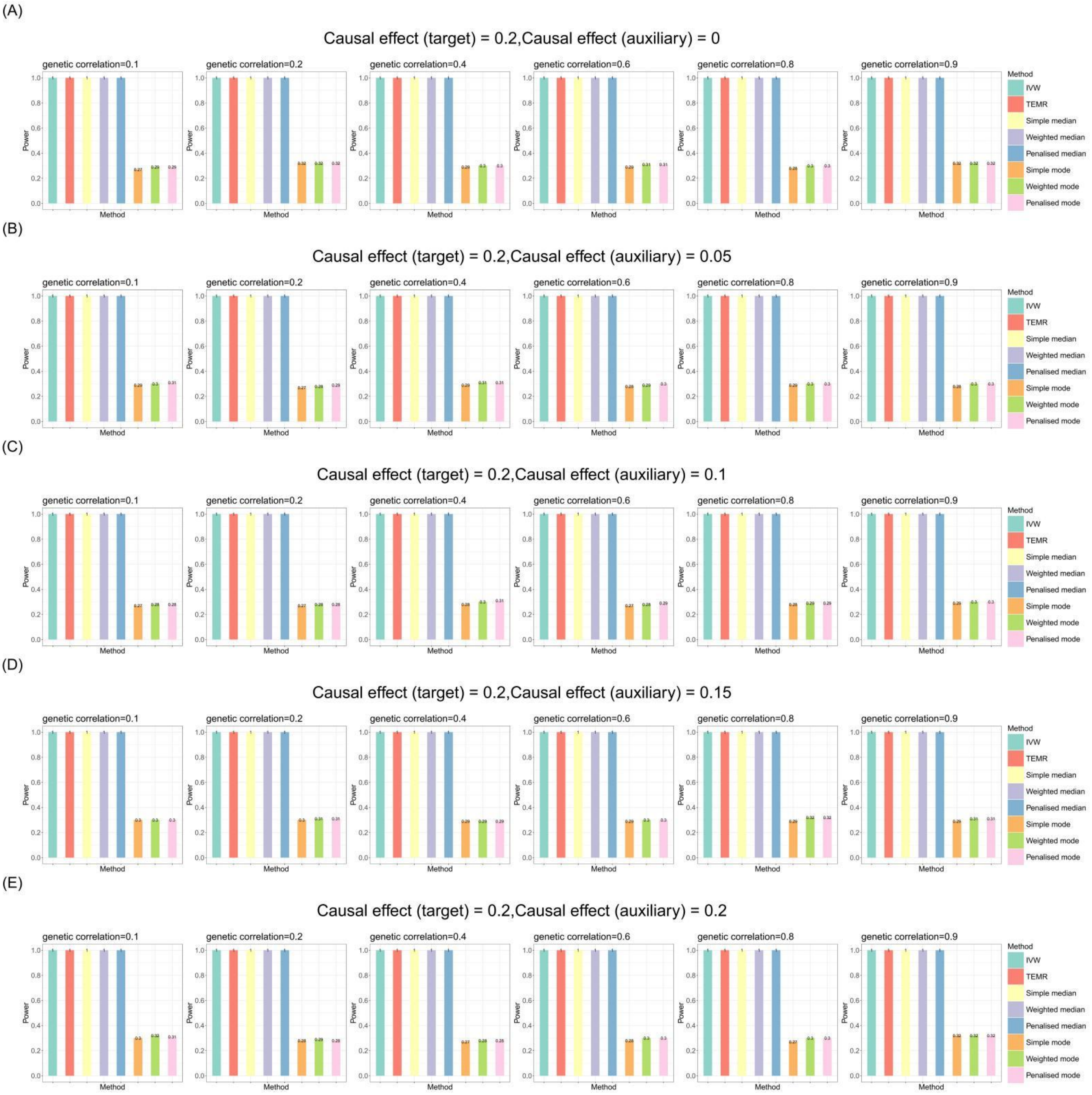


**Figure S30. Bar chart plots of simulation results with categorical outcome for causal effect estimation in the target population when there is one auxiliary population (balance horizontal pleiotropy).**

Sample size of target population is 3,000 and the sample size of auxiliary population is 300,000. IVs include 100 common SNPs. Bar chart plots illustrate the statistical power performance when the causal effect estimation is 0.2 in the target population, and from 0 to 0.2 in the auxiliary population, respectively. IVW, Inverse-variance weighted method.

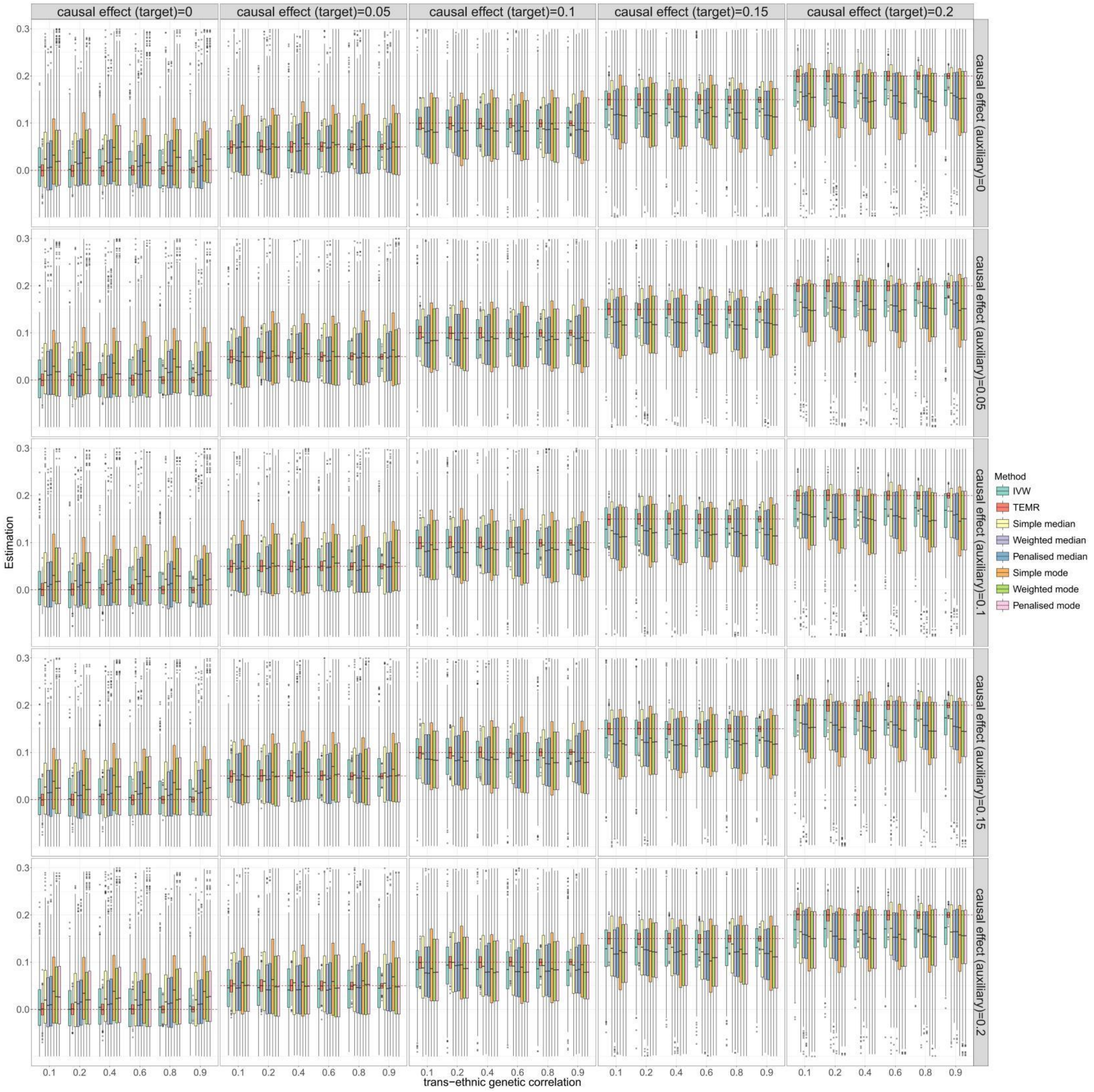


**Figure S31. Boxplots of simulation results with categorical outcome for causal effect estimation in the target population when there is one auxiliary population (directional horizontal pleiotropy).**

Sample size of target population is 3,000 and the sample size of auxiliary population is 300,000. IVs include 100 common SNPs. Boxplots show the performances of causal effect estimation in target population when the causal effect of target/auxiliary population is 0 to 0.2, respectively. IVW, Inverse-variance weighted method.

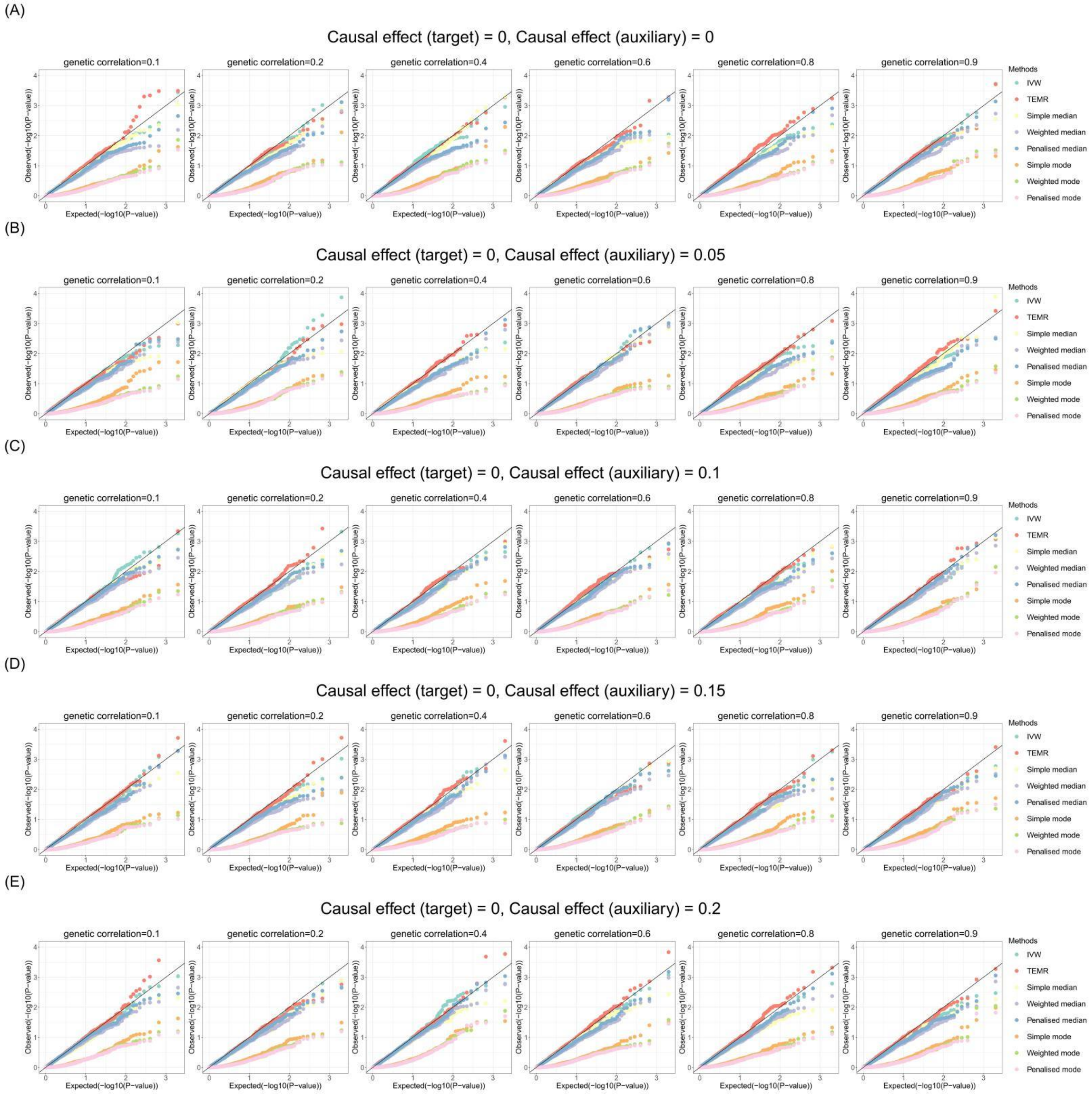


**Figure S32. Q-Q plots of simulation results with categorical outcome for causal effect estimation in the target population when there is one auxiliary population (directional horizontal pleiotropy).**

Sample size of target population is 3,000 and the sample size of auxiliary population is 300,000. IVs include 100 common SNPs. Q-Q plots show the performances of Type I error rates of zero causal effect estimation in target population when the causal effect of auxiliary population is 0 to 0.2, respectively. IVW, Inverse-variance weighted method.

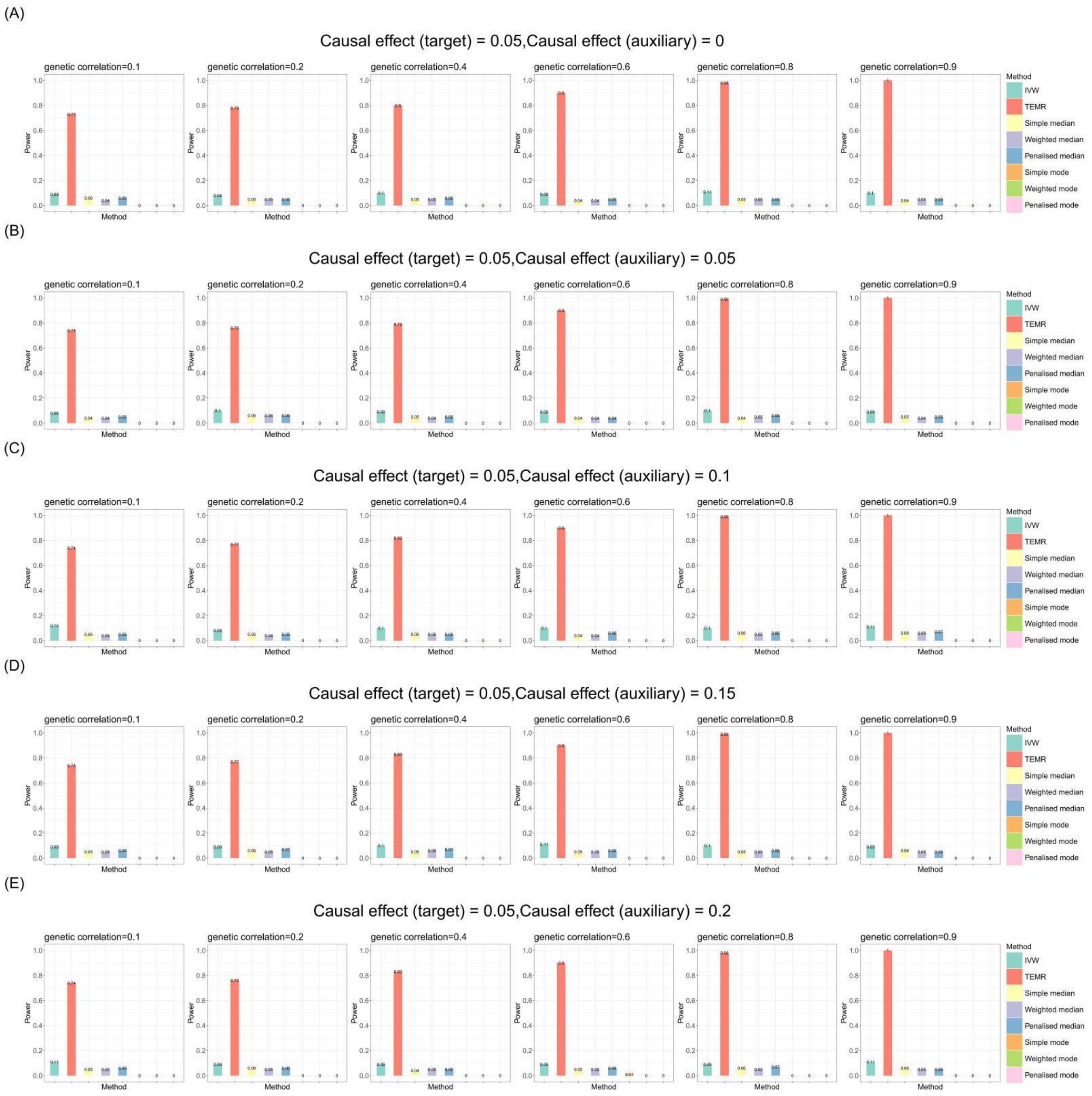


**Figure S33. Bar chart plots of simulation results with categorical outcome for causal effect estimation in the target population when there is one auxiliary population (directional horizontal pleiotropy).**

Sample size of target population is 3,000 and the sample size of auxiliary population is 300,000. IVs include 100 common SNPs. Bar chart plots illustrate the statistical power performance when the causal effect estimation is 0.05 in the target population, and from 0 to 0.2 in the auxiliary population, respectively. IVW, Inverse-variance weighted method.

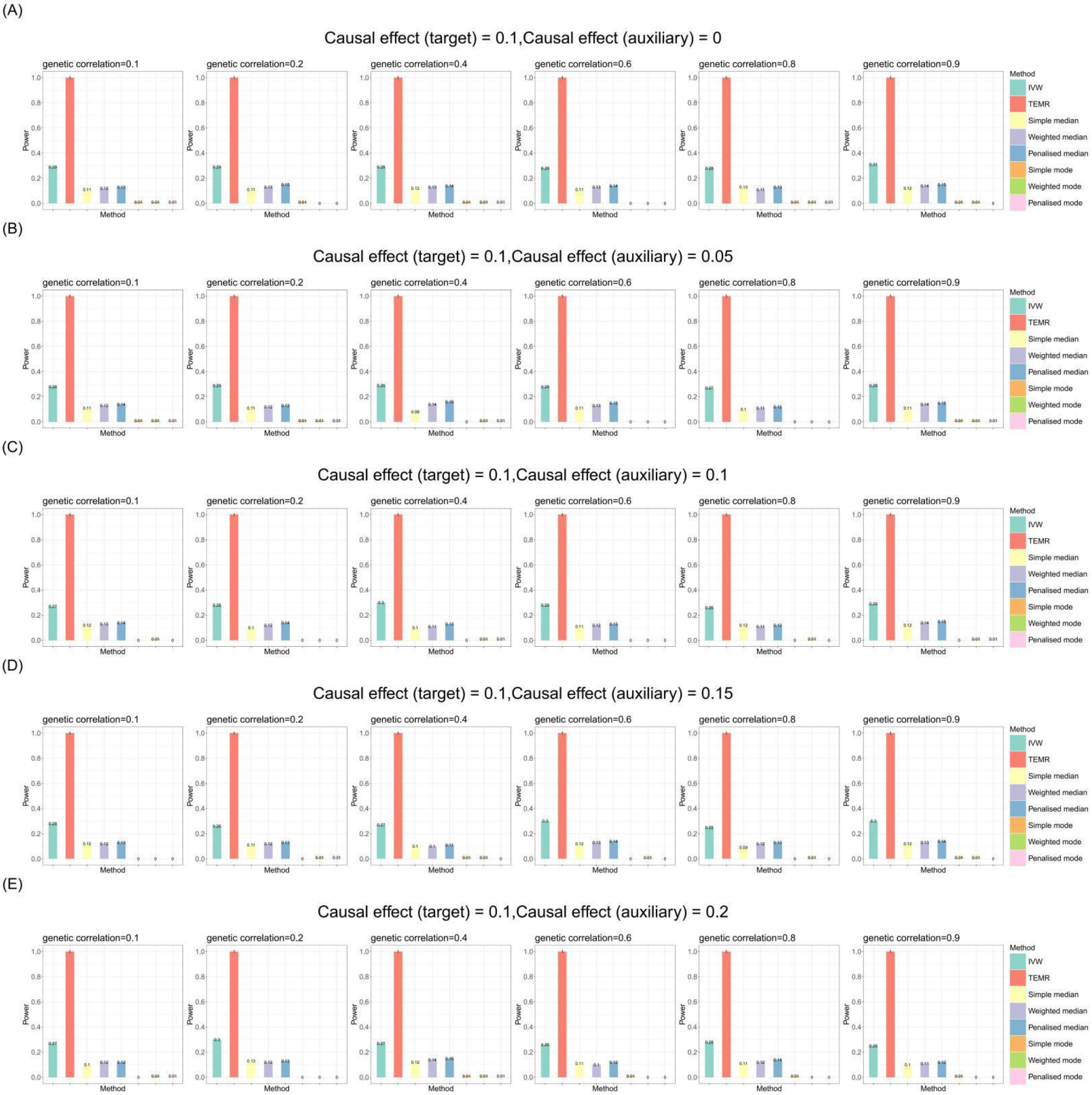


**Figure S34. Bar chart plots of simulation results with categorical outcome for causal effect estimation in the target population when there is one auxiliary population (directional horizontal pleiotropy).**

Sample size of target population is 3,000 and the sample size of auxiliary population is 300,000. IVs include 100 common SNPs. Bar chart plots illustrate the statistical power performance when the causal effect estimation is 0.1 in the target population, and from 0 to 0.2 in the auxiliary population, respectively. IVW, Inverse-variance weighted method.

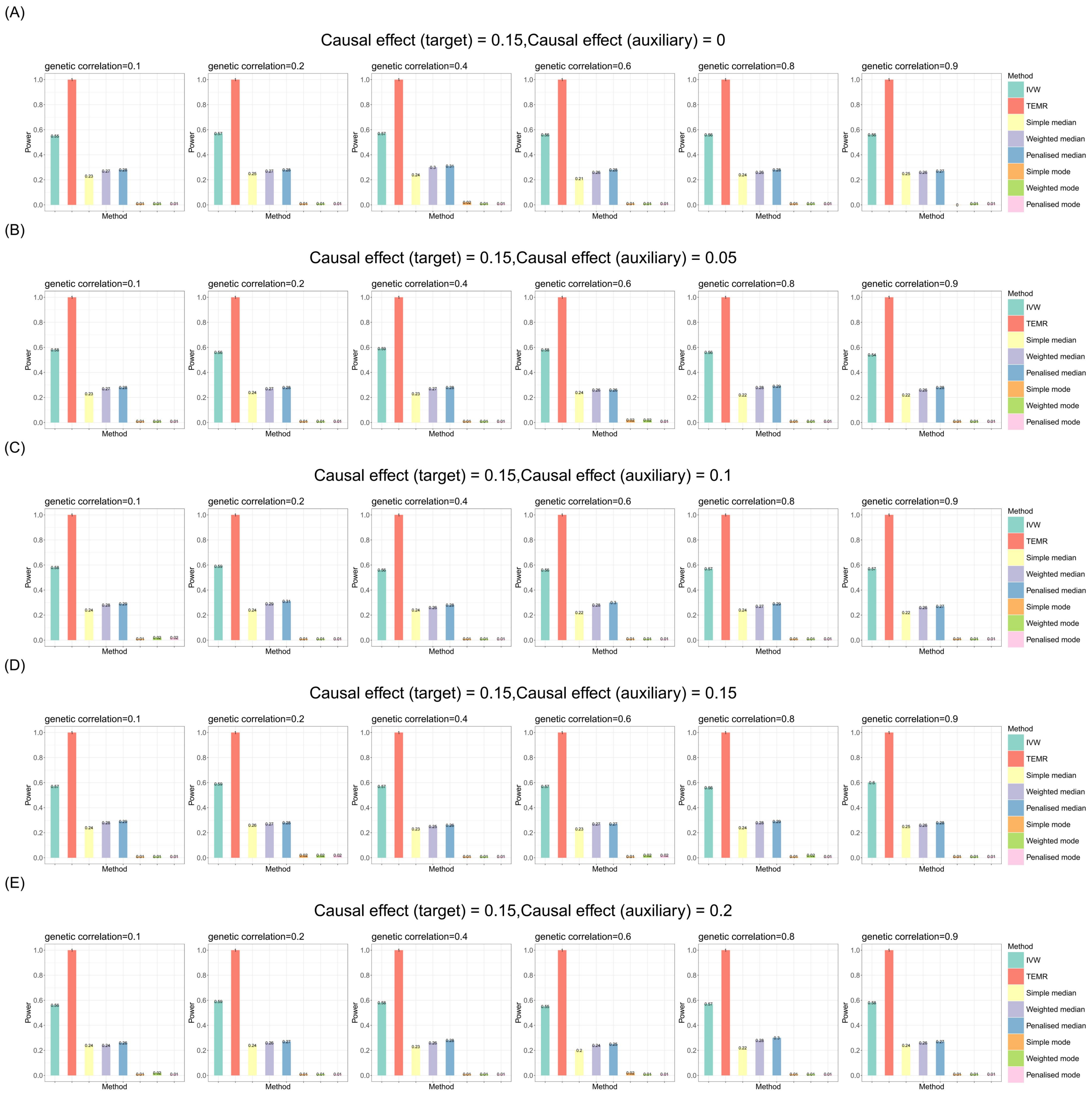


**Figure S35. Bar chart plots of simulation results with categorical outcome for causal effect estimation in the target population when there is one auxiliary population (directional horizontal pleiotropy).**

Sample size of target population is 3,000 and the sample size of auxiliary population is 300,000. IVs include 100 common SNPs. Bar chart plots illustrate the statistical power performance when the causal effect estimation is 0.15 in the target population, and from 0 to 0.2 in the auxiliary population, respectively. IVW, Inverse-variance weighted method.

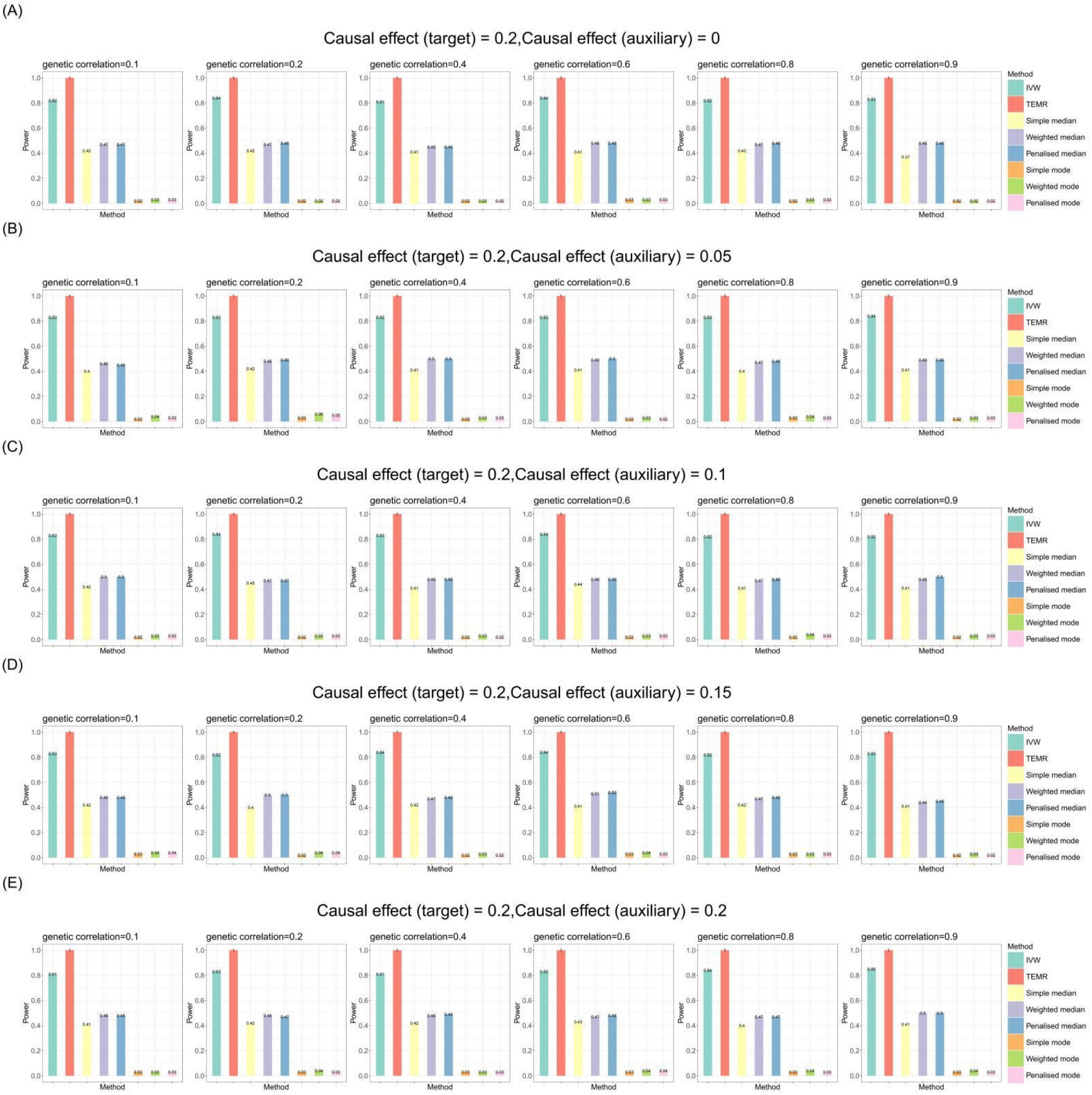


**Figure S36. Bar chart plots of simulation results with categorical outcome for causal effect estimation in the target population when there is one auxiliary population (directional horizontal pleiotropy).**

Sample size of target population is 3,000 and the sample size of auxiliary population is 300,000. IVs include 100 common SNPs. Bar chart plots illustrate the statistical power performance when the causal effect estimation is 0.2 in the target population, and from 0 to 0.2 in the auxiliary population, respectively. IVW, Inverse-variance weighted method.

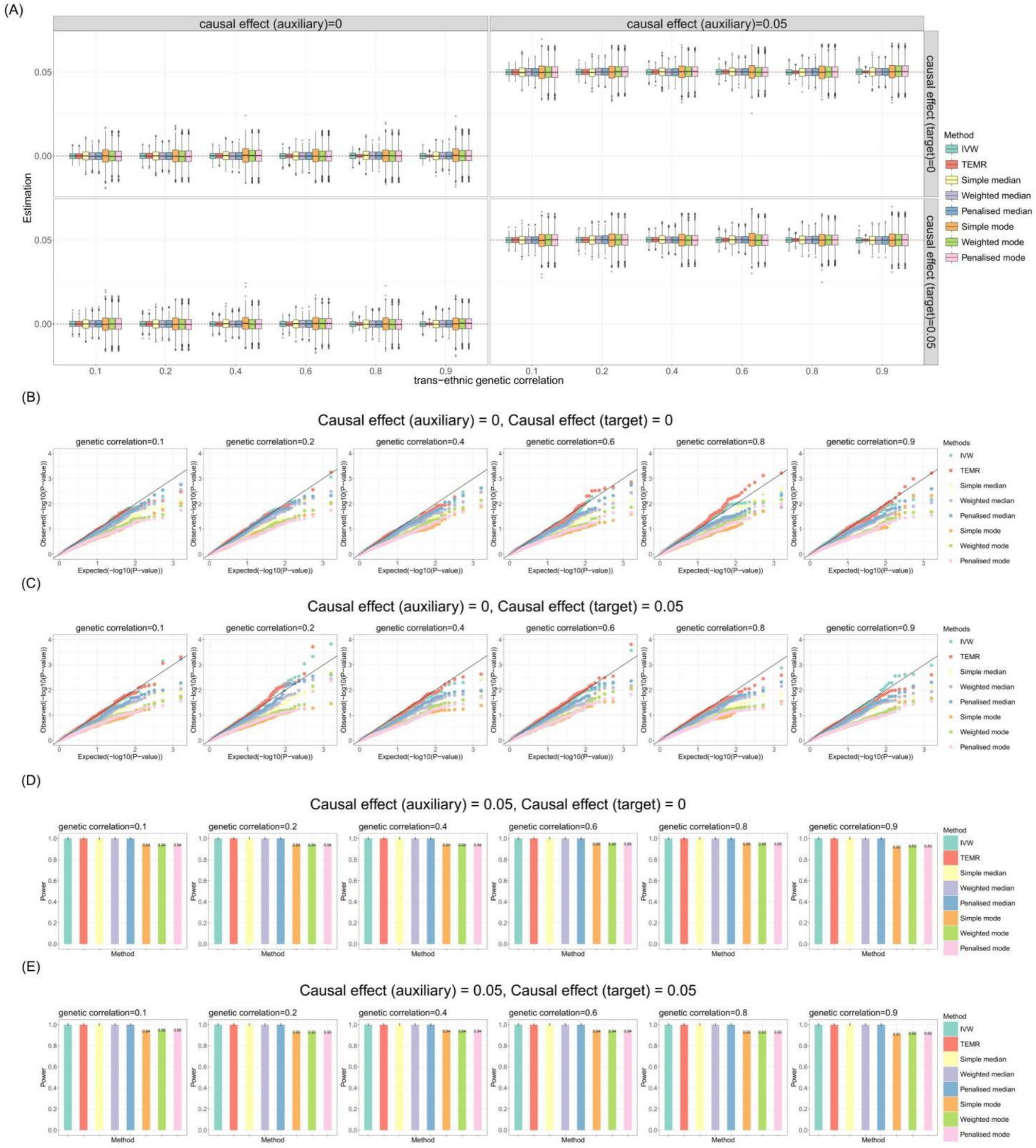


**Figure S37. Simulation results for causal effect estimation in the auxiliary population when there is one target population (no horizontal pleiotropy).**

Sample size of target population is 3,000 and the sample size of auxiliary population is 300,000. IVs include 100 common SNPs. **A)** Boxplots show the performances of causal effect estimation in auxiliary population; **B-C)** Q-Q plots show the performances of Type I error rates of zero causal effect estimation in auxiliary population when the causal effect of target population is 0 and 0.05, respectively; **D-E)** Bar chart plots show the performances of statistical power of non-zero causal effect estimation in auxiliary population when the causal effect of target population is 0 and 0.05, respectively. IVW, Inverse-variance weighted method

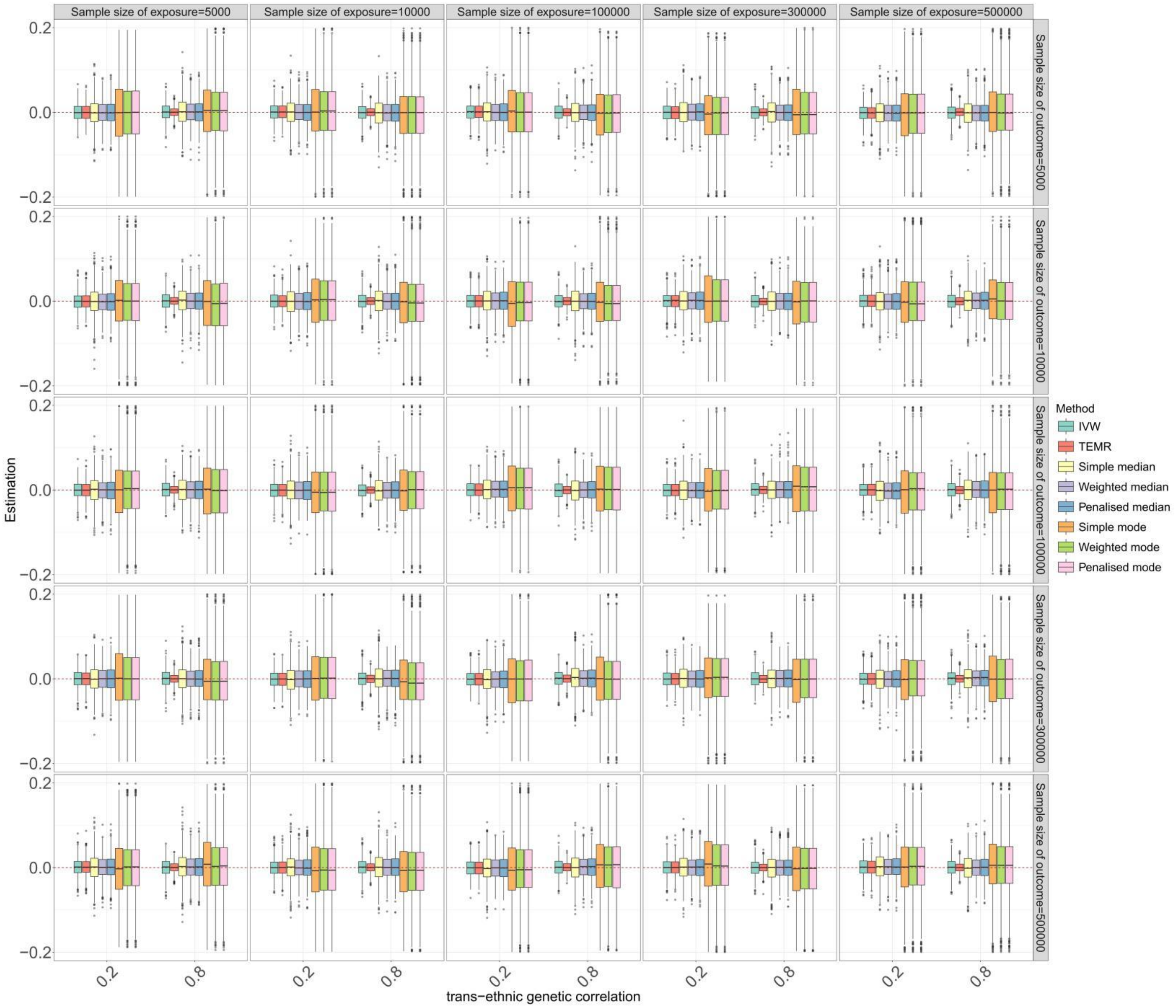


**Figure S38. Boxplots of simulation results for causal effect estimation in the target population with different sample sizes of exposure and outcome in auxiliary population.**

Continuous outcome, no horizontal pleiotropy. Sample size of target population is 3,000, and IVs include 100 common SNPs. Boxplots show the performances of causal effect estimation in target population when the causal effects of target population and auxiliary population are both 0. IVW, Inverse-variance weighted method

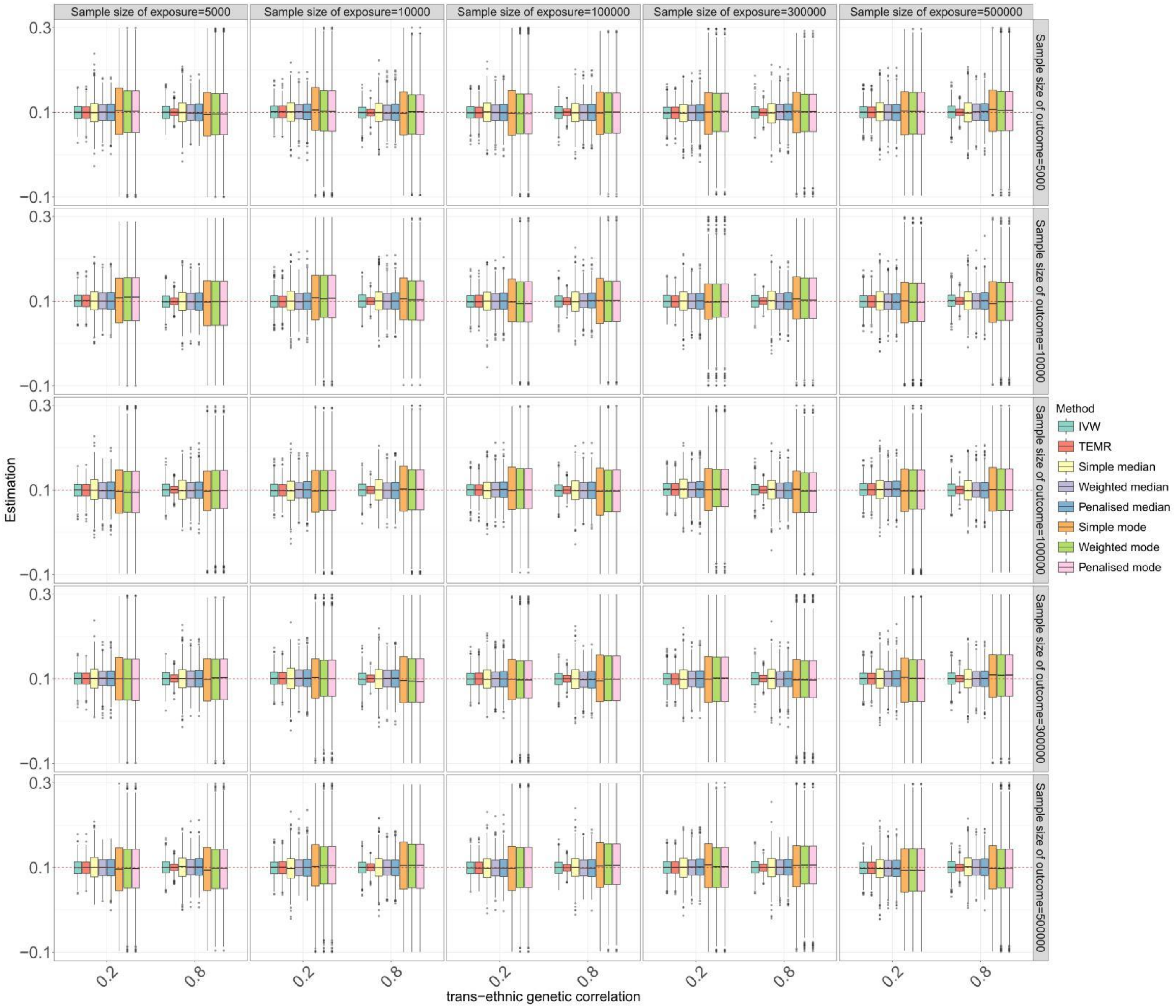


**Figure S39. Boxplots of simulation results for causal effect estimation in the target population with different sample sizes of exposure and outcome in auxiliary population.**

Continuous outcome, no horizontal pleiotropy. Sample size of target population is 3,000, and IVs include 100 common SNPs. Boxplots show the performances of causal effect estimation in target population when the causal effects of target population and auxiliary population are both 0.1. IVW, Inverse-variance weighted method

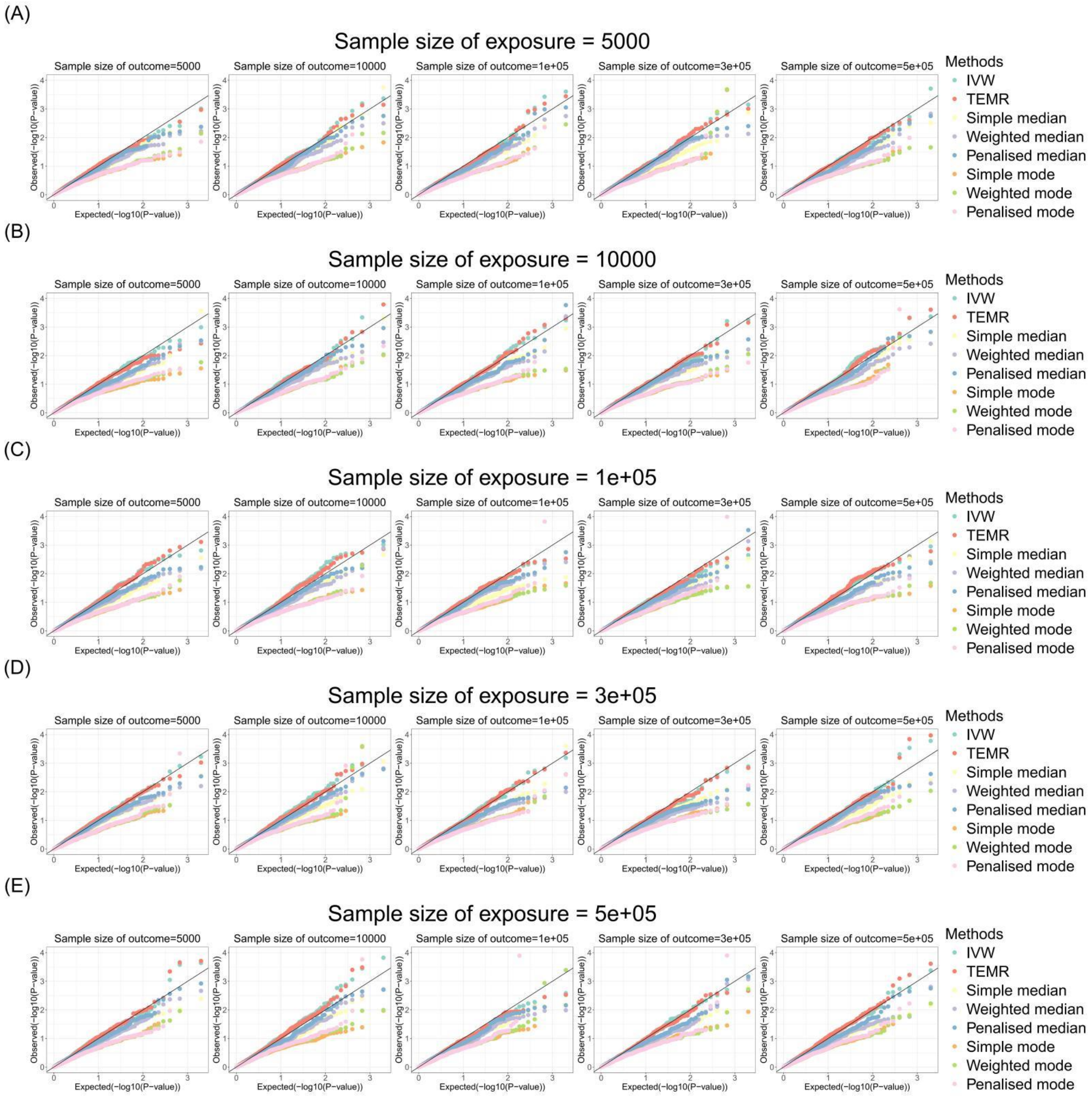


**Figure S40. Q-Q plots of simulation results for causal effect estimation in the target population with different sample sizes of exposure and outcome in auxiliary population (genetic correlation is 0.2).**

Continuous outcome, no horizontal pleiotropy. Sample size of target population is 3,000 and IVs include 100 common SNPs. Q-Q plots show the performances of Type I error rates of zero causal effect estimation in target population when the causal effects of target population and auxiliary population are both 0. IVW, Inverse-variance weighted method

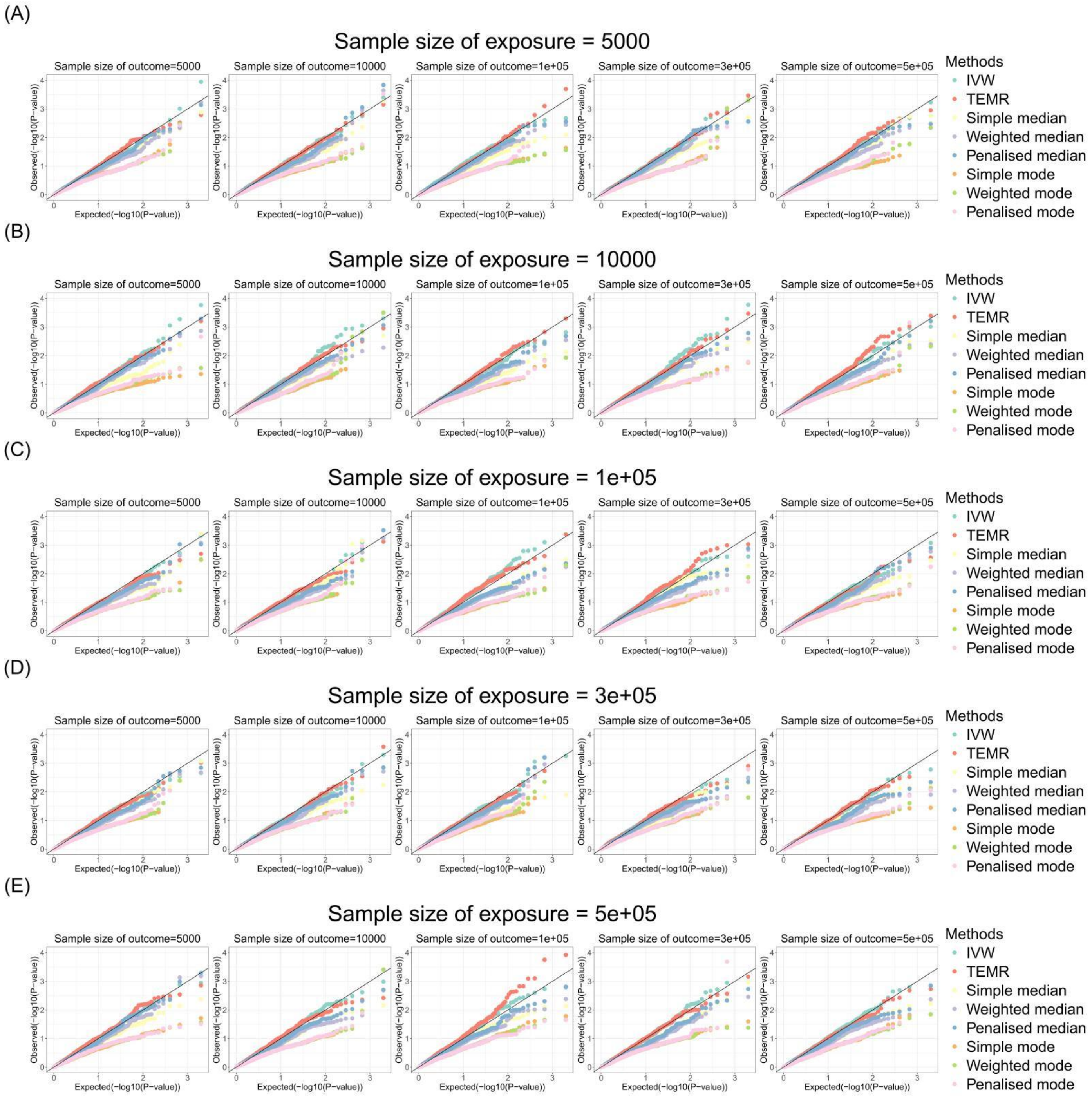


**Figure S41. Q-Q plots of simulation results for causal effect estimation in the target population with different sample sizes of exposure and outcome in auxiliary population (genetic correlation is 0.8).**

Continuous outcome, no horizontal pleiotropy. Sample size of target population is 3,000 and IVs include 100 common SNPs. Q-Q plots show the performances of Type I error rates of zero causal effect estimation in target population when the causal effects of target population and auxiliary population are both 0. IVW, Inverse-variance weighted method

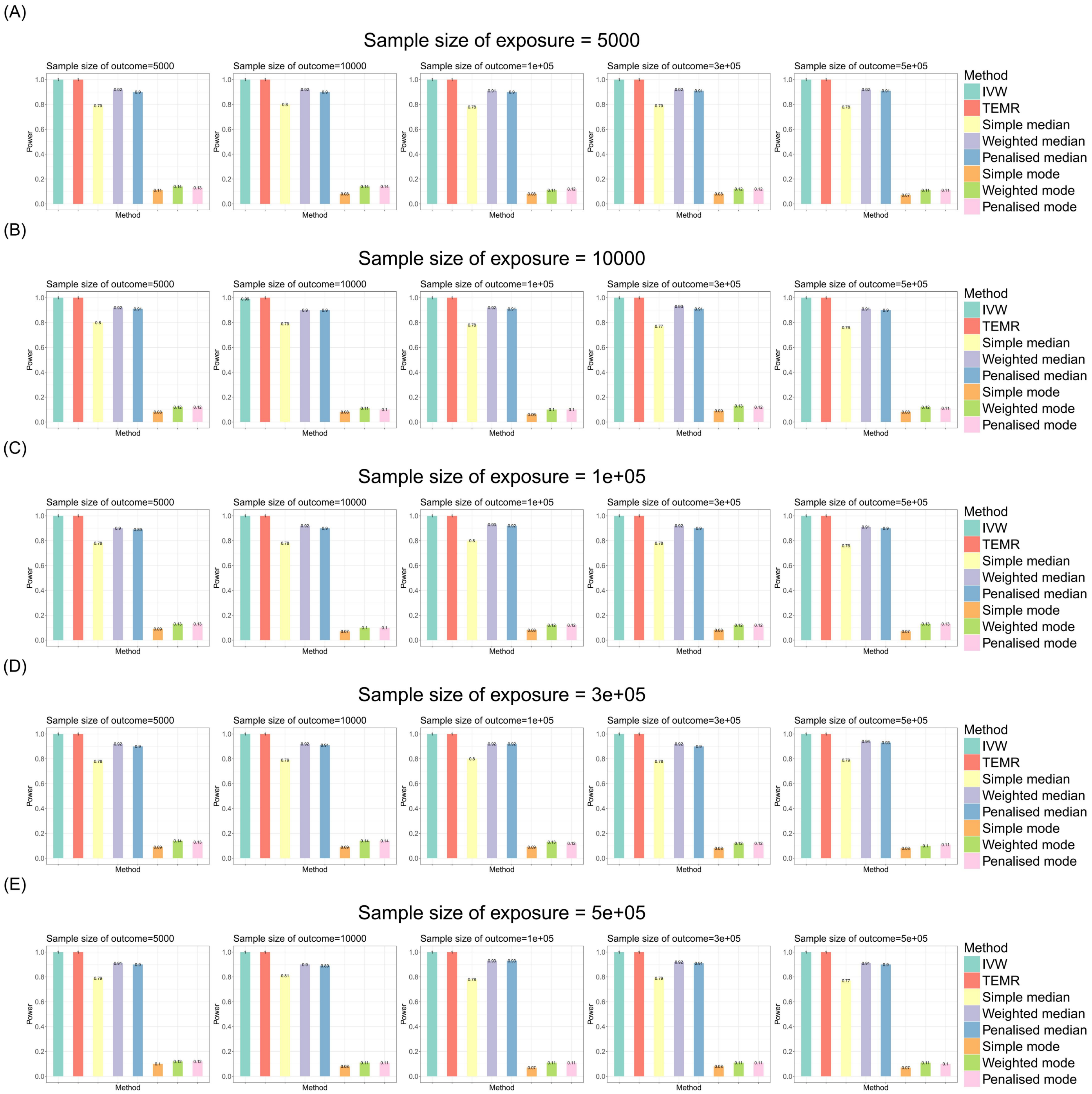


**Figure S42. Bar chart plots of simulation results for causal effect estimation in the target population with different sample sizes of exposure and outcome in auxiliary population (genetic correlation is 0.2).**

Continuous outcome, no horizontal pleiotropy. Sample size of target population is 3,000 and IVs include 100 common SNPs. Bar chart plots show the performances of statistical power of non-zero causal effect estimation in target population when the causal effects of target population and auxiliary population are both 0.1. IVW, Inverse-variance weighted method.

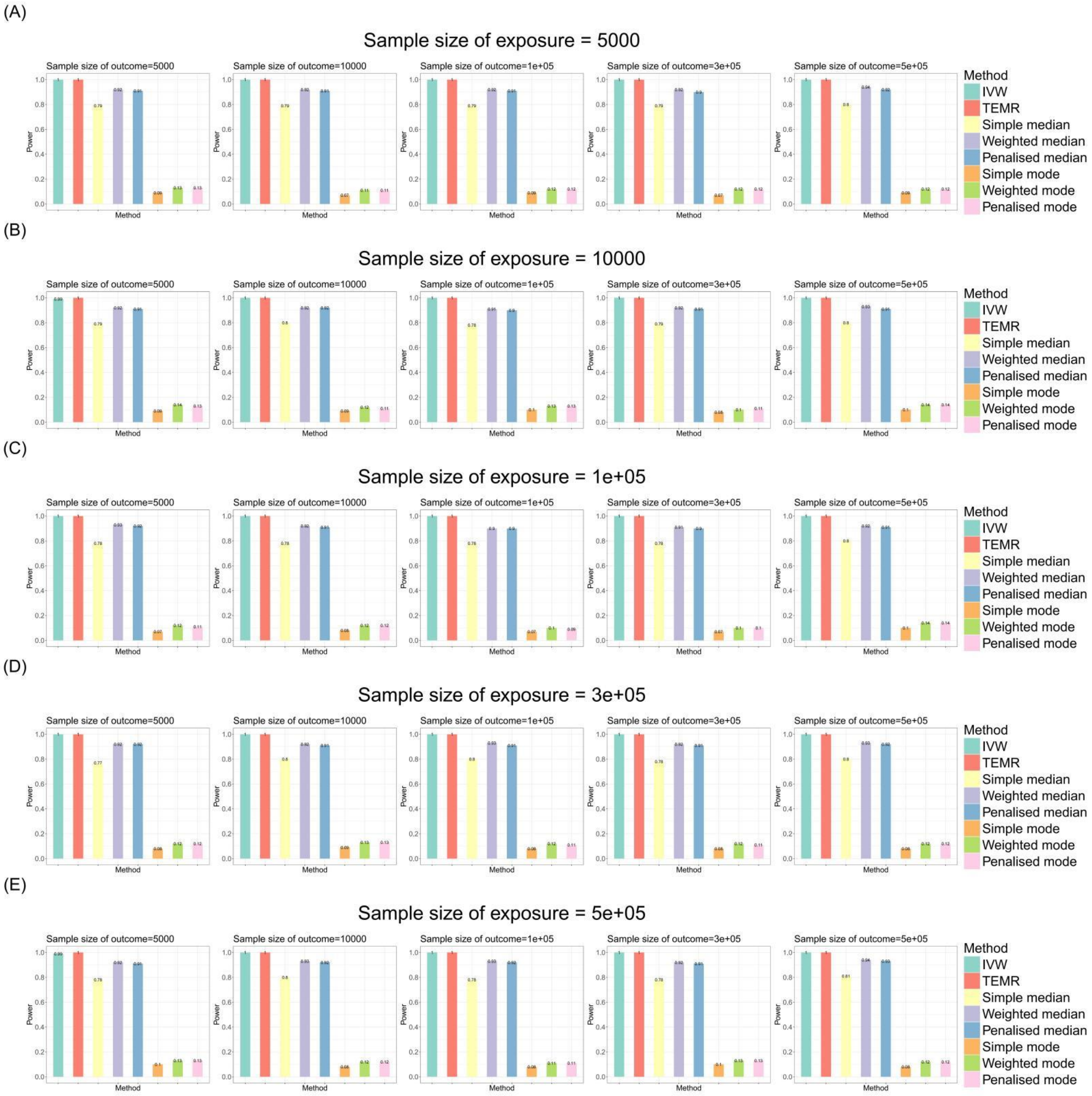


**Figure S43. Bar chart plots of simulation results for causal effect estimation in the target population with different sample sizes of exposure and outcome in auxiliary population (genetic correlation is 0.8).**

Continuous outcome, no horizontal pleiotropy. Sample size of target population is 3,000 and IVs include 100 common SNPs. Bar chart plots show the performances of statistical power of non-zero causal effect estimation in target population when the causal effects of target population and auxiliary population are both 0.1. IVW, Inverse-variance weighted method.

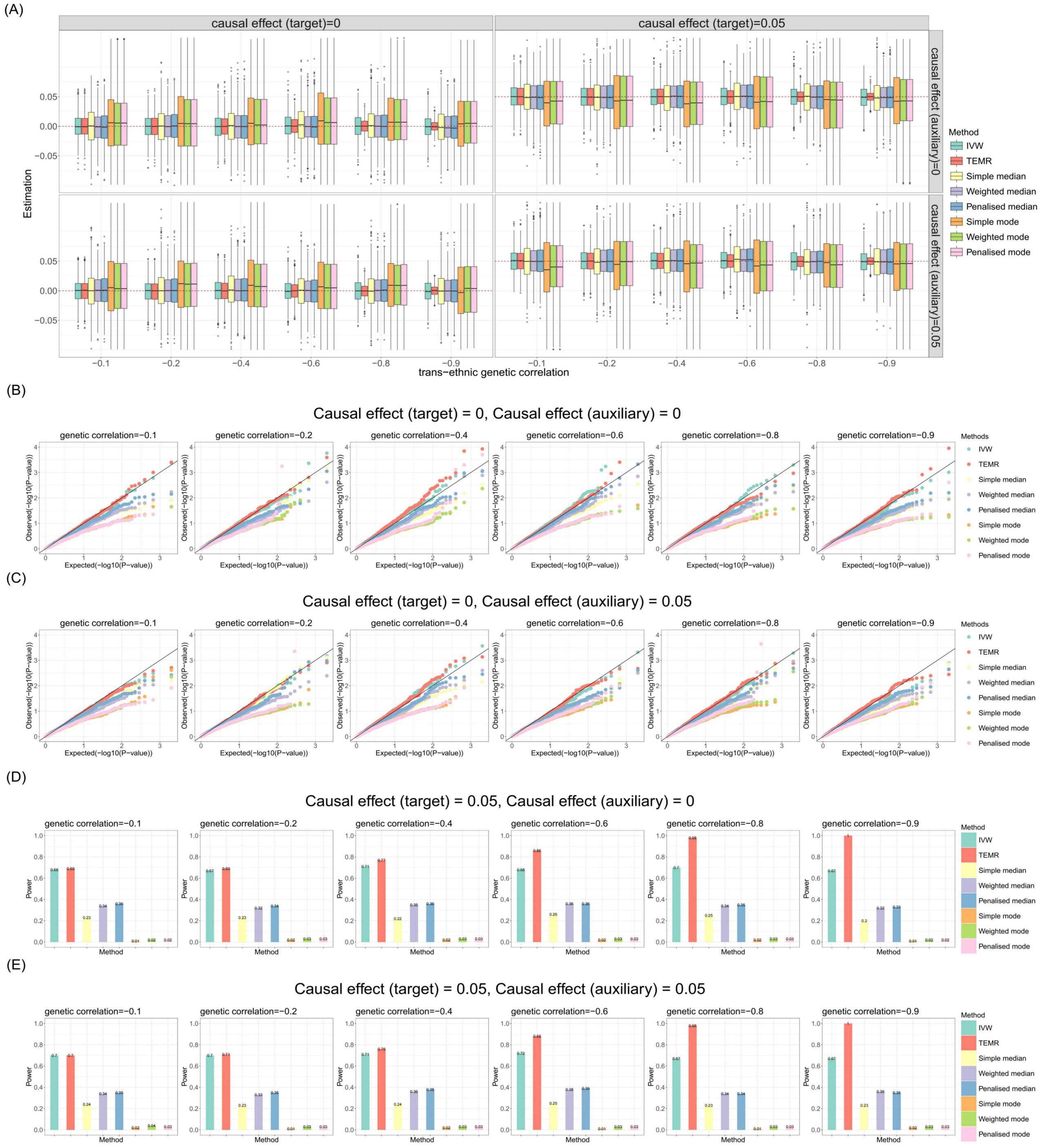


**Figure 44. Simulation results for causal effect estimation in the target population when there is one auxiliary population with negative genetic correlations (no pleiotropy).**

Sample size of target population is 3,000 and the sample size of auxiliary population is 300,000. IVs include 100 common SNPs. A) Boxplots show the performances of causal effect estimation in target population; B-C) Q-Q plots show the performances of Type I error rates of zero causal effect estimation in target population when the causal effect of auxiliary population is 0 and 0.05, respectively; D-E) Bar chart plots show the performances of statistical power of non-zero causal effect estimation in target population when the causal effect of auxiliary population is 0 and 0.05, respectively. IVW, Inverse-variance weighted method.

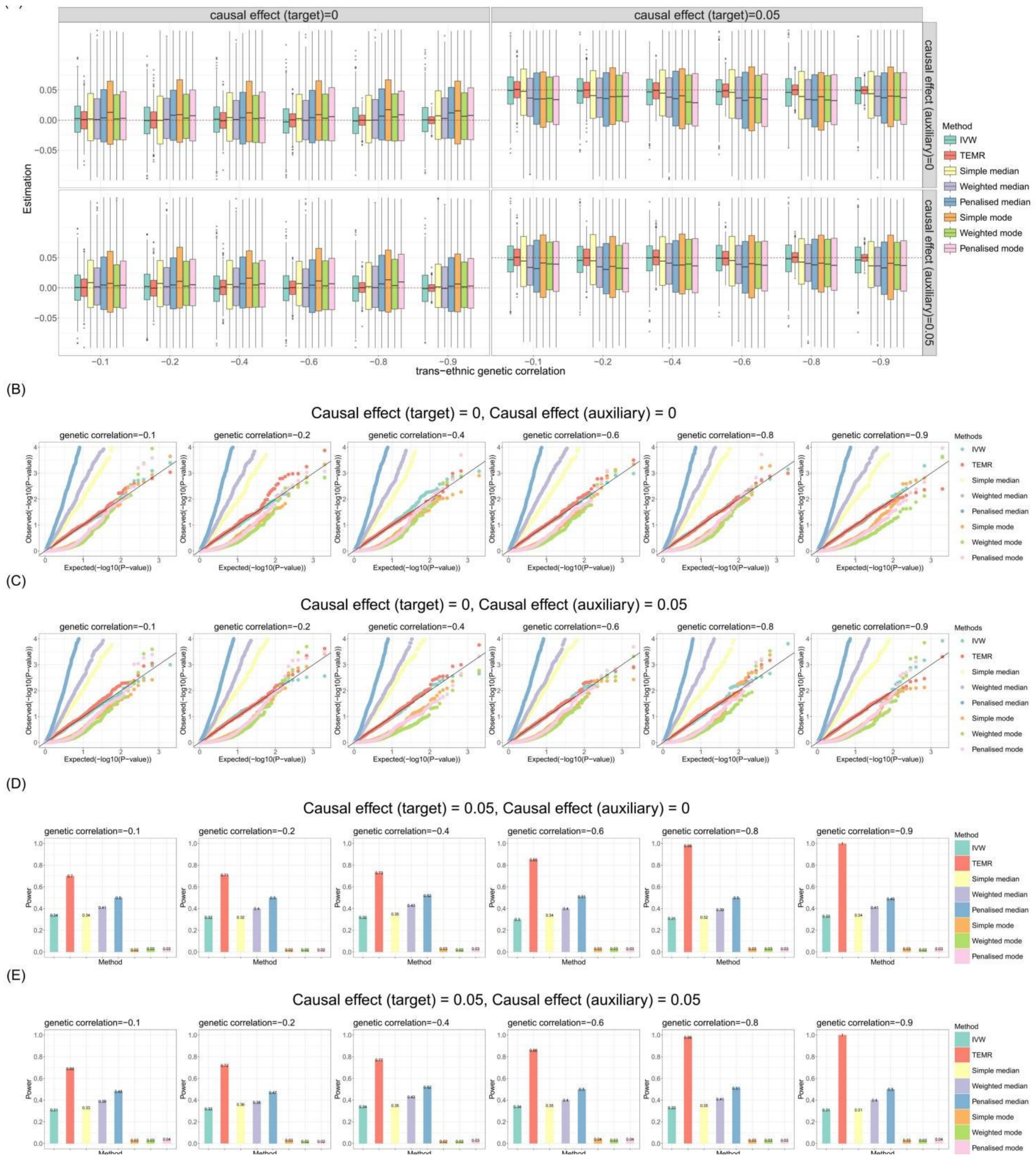


**Figure 45. Simulation results for causal effect estimation in the target population when there is one auxiliary population with negative genetic correlations (directional pleiotropy).**

Sample size of target population is 3,000 and the sample size of auxiliary population is 300,000. IVs include 100 common SNPs. A) Boxplots show the performances of causal effect estimation in target population; B-C) Q-Q plots show the performances of Type I error rates of zero causal effect estimation in target population when the causal effect of auxiliary population is 0 and 0.05, respectively; D-E) Bar chart plots show the performances of statistical power of non-zero causal effect estimation in target population when the causal effect of auxiliary population is 0 and 0.05, respectively. IVW, Inverse-variance weighted method.

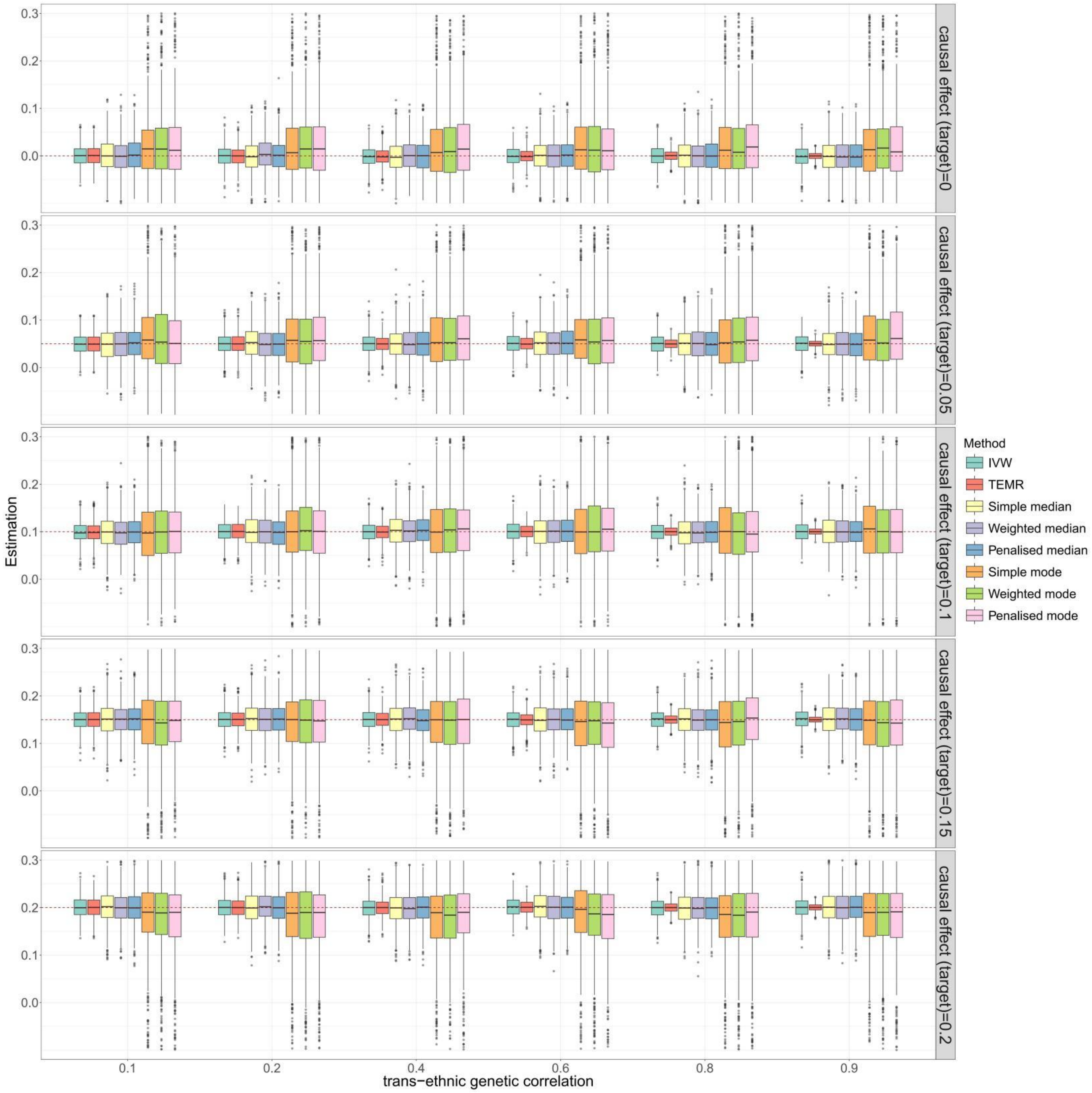


**Figure S46. Boxplots of simulation results with continuous outcome for causal effect estimation in the target population when there are three auxiliary population (no horizontal pleiotropy).** Sample size of target population is 3,000 and the sample size of auxiliary populations are 3,000, 3,000, 300,000. IVs include 100 common SNPs. Boxplots show the performances of causal effect estimation in target population when the causal effect of target popultion and auxiliary population are both from 0 to 0.2, respectively. IVW, Inverse-variance weighted method.

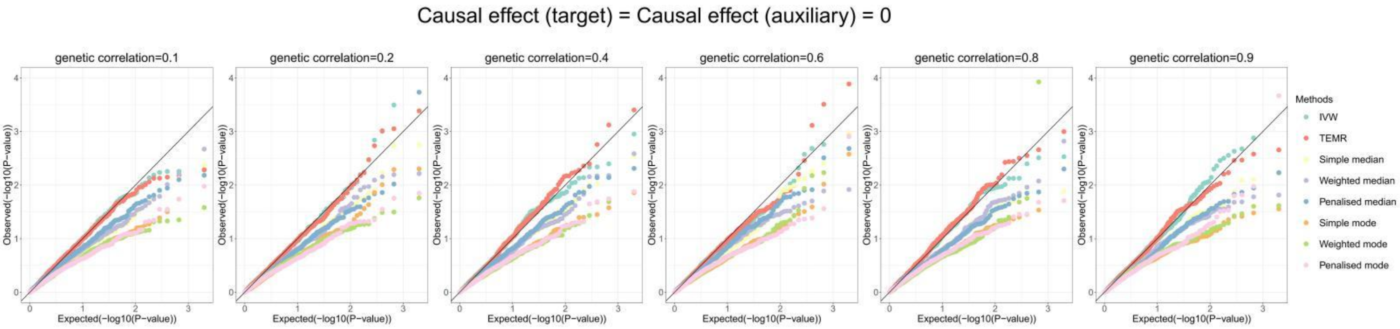


**Figure S47. Q-Q plots of simulation results with continuous outcome for causal effect estimation in the target population when there is one auxiliary population (no horizontal pleiotropy).**

Sample size of target population is 3,000 and the sample size of auxiliary populations are 3,000, 3,000, 300,000. IVs include 100 common SNPs. Q-Q plots show the performances of Type I error rates of zero causal effect estimation in target population when the causal effect of auxiliary population is also 0. IVW, Inverse-variance weighted method.

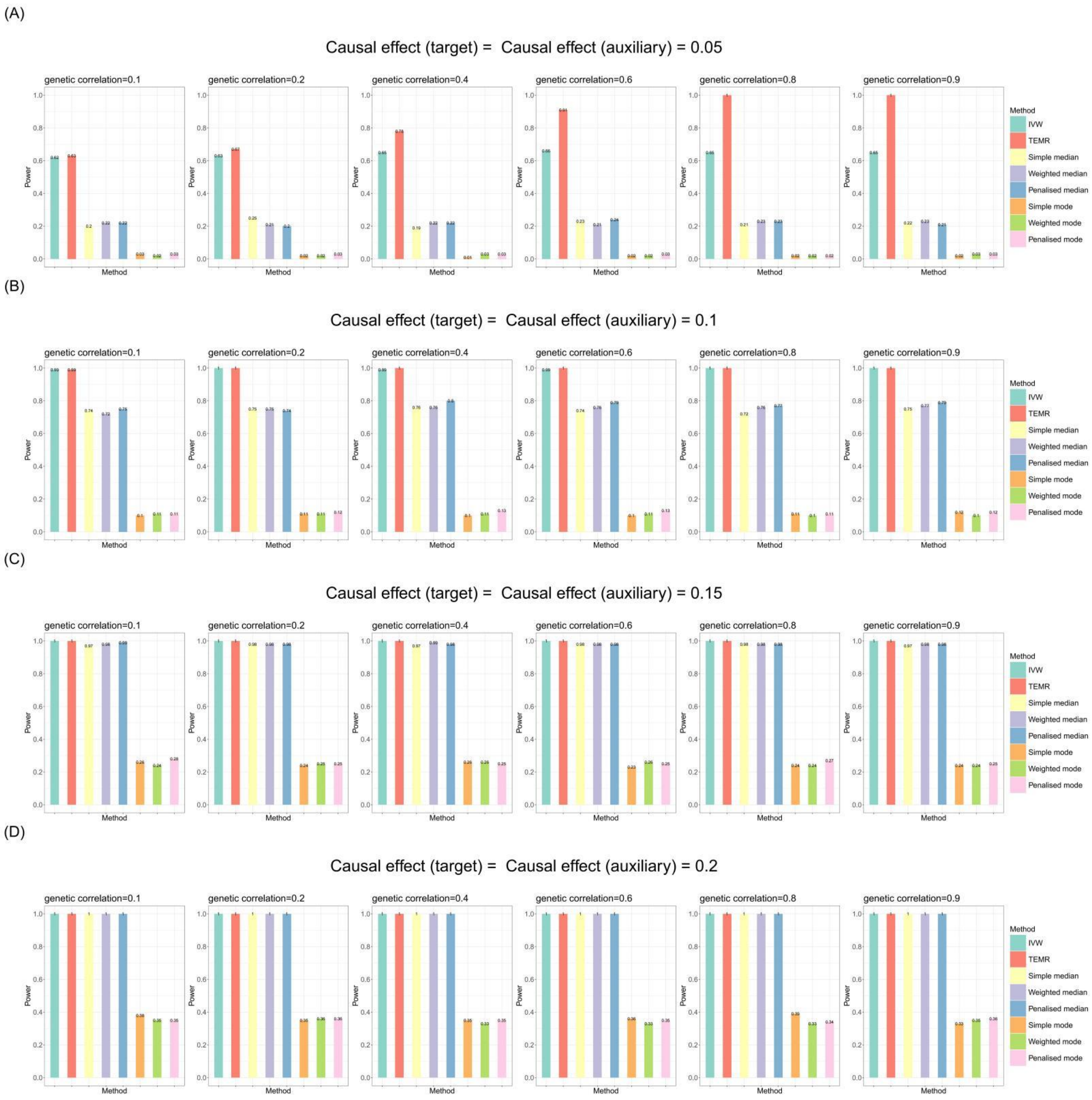


**Figure S48. Bar chart plots of simulation results with continuous outcome for causal effect estimation in the target population when there is one auxiliary population (no horizontal pleiotropy).**

Sample size of target population is 3,000 and the sample size of auxiliary populations are 3,000, 3,000, 300,000. IVs include 100 common SNPs. Bar chart plots illustrate the statistical power performance when the causal effect estimations in the target population and in the auxiliary population are both from 0 to 0.2, respectively. IVW, Inverse-variance weighted method.

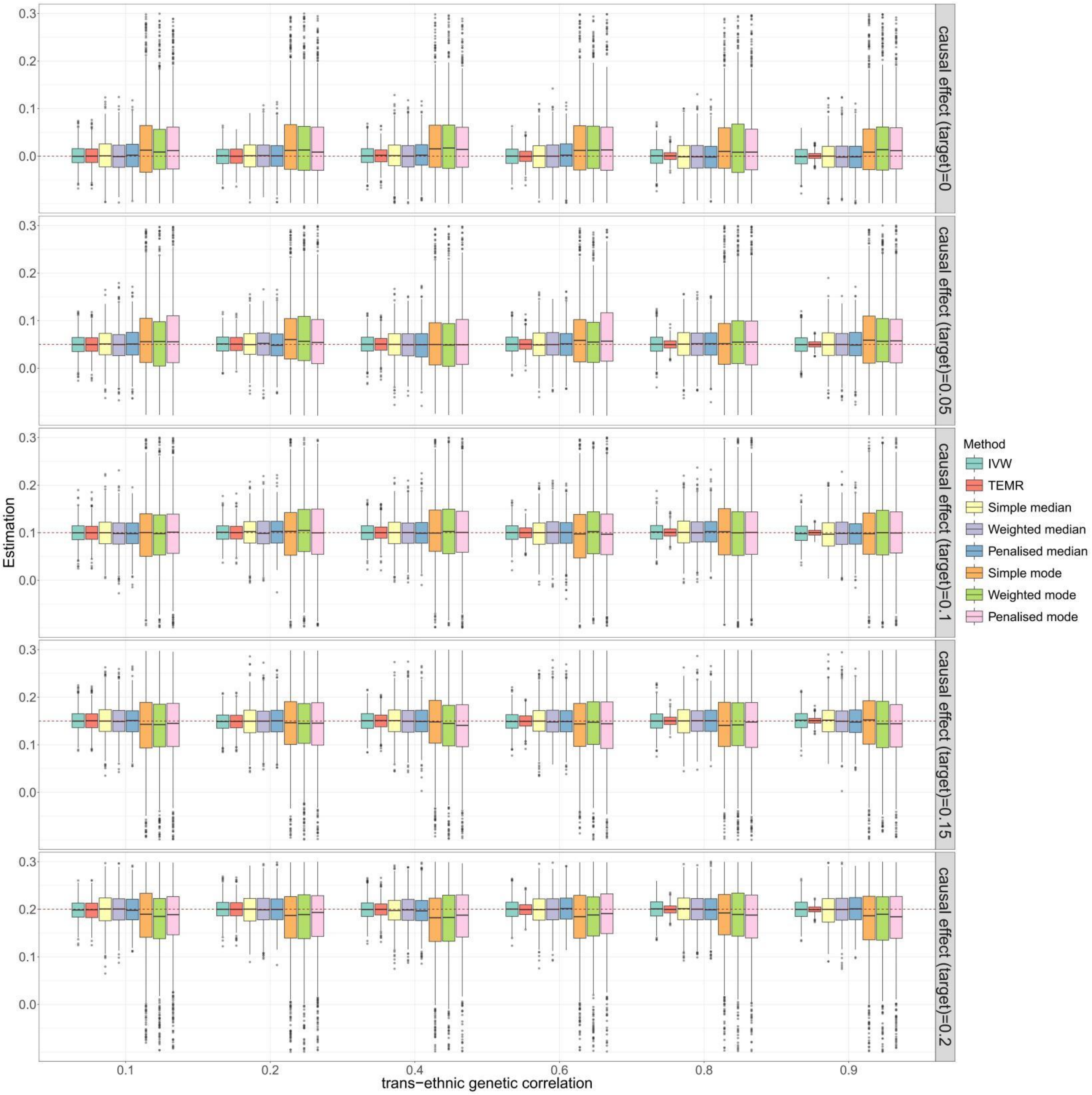


**Figure S49. Boxplots of simulation results with continuous outcome for causal effect estimation in the target population when there are three auxiliary population (balance horizontal pleiotropy).**

Sample size of target population is 3,000 and the sample size of auxiliary populations are 3,000, 3,000, 300,000. IVs include 100 common SNPs. Boxplots show the performances of causal effect estimation in target population when the causal effect of target popultion and auxiliary population are both from 0 to 0.2, respectively. IVW, Inverse-variance weighted method.

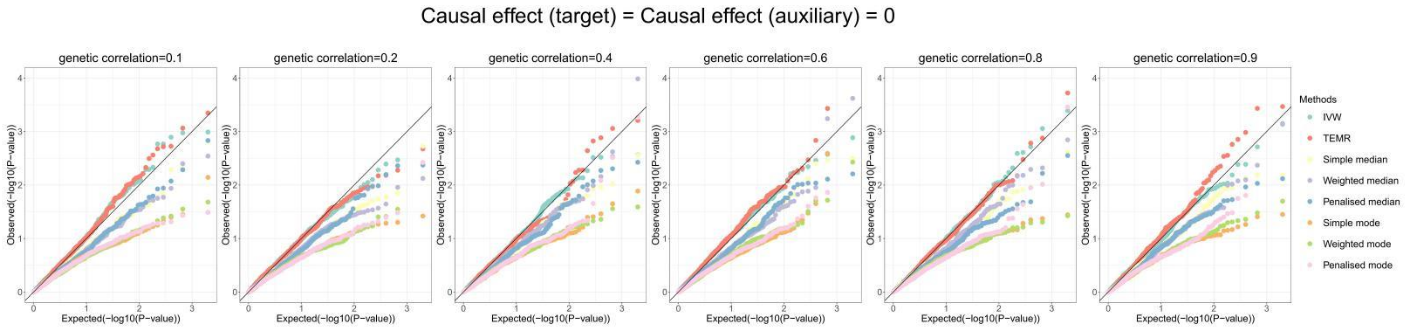


**Figure S50. Q-Q plots of simulation results with continuous outcome for causal effect estimation in the target population when there is one auxiliary population (balance horizontal pleiotropy).**

Sample size of target population is 3,000 and the sample size of auxiliary populations are 3,000, 3,000, 300,000. IVs include 100 common SNPs. Q-Q plots show the performances of Type I error rates of zero causal effect estimation in target population when the causal effect of auxiliary population is also 0. IVW, Inverse-variance weighted method.

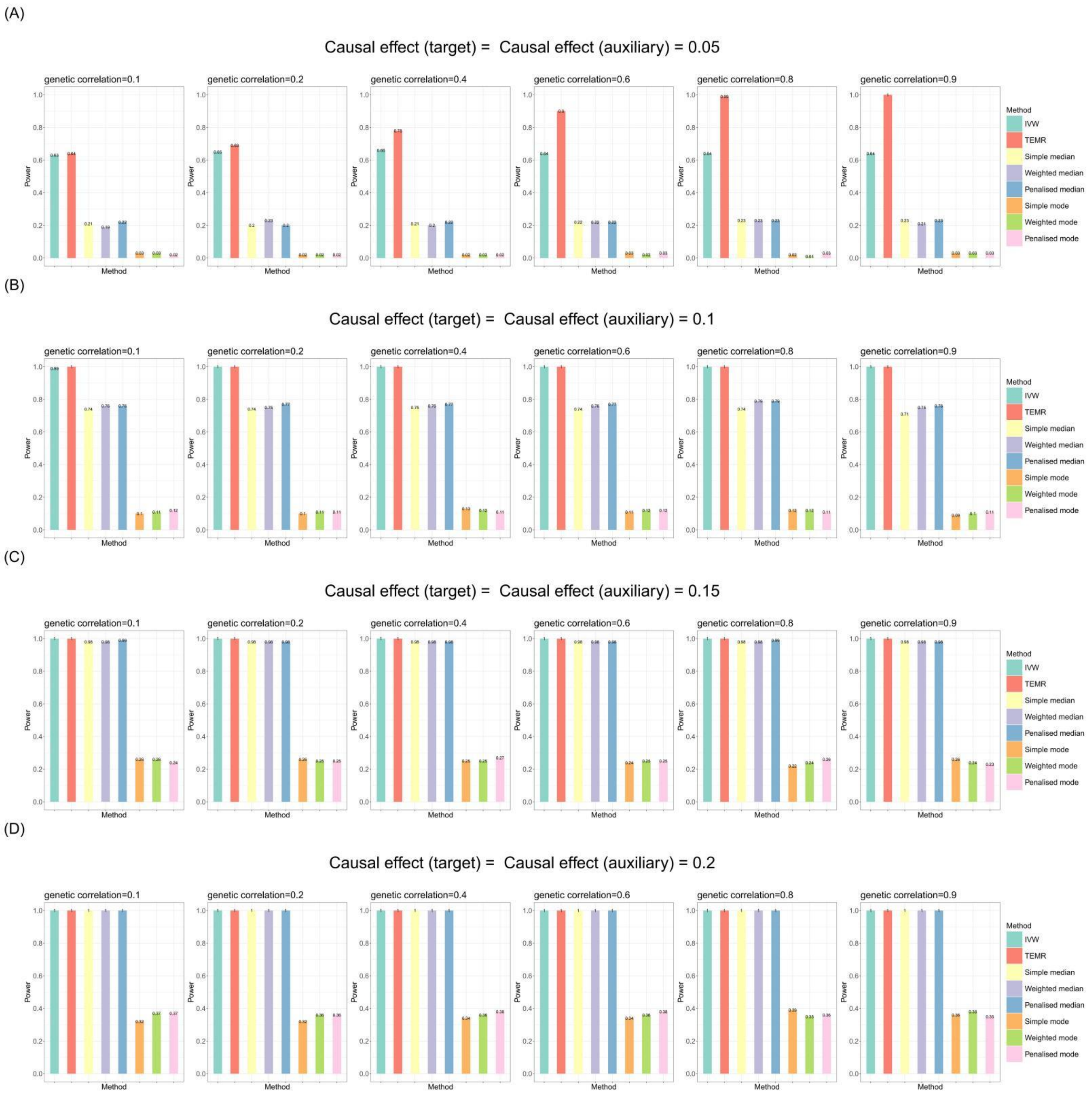


**Figure S51. Bar chart plots of simulation results with continuous outcome for causal effect estimation in the target population when there is one auxiliary population (balance horizontal pleiotropy).**

Sample size of target population is 3,000 and the sample size of auxiliary populations are 3,000, 3,000, 300,000. IVs include 100 common SNPs. Bar chart plots illustrate the statistical power performance when the causal effect estimations in the target population and in the auxiliary population are both from 0 to 0.2, respectively. IVW, Inverse-variance weighted method.

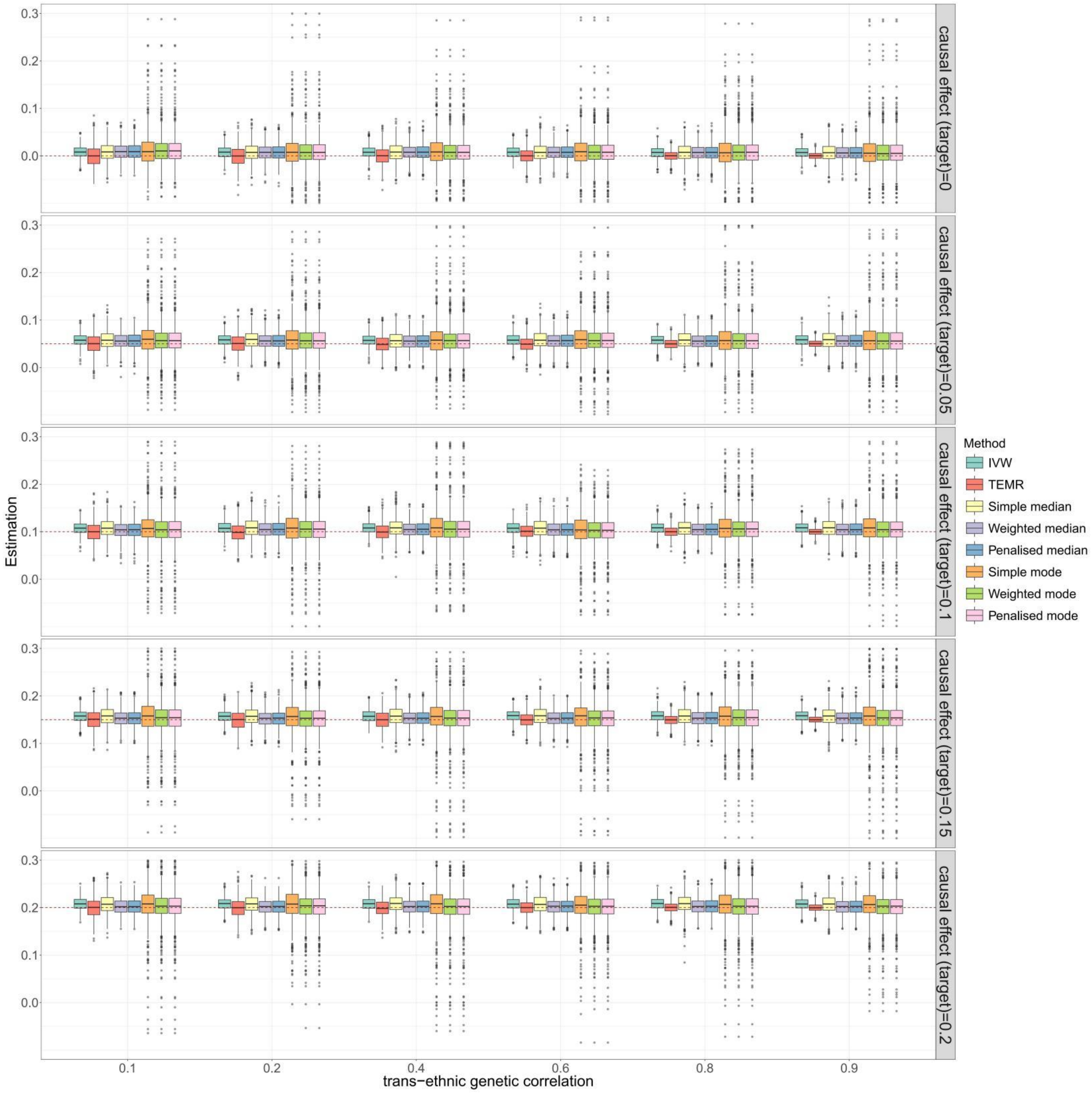


**Figure S52. Boxplots of simulation results with continuous outcome for causal effect estimation in the target population when there are three auxiliary population (directional horizontal pleiotropy).**

Sample size of target population is 3,000 and the sample size of auxiliary populations are 3,000, 3,000, 300,000. IVs include 100 common SNPs. Boxplots show the performances of causal effect estimation in target population when the causal effect of target popultion and auxiliary population are both 0 to 0.2, respectively. IVW, Inverse-variance weighted method.

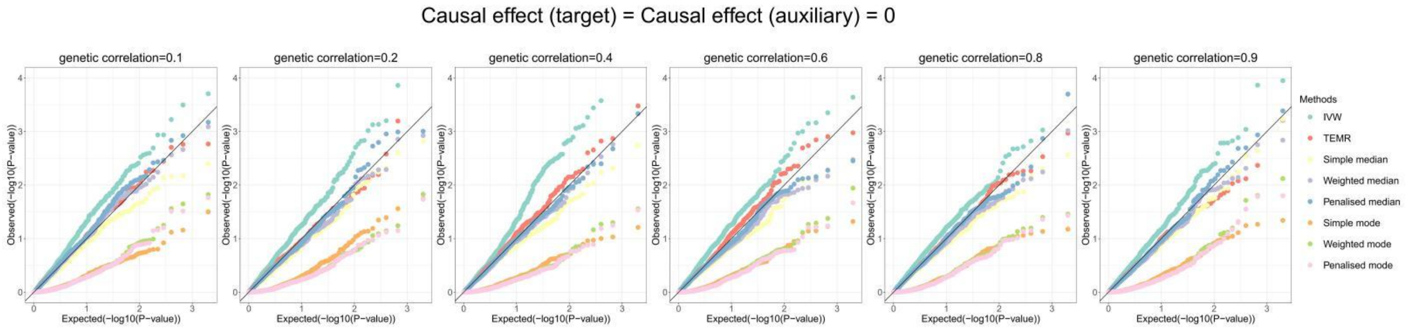


**Figure S53. Q-Q plots of simulation results with continuous outcome for causal effect estimation in the target population when there is one auxiliary population (directional horizontal pleiotropy).**

Sample size of target population is 3,000 and the sample size of auxiliary populations are 3,000, 3,000, 300,000. IVs include 100 common SNPs. Q-Q plots show the performances of Type I error rates of zero causal effect estimation in target population when the causal effect of auxiliary population is also 0. IVW, Inverse-variance weighted method.

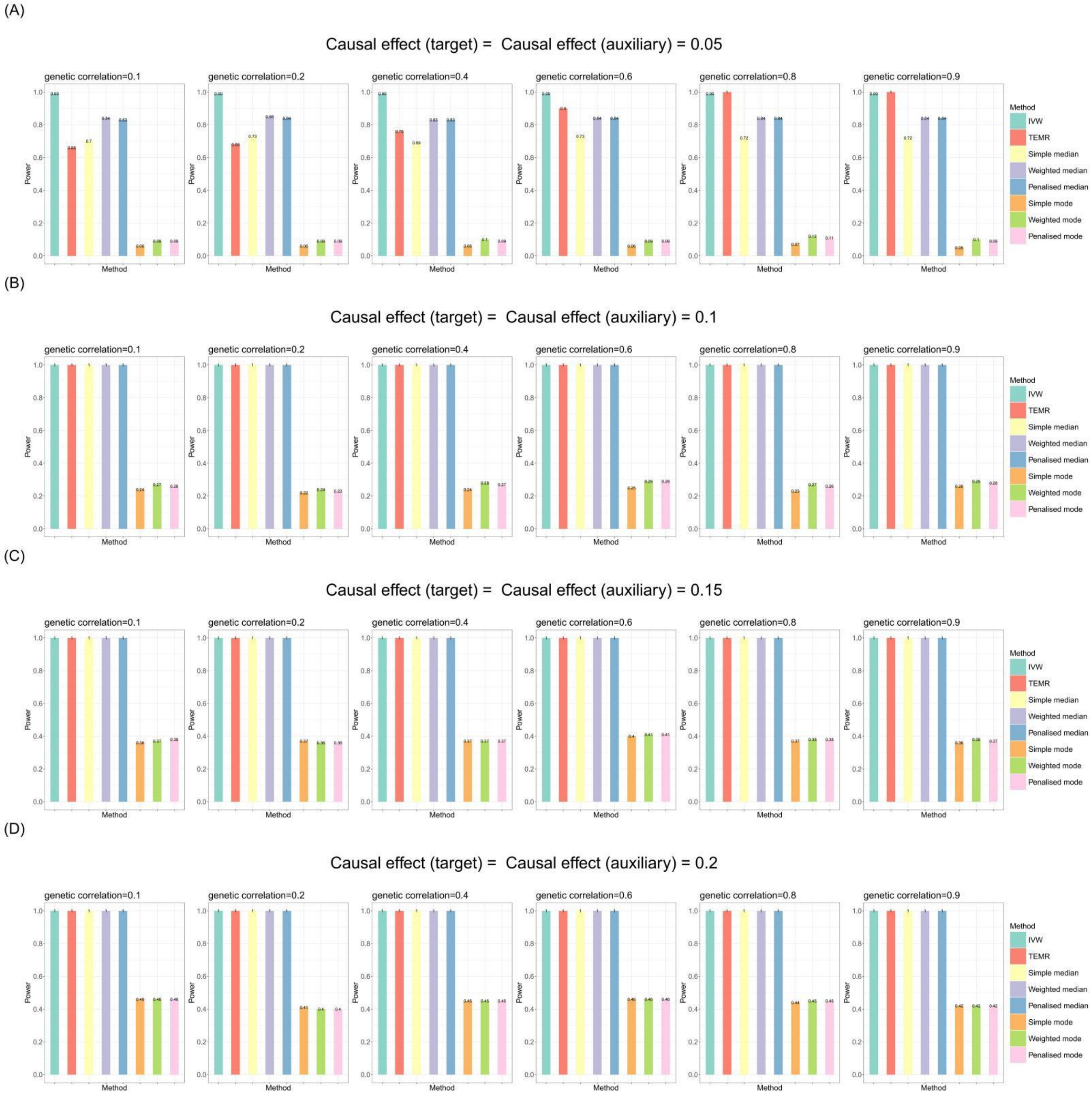


**Figure S54. Bar chart plots of simulation results with continuous outcome for causal effect estimation in the target population when there is one auxiliary population (directional horizontal pleiotropy).**

Sample size of target population is 3,000 and the sample size of auxiliary populations are 3,000, 3,000, 300,000. IVs include 100 common SNPs. Bar chart plots illustrate the statistical power performance when the causal effect estimations in the target population and in the auxiliary population are both from 0 to 0.2, respectively. IVW, Inverse-variance weighted method.

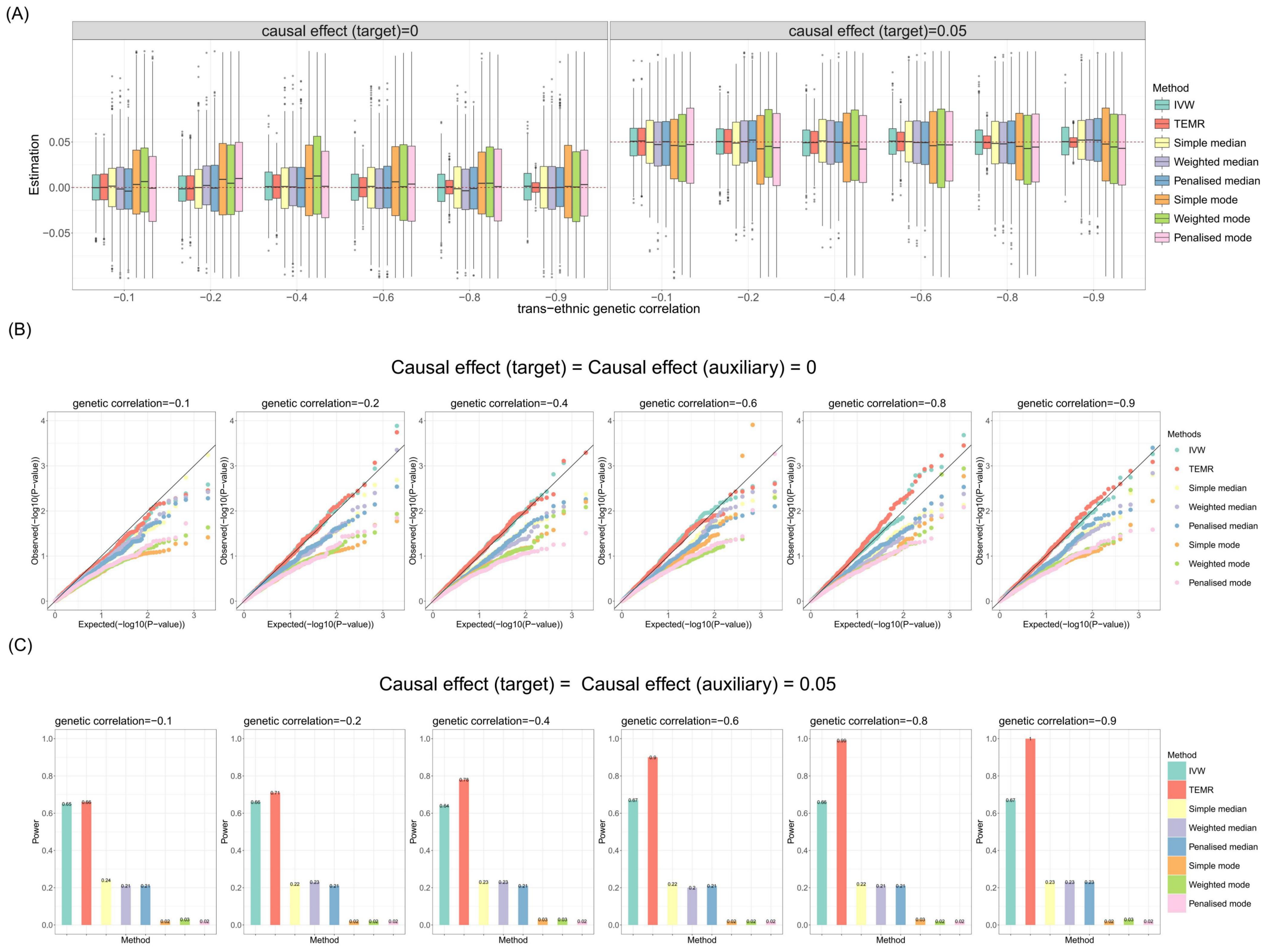


**Figure 55. Simulation results for causal effect estimation in the target population when there is three auxiliary population with different genetic correlations (no pleiotropy).**

Continuous outcome. Sample size of target population is 3,000, and the sample size of auxiliary populations are 3,000, 3,000, 300,000. IVs include 100 common SNPs. A) Boxplots show the performances of causal effect estimation in target population; B) Q-Q plots show the performances of Type I error rates of zero causal effect estimation in target population; C) Bar chart plots show the performances of statistical power of non-zero causal effect estimation in target population. IVW, Inverse-variance weighted method.

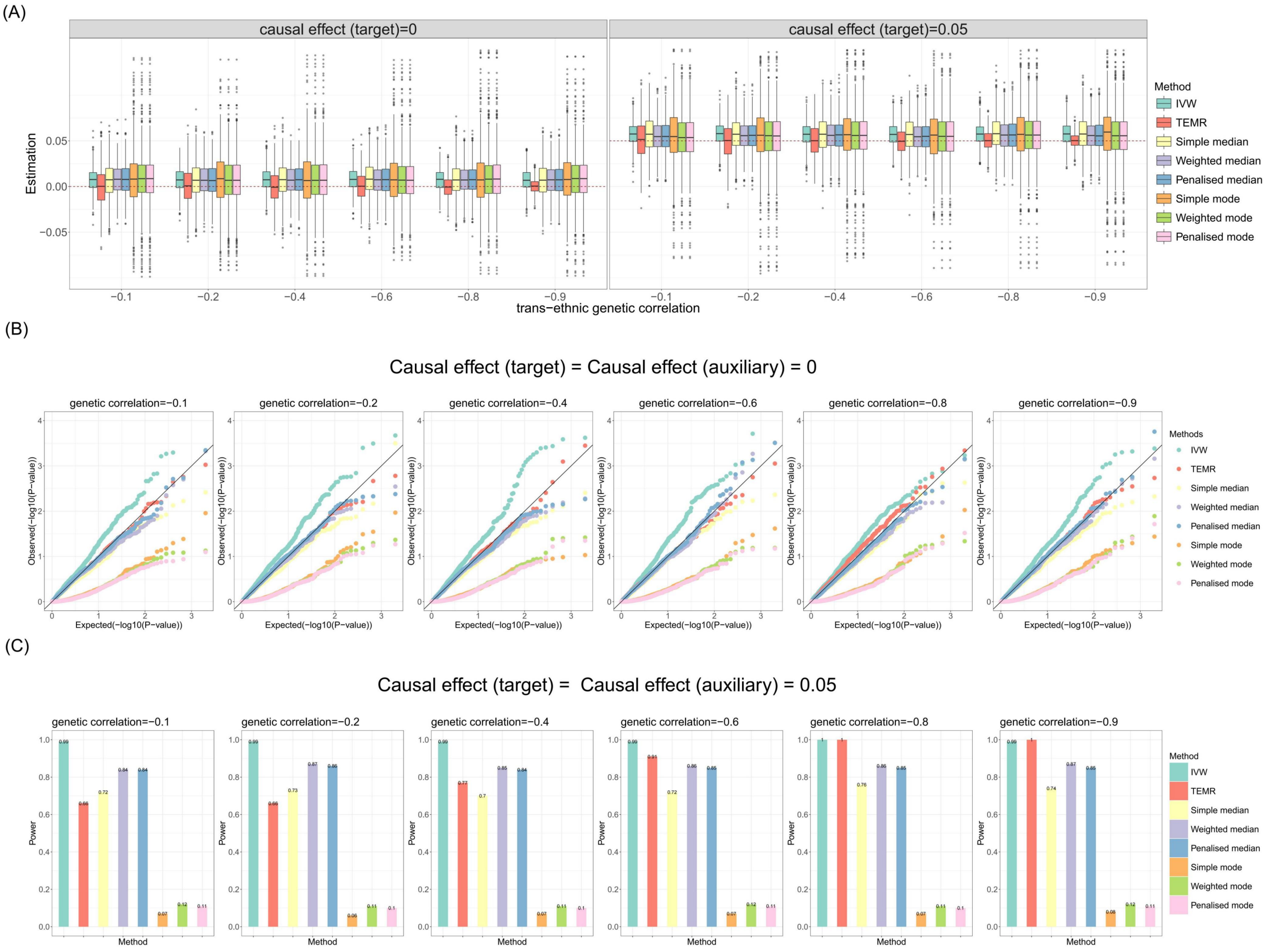


**Figure 56. Simulation results for causal effect estimation in the target population when there is three auxiliary population with different genetic correlations (directional pleiotropy).**

Continuous outcome. Sample size of target population is 3,000, and the sample size of auxiliary populations are 3,000, 3,000, 300,000. IVs include 100 common SNPs. A) Boxplots show the performances of causal effect estimation in target population; B) Q-Q plots show the performances of Type I error rates of zero causal effect estimation in target population; C) Bar chart plots show the performances of statistical power of non-zero causal effect estimation in target population. IVW, Inverse-variance weighted method.
